# DNA methylation-based biomarkers and prediction models for the survival of patients with colorectal cancer: systematic review and external validation study

**DOI:** 10.1101/2022.11.03.22281595

**Authors:** Tanwei Yuan, Dominic Edelmann, Jakob N. Kather, Ziwen Fan, Katrin E. Tagscherer, Wilfried Roth, Melanie Bewerunge-Hudler, Alexander Brobeil, Matthias Kloor, Hendrik Bläker, Barbara Burwinkel, Hermann Brenner, Michael Hoffmeister

## Abstract

**Objectives:** To identify existing DNA methylation-based prognostic biomarkers and prediction models for colorectal cancer (CRC) prognosis and to validate them in a large external cohort.

**Design:** Systematic review and external validation study.

**Data source:** Systematic search in PubMed and Web of Science until October 2022 to identify epigenome-wide studies reporting methylation at CpG sites (CpGs) associated with survival among CRC patients. Validation data were drawn from the 2310 CRC patients of the DACHS study recruited from 22 hospitals in the Rhine-Neckar region in the southwest of Germany.

**Main outcome measures:** Overall survival (OS) in CRC patients.

**Results:** We identified 200 unique CpGs and 10 CpG-based prognostic models derived from 15 studies. In the external validation analysis, 1252 of 2310 patients died during follow-up (median 10.4 years). Thirty-nine CpGs (20%) and five prognostic models (50%) were independently associated with overall survival after adjustment for clinical variables. The five models had unsatisfactory discrimination ability, with area under the receiver operating characteristic curves at five years ranging from 0.54 to 0.60. The calibration accuracy of the five models using recalibrated baseline survival was also poor, and no relevant added prognostic value to traditional clinical variables was observed. Based on the Prediction Model Risk of Bias Assessment Tool, all models were rated as high risk of bias.

**Conclusions:** Only 20% of published CpGs associated with survival in CRC patients could be externally validated. So far derived published CpG-based prognostic models for CRC do not seem to be useful for clinical practice.

**Summary box:** *What is already known on this topic:*

- Several studies have suggested that DNA methylation biomarkers could have the potential to improve prognostic accuracy for patients with colorectal cancer (CRC), but these studies mostly did not include large-scale external validation
- Many CpG sites associated with CRC prognosis and prognostic models based on these CpGs have been proposed
- An independent study to validate these biomarkers and prediction models is essential for assessing their utility in clinical practice, but has not yet performed

*What this study adds:*

- This external validation study verified the prognostic relevance of a fraction of existing DNA methylation-based prognostic biomarkers for CRC
- Published CpG-based prognostic models all performed poorly in our external validation and were rated as at high risk of bias, so they do not seem to be useful for clinical practice

## INTRODUCTION

Colorectal cancer (CRC) is the third most common cancer and a leading cause of cancer-related death worldwide.^1 2^ Globally, there were 1.93 million new cases of CRC and 0.94 million CRC-related deaths in 2020, and the numbers are projected to continue to rise in the future.^2^

Aberrant DNA methylation is an important hallmark of CRC development, and DNA methylation biomarkers have shown promise for CRC prognostication.^3^ DNA methylation is one of the most common epigenetic modifications regulating gene expression.^4^ This process involves the addition of a methyl group to the C5 position of the cytosine nucleotide, located next to a guanine nucleotide (i.e., CpG dinucleotide, Figure 1A).^4^ Due to the stable nature of DNA as compared with RNA and protein, testing DNA methylation biomarkers from collected specimens can be easily realized in clinical settings.^5^ Meanwhile, the accessibility of genome-wide epigenetic technologies enables high-throughput analysis of aberrant DNA methylation in tumor tissue.^6^ Accumulating evidence supports the prognostic relevance of various DNA methylation biomarkers in patients with CRC,^3^ and several prognostic models based on prognostically-relevant CpGs have been developed.^7-15^ However, so far, none of these potential prognostic markers or models have been translated to clinical practice, hampered by a lack of validation analyses in large independent patient cohorts,^3^ which is a huge problem in cancer biomarker research in general. Innumerable biomarkers are published each year, but only very few ever make it to clinical practice.

**Figure 1.**
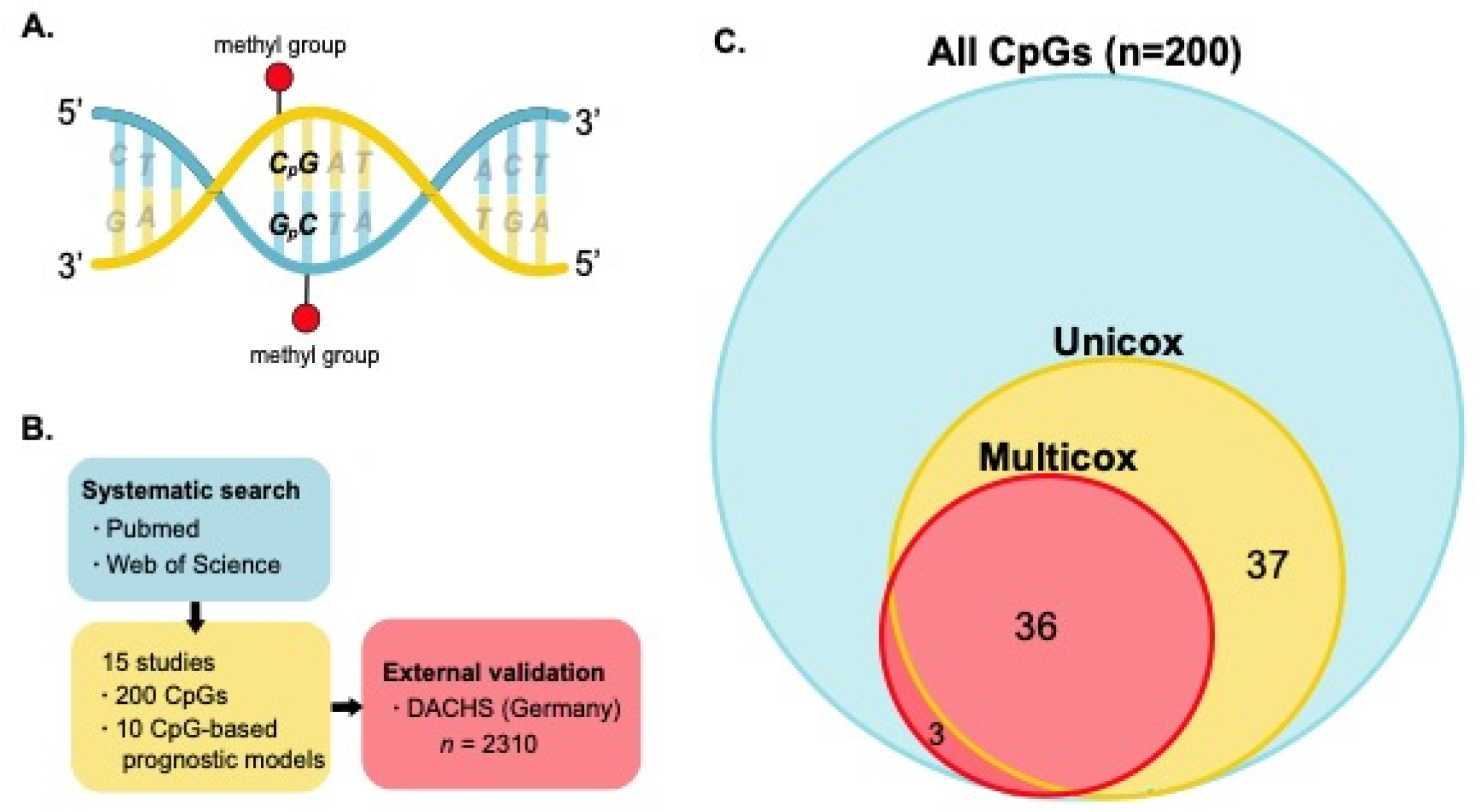
A) Schematic illustration of DNA methylation B) Workflow of the validation study C) The number of CpGs significant in Cox regression analyses. The number of CpGs to be validated was 200 in total. ‘Unicox’ represents the number of CpGs significant in univariable Cox regression analysis, and ‘Multicox’ represents the number of CpGs significant in multivariable Cox regression analyses with adjustment of age, sex, TNM stage, treatment, and cancer location. DNA icon in A) was downloaded from vecteezy.com under free license.

In this study, we aimed to externally validate previously reported prognostically relevant CpGs and CpG-based prognostic models for the survival of patients with CRC, based in a large, multicentric study cohort.

## METHODS

### Systematic review

We identified eligible studies reporting prognostic CpG sites for CRC patients through a systematic search in Pubmed and Web of Science. The complete search strategy is shown in Supplementary Table 1. We assessed the methodological quality of prognostic models based on a tool to assess the risk of bias and applicability of prediction model studies (PROBAST).^16^ A detailed description of the systematic review is provided in the Supplementary Methods.

### Validation cohort

Our external validation analysis is based on data from the DACHS study, a large population-based case-control and patient cohort study on CRC conducted in the Rhine-Neckar region in the southwest of Germany from 2003-2021. Patients in the present study were diagnosed between 2003 and 2013 and were followed up until 2020. Details of the DACHS study have been published elsewhere.^17 18^ We conducted and reported this study according to the Transparent Reporting of a multivariable prediction model for Individual Prognosis or Diagnosis statement (TRIPOD, the checklist can be found in Supplementary Table 2).^19^

Patients who had a first diagnosis of primary CRC, were above 30 years old, and were physically and mentally capable of participating in a one-hour interview, were recruited from a total of 22 hospitals in the study region. At baseline, patients provided extensive information during a face-to-face interview, carried out by trained interviewers using a standardized questionnaire. Genome-wide methylation analysis was performed on tissue DNA from formalin-fixed paraffin-embedded tumor blocks using the Illumina Human Methylation 450 Bead-Chip (Illumina, San Diego, CA, USA), which interrogated over 485000 CpG sites.^9^ Patients were followed at 3, 5, and 10 years after diagnosis, respectively. Details of data collection are provided in the Supplementary Methods.

For this validation study, we only included patients who had available genome-wide DNA methylation information and at least one follow-up record. Additionally, we excluded overlapping patients when validating one study that had been based on patients from the DACHS cohort to ensure independent validation. The sample size of the validation study was determined by all available data on the DACHS cohort during the study period.

### Statistical analyses

All statistical analyses were performed using R version 4.0.0; the R code can be found at https://github.com/TanweiY/CpGs-validation. The R package ‘CHAMP’ was used to read and preprocess raw DNA methylation data files generated by the iScan array scanner.^20^ The methylation degree of each CpG site was converted into β values, ranging from 0 (completely unmethylated) to 1 (fully methylated). For presenting patient baseline characteristics, absolute and relative frequencies are shown for categorical variables and the median with range for continuous variables. For dealing with missing information in patient characteristics, multiple imputation was performed with the R package ‘MICE’^21^ to generate m=20 imputed datasets. Details regarding DNA methylation pre-processing and the imputation procedure can be found in the Supplementary Methods.

The primary outcome of our validation was overall survival (OS), because all the included studies found relevant CpGs using the outcome of OS. The survival time for OS was calculated as the time between the date of CRC diagnosis and the date of death from any cause or censoring. Our secondary outcome was disease-free survival (DFS), and its survival time was measured as the date of diagnosis to the date of recurrence (i.e., reappearance of CRC, metastases, or death from CRC) or censoring. Median follow-up time was computed using the reverse Kaplan-Meier method.^19^ We used a cumulative incidence function within a competing risks framework to calculate the cumulative incidence of recurrence,^22^ with death from other causes considered a competing event.

For the validation of individual CpGs and prognostic models, we used Cox proportional hazard models to assess unadjusted and adjusted hazard ratios (HRs) with 95% confidence intervals (CIs) for OS, and cause-specific Cox proportional hazard models for DFS. Covariables adjusted for in the multivariable Cox regression models included age, gender, treatment, TNM stage, and tumor location. In sensitivity analyses, microsatellite instability (MSI) was additionally included in the adjusted models. For each CpG to validate, the original β value was converted to the *Z* score for inclusion into the Cox regression models. The prognostic scores for each patient were calculated based on the equations of prognostic models provided by the identified studies and were categorized into quartiles before entering Cox regression models.

For prognostic models that were found to be significant in the adjusted models, we plotted dose-response curves to illustrate the association between each prognostic score and the risk of all-cause death by restricted cubic splines adjusted for age, gender, treatment, TNM stage, and tumor location. We quantified the discriminative power for all prognostic models using the time-dependent AUC at 1, 3, 5, and 8 years after CRC diagnosis using the R package ‘riskRegression’.^23^ AUCs were calculated using both the original continuous score values and quartiles of score values. We additionally performed stratified sensitivity analyses for AUCs of prognostic models by patient characteristics. The calibration performance of models was measured with calibration plots in which patients were evenly divided into ten groups and the predicted survival probability (x-axis) by prognostic models was compared against the observed survival probability (y-axis) at 2, 5, and 10 years after CRC diagnosis respectively. A 45° diagonal line represents perfect calibration, while deviation below or above this line implies overestimated or underestimated survival. Since none of the included studies provided information on baseline survival for the model, we used the recalibrated baseline survival.

In order to evaluate the added value of prognostic models, we compared the Cox models including clinical variables only (age, sex, and TNM stage) and the models including both clinical variables and prognostic scores using corresponding likelihood ratio tests.^24^ Besides, we also calculated the differences in AUCs between the two models.

## RESULTS

### Characteristics of studies to be validated

The flowchart for the selection of studies is shown in Supplementary Figure 1. We contacted the corresponding authors for a total of four studies^25-28^ to request necessary information (i.e., the list of prognostically relevant CpGs, equations for CpG-based prognostic models) twice, but none of them provided the information we needed. Therefore, 15 studies^7-15 26 28-32^ were finally included in the validation study (Figure 1B). Characteristics of included studies are shown in Supplementary Table 3. Included studies were published between 2017 and 2022. All, except for one,^9^ used data from the Cancer Genome Atlas Program (TCGA). Four studies only reported individual prognostically-associated CpGs,^13 29 31 32^ and 11 studies additionally constructed CpG-based prognostic models, two of which^26 28^ did not provide an equation to calculate the prognostic scores.

### Characteristics of the validation cohort

Of the patients included in the DACHS study between 2003 and 2013, a genome-wide methylation array was performed for 2316 patients. All of the 200 prognostically-associated CpGs extracted from included studies were contained in our genome-wide methylation data after pre-processing, without missing values. After excluding patients without follow-up information, 2310 patients were included in the primary validation analyses. For the validation of the study by Gündert et al, which was based on data from the DACHS study,^9^ we excluded 572 overlapping patients to ensure independent validation.

The patients’ outcome characteristics are summarized in Supplementary Table 4. The median follow-up time for the 2310 participants was 10.4 years (interquartile range [IQR] 10.4-12.4). A total of 1252 (54%) participants died from any cause during the follow-up period, with cumulative mortality rates at three, five, and ten years after CRC diagnosis being 21%, 32%, and 51%, respectively. The proportions of missing values for the patients’ characteristics were all below 14% (Table 1). The cumulative incidence curves of the patients overall and those of patients without any missing values for baseline characteristics were very close (Supplementary Figure 2). Compared with the original derivation cohorts, our validation cohort had much larger sample size (2310 vs 127-572), an older study population at CRC diagnosis (median: 69 vs 64-69), a slightly higher proportion of patients at TNM stage III/IV (48% vs 17%-47%), and a higher overall death rate (54% vs 14%-24%).

**Table 1.** Characteristics of the validation cohort.

| **Variable** | **Validation cohort**<br>**N = 2310** | **Missing<sup>1</sup>** |
| --- | --- | --- |
| Diagnosis year |  |  |
| Range | 2003-2013 | 0 |
| Median | 2007 |  |
| Age at diagnosis |  | 0 |
| Range | 30-96 |  |
| Median | 69 |  |
| Gender |  | 0 |
| Female | 964 (42%) |  |
| Male | 1346 (58%) |  |
| Stage at diagnosis |  | 42 |
| Stage I | 397 (18%) |  |
| Stage II | 788 (35%) |  |
| Stage III | 747 (33%) |  |
| Stage IV | 336 (15%) |  |
| CRC location |  | 1 |
| Distal colon | 636 (28%) |  |
| Proximal colon | 851 (37%) |  |
| Rectum | 822 (36%) |  |
| Treatment |  | 6 |
| Yes | 1087 (47%) |  |
| No | 1217 (53%) |  |
| Microsatellite instability |  | 318 |
| Yes | 223 (11%) |  |
| No | 1769 (89%) |  |
| BRAF mutation |  | 230 |
| Yes | 166 (8%) |  |
| No | 1914 (92%) |  |
| KRAS mutation |  | 229 |
| Yes | 693 (33%) |  |
| No | 1388 (67%) |  |
| CIMP phenotype |  | 111 |
| High | 388 (18%) |  |
| Low | 1811 (82%) |  |

<sup>1</sup>Missing values were imputed by multiple imputation for 20 times. CRC = colorectal cancer.

### Validation of prognostically relevant CpGs

None of the 200 extracted CpGs was reported by more than one study. Among the 200 unique CpGs, 73 (37%) were significantly associated with OS in univariable Cox regression analysis, and only 39 (20%) were significant in multivariable Cox regression adjusting for clinical variables (Figure 1C). Table 2 shows the results for the multivariable Cox regression. All, except for three CpGs, which were significantly associated with OS in our validation dataset when adjusting for clinical variables, had the same direction of association as those reported in the original studies. For the secondary outcome of DFS, results were largely similar, though slightly more CpGs (55, 27%) were significantly associated with DFS when adjusting for clinical variables. Results were also similar when additionally adjusting for MSI status. Details for the validation of all CpGs can be found in Supplementary Table 5.

**Table 2.** Confirmed CpGs at *P* value < 0.1 in multivariable Cox regression and direction of the hazard ratios compared to the original studies.^1^.

| CpG | Endpoint in development study | Validation results |  |  |
| --- | --- | --- | --- | --- |
| | | $P$ value<sup>1</sup> | aHR(95%CI)<sup>2</sup> | Direction<sup>3</sup> |
| cg05165940 | OS | <0.001 | 1.13 (1.07, 1.19) | Same↑ |
| cg05646575 | OS and DSS<sup>4</sup> | <0.001 | 0.85 (0.80, 0.90) | Same↓ |
| cg08804626 | OS and DSS<sup>4</sup> | <0.001 | 0.86 (0.81, 0.92) | Same↓ |
| cg12510999 | OS and DSS<sup>4</sup> | <0.001 | 0.88 (0.82, 0.94) | Same↓ |
| cg18195165 | OS and DSS<sup>4</sup> | <0.001 | 0.89 (0.83, 0.94) | Same↓ |
| cg19184885 | OS and DSS<sup>4</sup> | <0.001 | 0.89 (0.83, 0.95) | Same↓ |
| cg21770617 | OS and DFS<sup>4</sup> | <0.001 | 1.11 (1.05, 1.18) | Same↑ |
| cg23750514 | OS and DSS<sup>4</sup> | <0.001 | 0.83 (0.78, 0.88) | Same↓ |
| cg00660989 | OS | 0.001 | 0.91 (0.86, 0.96) | Same↓ |
| cg11056055 | OS and DSS<sup>4</sup> | 0.001 | 0.90 (0.84, 0.95) | Same↓ |
| cg14270346 | OS and DSS<sup>4</sup> | 0.001 | 0.90 (0.84, 0.96) | Same↓ |
| cg21096966 | OS | 0.001 | 0.91 (0.86, 0.96) | Same↓ |
| cg27525037 | OS | 0.001 | 0.91 (0.85, 0.96) | NR |
| cg01131395 | OS and DSS<sup>4</sup> | 0.002 | 0.90 (0.85, 0.96) | Same↓ |
| cg08729279 | OS and DSS<sup>4</sup> | 0.002 | 0.90 (0.85, 0.96) | Same↓ |
| cg18736676 | OS and DSS<sup>4</sup> | 0.002 | 0.91 (0.86, 0.97) | Same↓ |
| cg23285774 | OS | 0.002 | 0.92 (0.87, 0.97) | Same↓ |
| cg08021727 | OS | 0.003 | 1.10 (1.03, 1.16) | Same↑ |
| cg16399624 | OS and DSS<sup>4</sup> | 0.003 | 0.91 (0.86, 0.97) | Same↓ |
| cg19340296 | OS and DSS<sup>4</sup> | 0.003 | 0.90 (0.85, 0.97) | Same↓ |
| cg05194618 | OS | 0.004 | 0.92 (0.87, 0.97) | Same↓ |
| cg24674703 | OS | 0.004 | 1.09 (1.03, 1.15) | Opposite |
| cg08617020 | OS and DSS<sup>4</sup> | 0.007 | 0.92 (0.86, 0.98) | Same↓ |
| cg12751565 | OS and DFS<sup>4</sup> | 0.007 | 1.08 (1.02, 1.15) | Same↑ |
| cg04138846 | OS | 0.009 | 1.09 (1.02, 1.16) | Same↑ |
| cg06328127 | OS | 0.009 | 1.08 (1.02, 1.14) | Same↑ |
| cg00147009 | OS | 0.013 | 0.93 (0.87, 0.98) | Same↓ |
| cg16336556 | OS and DSS<sup>4</sup> | 0.013 | 0.92 (0.86, 0.98) | Same↓ |
| cg05821816 | OS | 0.015 | 0.93 (0.87, 0.99) | Same↓ |
| cg14175690 | OS | 0.016 | 1.08 (1.01, 1.15) | NR |
| cg15786837 | OS | 0.018 | 1.07 (1.01, 1.13) | Opposite |
| cg25074185 | OS | 0.022 | 0.93 (0.87, 0.99) | NR |
| cg24888257 | OS | 0.025 | 0.94 (0.89, 0.99) | Same↓ |
| cg00110724 | OS | 0.026 | 1.07 (1.01, 1.15) | Same↑ |
| cg01340952 | OS | 0.026 | 0.94 (0.88, 0.99) | Same↓ |
| cg03977782 | OS | 0.026 | 0.93 (0.88, 0.99) | Opposite |
| cg03017653 | OS | 0.032 | 1.07 (1.01, 1.13) | Same↑ |
| cg06952671 | OS | 0.036 | 1.06 (1.00, 1.13) | NR |
| cg06884352 | OS and PFS<sup>4</sup> | 0.047 | 1.06 (1.00, 1.12) | Same↑ |
|  |  | *P* value<sup>1</sup> | aHR(95%CI)<sup>2</sup> | Direction<sup>3</sup> |
| cg14983135 | OS and DSS<sup>4</sup> | 0.052 | 0.94 (0.88, 1.00) | Same↓ |
| cg07936950 | OS | 0.06 | 1.06 (1.00, 1.13) | Same↑ |
| cg20428713 | OS | 0.061 | 1.06 (1.00, 1.12) | Same↑ |
| cg04992638 | OS | 0.068 | 1.05 (1.00, 1.12) | NR |
| cg10758824 | OS and DSS<sup>4</sup> | 0.068 | 0.94 (0.88, 1.00) | Same↓ |
| cg02654360 | OS | 0.069 | 0.95 (0.89, 1.00) | NR |
| cg02760766 | OS | 0.069 | 1.05 (1.00, 1.10) | NR |
| cg13066016 | OS | 0.081 | 1.05 (0.99, 1.12) | Same↑ |
| cg02245020 | OS | 0.091 | 0.95 (0.90, 1.01) | NR |
| cg15105326 | OS | 0.092 | 0.95 (0.90, 1.01) | Opposite |

NR = Not reported; OS = Overall survival; DSS = Disease-specific survival; DFS = Disease-free survival; PFS = Progression-free survival
<sup>1</sup> Only CpGs with *P* values < 0.1 for the main outcomes (i.e., OS) of original discovery studies were listed on the table.
<sup>2</sup> Multivariable Cox regression analyses with adjustment of age, sex, TNM stage, treatment, and cancer location.
<sup>3</sup> Compare the direction of association reported in the original study and that in this validation study; ↑ represents higher methylation level at the CpG site is associated with higher mortality risk, while ↓ represents lower methylation level of the CpG site is associated with lower mortality risk.
<sup>4</sup> OS was the main outcome used to find relevant CpGs in the original discovery study.
The original methylation level ( $\beta$ value) was converted to the *Z* score; HRs refer to the relative risk of increasing methylation levels per one standard deviation from the mean.

### Validation of CpG-based prognostic models

We identified a total of ten CpG-based prognostic models derived from nine studies.^7-12 14 28 30^ The equation to calculate prognostic score, the distribution of each prognostic score in our validation cohort, and the performance of these models reported by original development studies are shown in Supplementary Table 6. Five prognostic models from four studies^8-10 14^ were independently associated with OS (Supplementary Table 7). The relationships between the prognostic scores and survival were largely linear (Figure 2A). An increasing trend of relative risk from the lowest (Q1) to the highest (Q4) quartile was observed for four out of the five prognostic models, but the relative risks for Q4 compared with Q1 were all below 2.0 (Figure 2B).

**Figure 2.**
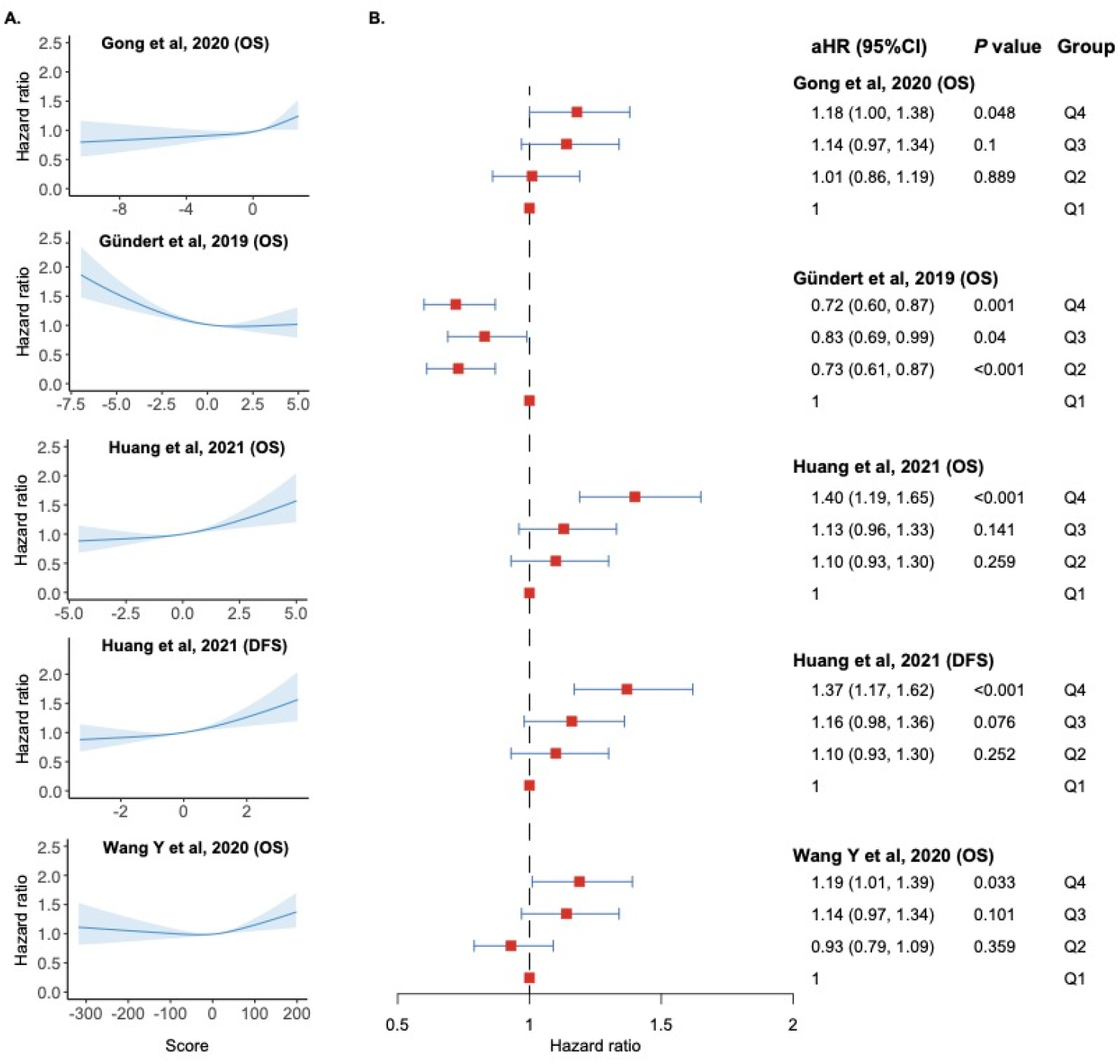
A) Dose-response association of prognostic scores and survival in validation analyses based on data from the DACHS study. Restricted cubic splines analyses with adjustment for age, sex, tumor location, TNM stage, and treatment B) Relative risk by quartiles of prognostic scores in the validation cohort. aHR = adjusted hazards ratio; CI = Confidence interval; OS = Overall survival; DFS = Disease-free survival.

The time-dependent AUCs generated by using the continuous prognostic score and by using quartiles of the score were almost the same and were all low (nearly all <0.6) (Supplementary Table 8). For the five prognostic models independently associated with survival (Figure 3A), the highest AUCs were observed for the model developed by Gu□ndert et al,^9^ which was also derived from data of the DACHS study, with time-dependent AUCs ranging from 0.59 at 8 years to 0.64 at 1 year. The AUCs for other prognostic models were all around 0.55. The poor AUCs persisted in all the stratified analyses by age, gender, TNM stage, tumor location, treatment, MSI status, BRAF mutation, KRAS mutation, and CIMP phenotype (Supplementary Table 9). The AUCs in our validation study were in contrast to the optimistic AUCs reported by the original studies. Similarly, these models showed very poor calibration accuracy at 2, 5, and 10 years after CRC diagnosis (Supplementary Figure 3).

**Figure 3.**
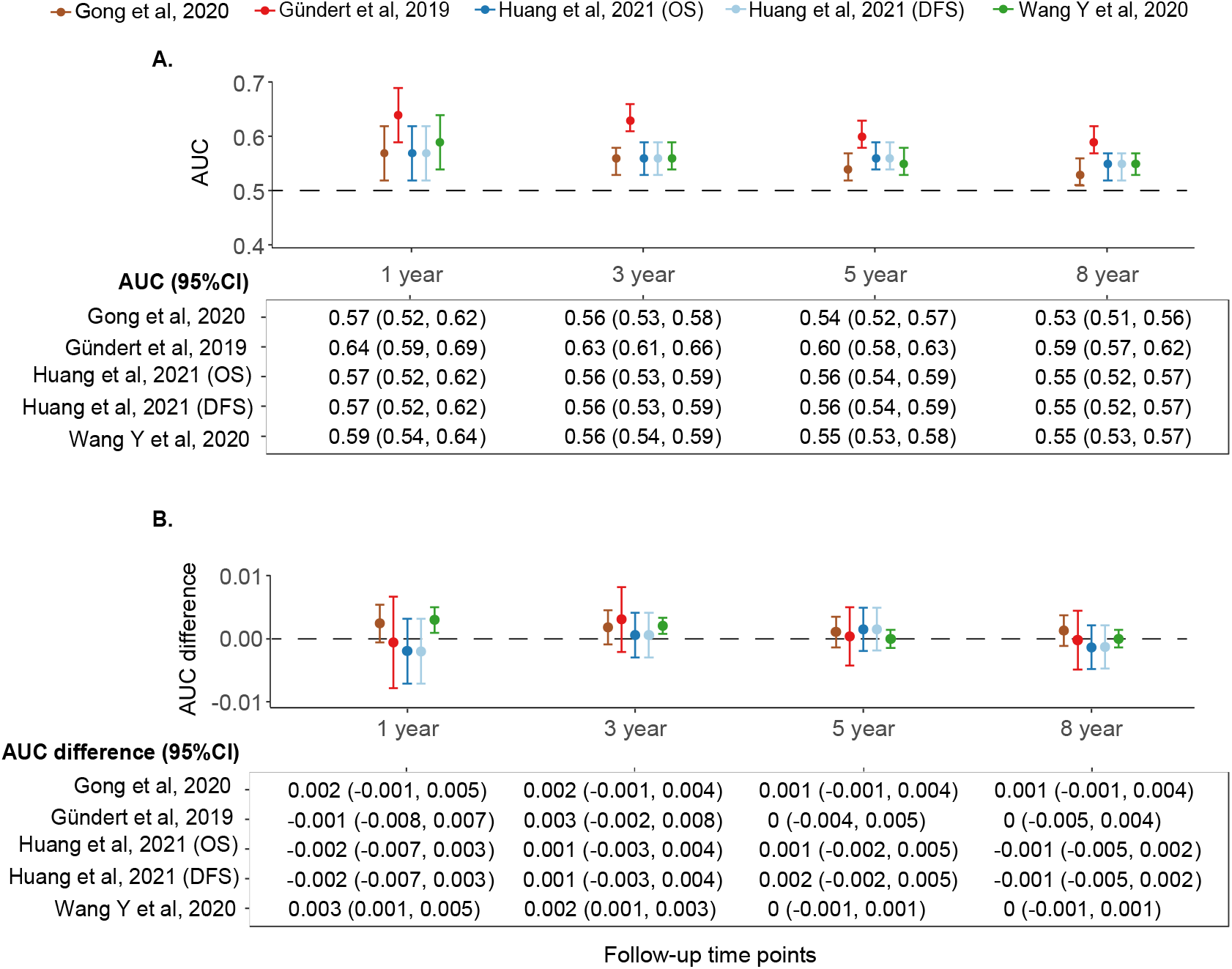
A) Overall and time-dependent AUC for prognostic scores significant in multivariable Cox regression B) Differences in overall and time-dependent AUC comparing models based on clinical variables only and those with added prognostic scores. AUC = Area under the receiver operating characteristic curve; CI = Confidence interval; OS = Overall survival; DFS = Disease-free survival.

Regarding the added value of the five prognostic models, four of them could improve the goodness-of-fit of the Cox model based on age, sex, and TNM stage when added to the model (Supplementary Table 10). Nevertheless, they could only improve the time-dependent AUCs by at most 0.003 when comparing the two models (Figure 3B). Marginal improvement in AUCs was also observed for the secondary outcome of DFS (Supplementary Table 11).

### Methodological quality of studies on CpG-based prognostic models

According to PROBAST, the nine model development studies were all rated as at high risk of bias (ROB). Figure 4 and Supplementary Table 12 provide details for the assessment of each model. More than half of the studies had a high ROB for the participant domain, which means patients used to develop the model might not be representative of the model’s targeted populations. The other studies did not report patients’ inclusion or exclusion criteria.

**Figure 4.**
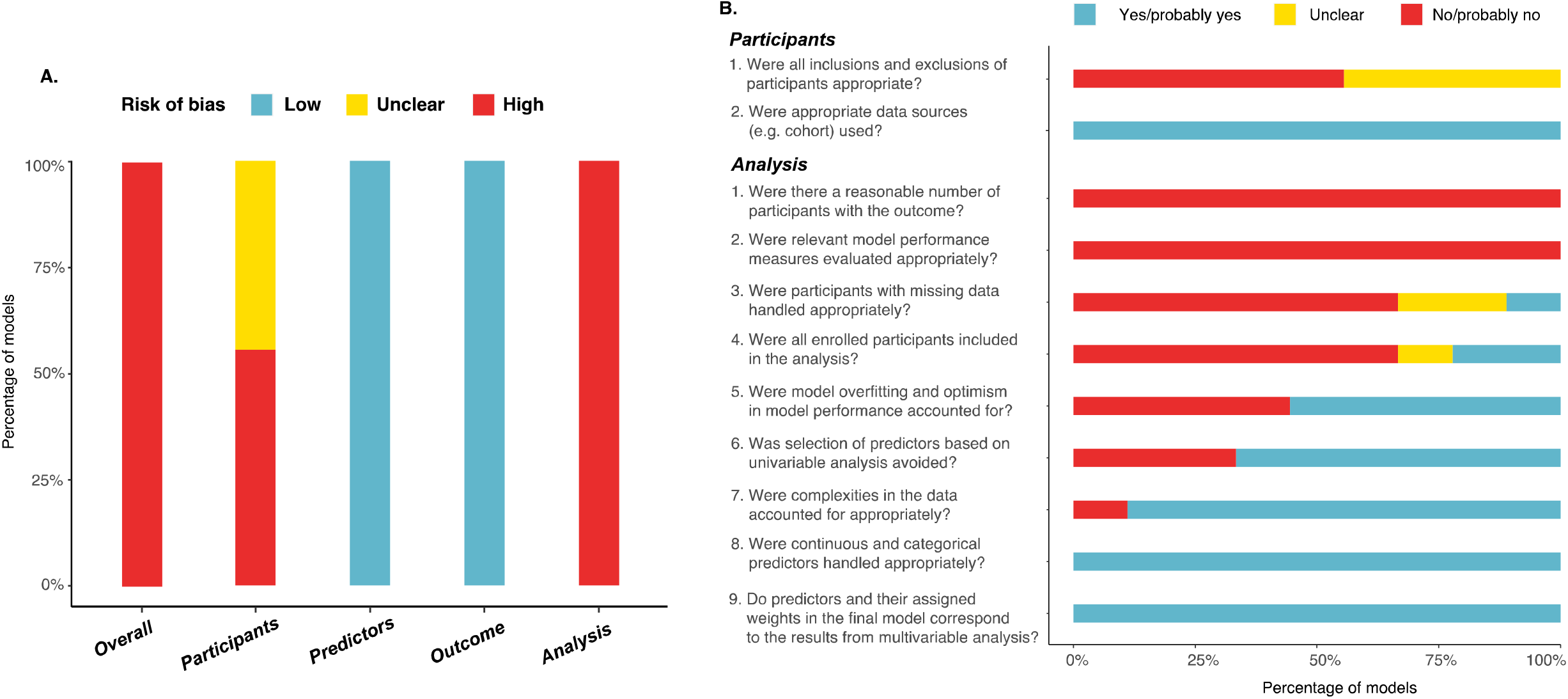
PROBAST (prediction model risk of bias assessment tool) risk of bias for all model development studies (n=9, A) and broken down for two domains with high risk of bias (B) The risk of bias for the predictors domain and outcome domain were low for all models, because these models used the same type of predictors (i.e., DNA methylation markers) and outcome (i.e., overall survival), and nearly all of them were derived from the TCGA database, in which the definition and assessment of predictors and outcomes were standardized.

Notably, all studies were at high ROB for the analysis domain. The main reason was insufficient sample size in comparison with the genome-wide methylation information investigated. None of the models were evaluated for calibration. Only one study handled missing values properly by multiple imputations,^9^ while the remaining studies either excluded patients with missing values or did not report this information. Four out of nine studies (44%) did not take measures to account for model overfitting,^7 8 10 14^ and three of the nine studies selected CpGs simply based on the *p*-values in univariable and multivariable analyses without considering established clinical knowledge.^8 14 28^

## DISCUSSION

We performed the first comprehensive external validation of the so far published 200 prognostically relevant CpG sites and 10 CpG-based prognostic models for CRC in a large multicentric patient cohort. Only 39 CpGs (20%) and five prognostic models (50%) were associated with OS when adjusting for clinical variables. The model performance for the five prognostic models was poor in both discrimination and calibration, and they all showed a negligible added prognostic value when used in conjunction with clinical variables. Based on the framework of PROBAST, all of the model development studies were rated as high risk of bias.

We found that none of the CpGs identified in the original studies that were associated with CRC survival was overlapping between studies, and only small fraction (20%) could be successfully validated in our external patient cohort. The main reason might be the extreme multicollinearity nature of the high dimensional DNA methylation data.^33^ The number of CpGs investigated using a genome-wide approach is enormous and many of them are highly correlated. Commonly used feature selection methods for regression models (e.g., LASSO) can select only one CpG that is highly correlated with other CpGs, which are then excluded from the model.^33^ Consequently, studies using different patient cohorts and different feature selection strategies are likely to identify different sets of CpGs with prognostic values for CRC.^33^ Due to the same reason, it is likely that some of the CpGs that failed to be validated in our study might still hold prognostic value to some extent, as they might be correlated with the CpGs that truly are predictive of survival.

In our external validation, only half of the 10 identified CpG-based prognostic models were independently associated with survival, all of which showed poor model performance. This finding is in stark contrast to the moderate to excellent performance reported in the original studies. Only three of the original studies performed external validation,^9 10 30^ and model performance was also lower in the validation cohorts. Several factors may contribute to the reduced performance in our external validation. The first common contributor is the difference between our validation cohort and original model development cohorts in terms of methodological and patient characteristics.^19^ Indeed, the best discriminative performance was observed for the prognostic model developed in a similar but non-overlapping patient population from the DACHS study. Nevertheless, the highest AUC was still below 0.65, and the poor discrimination was consistent in our sensitivity analyses stratified by patient characteristics. Moreover, all models had poor calibration accuracy even using the recalibrated baseline survival. Second, the high multicollinearity nature of high dimensional data may also partly explain the poor performance of sparse CpG-based prognostic models.

Finally, all models were prone to a high risk of bias according to the PROBAST assessment,^16^ especially in the domain of statistical analysis, which might explain the poor model performance in external validation. Specifically, the available sample sizes and the number of death events were naturally limited compared with the enormous number of CpGs investigated, increasing the risk of overfitting the model.^19^ The small sample size can also increase the instability of the model.^34^ Still, nearly half of the studies did not take any measure, such as resampling techniques, to control for these issues. Also, missing data were rarely properly handled, and model calibration was never evaluated. These methodological issues in model development could have resulted in unreliable models, which leads to low generalizability.

Our results showed that the five prognostic models added no additional prognostic value to commonly used clinical variables like age, sex, and TNM stage. Of note, six included studies first constructed a Cox prognostic model based on individual CpGs to create a methylation score, which was subsequently used as a single predictor, together with other clinical variables, to construct a combined Cox prognostic model.^7 9 10 12 28 30^ Although the reported performance for these combined prognostic models were excellent, most of these models were evaluated in the model development dataset only.^7 10 12 28 30^ We were unable to externally validate the five prognostic models based on a combination of clinical variables and CpG-based prognostic score, since none of the studies provided sufficiently detailed information to calculate the model.^7 9 10 12 28 30^

A key strength of this validation study is the use of a large patient cohort with very few missing data. Besides, in order to provide a comprehensive appraisal of prognostic models, we combined the quantitative external validation with the qualitative assessment of the risk of bias. Nonetheless, this study has some limitations. First, we were unable to validate two of the CpG-based prognostic models due to a lack of relevant information to calculate the model^26 28^, although efforts have been made to request the missing information. Second, our validation cohort and the development cohort of one prognostic model^9^ were both derived from the same patient cohort, though we have excluded overlapping patients when validating this model. Third, our systematic search only focused on epigenome-wide studies. Studies using a candidate gene approach might also identify prognostically meaningful CpGs and potentially useful CpG-based models. Lastly, this study was based on CRC patients in Germany only, with potentially limited generalizability.

In view of the poor validation results obtained by this study, well-designed studies are needed to further clarify the prognostic role of DNA methylation biomarkers in CRC. Such studies should ideally focus on patients with intermediate-stage CRC, have a large sample size, use methods capable of analyzing high-dimensional-omics data, and have an external dataset to validate their results. Most importantly, relevant methodological and reporting guidelines should be carefully followed throughout the research. For machine learning-based prognostic models, model interpretability should be considered, and an automatic system for computation should be developed for real-world application.

In conclusion, our study verified the prognostic relevance of a fraction of DNA methylation biomarkers for CRC reported by previous epigenome-wide studies. Published CpG-based prognostic models all performed poorly in our external validation and were rated as at high risk of bias. None of these models seemed useful for application in clinical practice. Additional work is needed to investigate meaningful DNA methylation biomarkers and develop relevant prognostic models, using methods capable of dealing with high dimensional methylation data and following existing methodological guidance.

## Data Availability

All data produced in the present study are available upon reasonable request to the authors

## Acknowledgement

The authors would like to thank the study participants, the interviewers who collected the data, and the medical documentalists who processed the data and follow-up information. The authors appreciate cooperating hospitals and pathology institutes that recruited patients for this study, provided tumor tissue samples or performed pathology analyses.

## Contributors

MH and TY had the idea for the study, and designed the protocol. TY conducted and reported this work under the continuous supervision of MH. DE gave inputs for data analysis and discussion. KET contributes to the design of figures. KET, WR, BHM, AB, MK, HB, and BB contributed to the acquisition of data. MH, DE, KET, ZF, WR, BHM, AB, MK, HB, BB, and HB critically revised the report. MH is the guarantor of this work and accepts full responsibility for the work, had access to the data, and controlled the decision to publish. The corresponding author attests that all listed authors meet authorship criteria and that no others meeting the criteria have been omitted.

## Ethical approval

The study was approved by the ethics committees of the Medical Faculty of the University of Heidelberg and of the Medical Chambers of Baden-Württemberg and Rhineland-Palatinate.

## Data sharing

The datasets used and analyzed during the current study are available from the corresponding author on reasonable request.

## References

1. Dekker E, Tanis PJ, Vleugels JLA, et al. Colorectal cancer. Lancet 2019;394(10207):1467–80.

2. Xi Y, Xu P. Global colorectal cancer burden in 2020 and projections to 2040. Transl Oncol 2021;14(10):101174.

3. Jung GA-OX, Hernández-Illán E, Moreira L, et al. Epigenetics of colorectal cancer: biomarker and therapeutic potential. Nat Rev Gastroenterol Hepatol 2020;17(2):111–30.

4. Egger G, Liang G Fau-Aparicio A, Aparicio A Fau-Jones PA, et al. Epigenetics in human disease and prospects for epigenetic therapy. Nature 2004;429(6990):457–63.

5. Hashimoto Y, Zumwalt TJ, Goel A. DNA methylation patterns as noninvasive biomarkers and targets of epigenetic therapies in colorectal cancer. Epigenomics 2016;8(5):685–703.

6. Yong WS, Hsu FM, Chen PY. Profiling genome-wide DNA methylation. Epigenetics Chromatin 2016;2016(9):26.

7. Chen F, Pei LJ, Liu SY, et al. Identification of a Novel Immune-Related CpG Methylation Signature to Predict Prognosis in Stage II/III Colorectal Cancer. Front Genet 2021;12:684349. doi: 10.3389/fgene.2021.684349

8. Gong S, Ye W, Liu T, et al. The Development of Three-DNA Methylation Signature as a Novel Prognostic Biomarker in Patients with Colorectal Cancer. Biomed Res Int 2020;2020:3497810. doi: 10.1155/2020/3497810 [published Online First: 2020/12/10]

9. Gündert M, Edelmann D, Benner A, et al. Genome-wide DNA methylation analysis reveals a prognostic classifier for non-metastatic colorectal cancer (ProMCol classifier). Gut 2019;68(1):101–10. doi: 10.1136/gutjnl-2017-314711 [published Online First: 2017/11/05]

10. Huang H, Zhang L, Fu J, et al. Development and validation of 3-CpG methylation prognostic signature based on different survival indicators for colorectal cancer. Mol Carcinog 2021;60(6):403–12. doi: 10.1002/mc.23300 [published Online First: 2021/04/08]

11. Li DH, D. XH, Liu M, et al. A 10-gene-methylation-based signature for prognosis prediction of colorectal cancer. Cancer Genet 2021;252-253:80–86. doi: 10.1016/j.cancergen.2020.12.009 [published Online First: 2021/01/15]

12. Wang X, Wang D, Liu J, et al. A novel CpG-methylation-based nomogram predicts survival in colorectal cancer. Epigenetics 2020;15(11):1213–27. doi: 10.1080/15592294.2020.1762368 [published Online First: 2020/05/13]

13. Wang XY, Zhang DS, Zhang C, et al. Identification of epigenetic methylation-driven signature and risk loci associated with survival for colon cancer. Ann Transl Med 2020;8(6):324. doi: 10.21037/atm.2020.02.94

14. Wang Y, Zhang M, Hu X, et al. Colon cancer-specific diagnostic and prognostic biomarkers based on genome-wide abnormal DNA methylation. Aging (Albany NY) 2020;12(22):22626–55. doi: 10.18632/aging.103874 [published Online First: 2020/11/18]

15. Yang CS, Zhang Y, Xu XQ, et al. Molecular subtypes based on DNA methylation predict prognosis in colon adenocarcinoma patients. Aging (Albany NY) 2019;11(24):11880–92. doi: 10.18632/aging.102492

16. Moons KGM, Wolff RF, Riley RD, et al. PROBAST: A Tool to Assess Risk of Bias and Applicability of Prediction Model Studies: Explanation and Elaboration. Ann Intern Med 2019;170(1):W1–W33.

17. Brenner H, Chang-Claude J, Jansen L, et al. Reduced risk of colorectal cancer up to 10 years after screening, surveillance, or diagnostic colonoscopy. Gastroenterology 2014;146(3):709–17.

18. Hoffmeister M, Jansen L, Rudolph A, et al. Statin use and survival after colorectal cancer: the importance of comprehensive confounder adjustment. J Natl Cancer Inst 2015;107(6):djv045.

19. Moons KG, Altman DG, Reitsma JB, et al. Transparent Reporting of a multivariable prediction model for Individual Prognosis or Diagnosis (TRIPOD): explanation and elaboration. Ann Intern Med 2015;162(1):W1–73.

20. Tian Y, Morris TJ, Webster AP, et al. ChAMP: updated methylation analysis pipeline for Illumina BeadChips. Bioinformatics 2017;33(24):3982–84. doi: 10.1093/bioinformatics/btx513 [published Online First: 2017/09/30]

21. van Buuren S K. G-O. mice: Multivariate Imputation by Chained Equations in R. J Stat Softw 2011;45:1–67.

22. Zhang Z. Survival analysis in the presence of competing risks. Ann Transl Med 2017;5(3):47. doi: 10.21037/atm.2016.08.62 [published Online First: 2017/03/03]

23. Gerds TA, Mw K. Medical Risk Prediction Models: With Ties to Machine Learning (1st ed.). New York: Chapman and Hall/CRC 2021.

24. Cook NR. Quantifying the added value of new biomarkers: how and how not. Diagn Progn Res 2018;2:14. doi: 10.1186/s41512-018-0037-2 [published Online First: 2019/05/17]

25. Hao X, Luo H, Krawczyk M, et al. DNA methylation markers for diagnosis and prognosis of common cancers. Proc Natl Acad Sci U S A 2017;114(28):7414–19. doi: 10.1073/pnas.1703577114

26. Yang X, Gao L, Zhang S. Comparative pan-cancer DNA methylation analysis reveals cancer common and specific patterns. Brief Bioinform 2017;18(5):761–73. doi: 10.1093/bib/bbw063 [published Online First: 2016/07/21]

27. Deng Y, Wan H, Tian J, et al. CpG-methylation-based risk score predicts progression in colorectal cancer. Epigenomics 2020;12(7):605–15. doi: 10.2217/epi-2019-0300 [published Online First: 2020/03/18]

28. Yang H, Jin W, Liu H, et al. A novel prognostic model based on multi-omics features predicts the prognosis of colon cancer patients. Mol Genet Genomic Med 2020;8(7):e1255. doi: 10.1002/mgg3.1255 [published Online First: 2020/05/13]

29. Hou X, He X, Wang K, et al. Genome-Wide Network-Based Analysis of Colorectal Cancer Identifies Novel Prognostic Factors and an Integrative Prognostic Index. Cell Physiol Biochem 2018;49(5):1703–16. doi: 10.1159/000493614 [published Online First: 2018/09/25]

30. Xiang R, Fu T. Gastrointestinal adenocarcinoma analysis identifies promoter methylation-based cancer subtypes and signatures. Sci Rep 2020;10(1):21234. doi: 10.1038/s41598-020-78228-y [published Online First: 2020/12/06]

31. Yin ZJ, Yan XM, Wang QM, et al. Detecting Prognosis Risk Biomarkers for Colon Cancer Through Multi-Omics-Based Prognostic Analysis and Target Regulation Simulation Modeling. Front Genet 2020;11:524. doi: 10.3389/fgene.2020.00524

32. Xin JY, Wu YL, Ben S, et al. CoSMeD: a user-friendly web server to estimate 5-year survival probability of left-sided and right-sided colorectal cancer patients using molecular data. Bioinformatics 2022;38(1):278–81. doi: 10.1093/bioinformatics/btab523

33. Gareth James DWTHRT. An introduction to statistical learning : with applications in R: New York : Springer, [2013] ©2013 2013.

34. Sauerbrei W, Boulesteix Al Fau-Binder H, Binder H. Stability investigations of multivariable regression models derived from low- and high-dimensional data. J Biopharm Stat 2011;21(6):1206–31.

