## Supplement for "DNA methylation-based biomarkers and prediction models for the survival of patients with colorectal cancer: systematic review and external validation study"

SUPPLEMNET

Abbreviations used in this supplement	3

Supplementary Methods	4

Supplementary Table 1: Search strategy	6

Supplementary Figure 1: Inclusion of studies for validation	6

Supplementary Table 2. Transparent Reporting of a multivariable prediction model for Individual Prognosis or Diagnosis statement Checklist	7

Supplementary Table 4: Outcomes of validation cohort	10

Supplementary Figure 2: K-M survival curves for OS (A) and cumulative incidence curves for DFS (B) comparing the overall sample and complete cases	10

Supplementary Table 5: Cox regression analyses for individual CpGs	11

Supplementary Table 6: Equations of prognostic models, their distributions in the validation cohort, and their performance in the original development study	17

Supplementary Table 7: Cox regression analyses for prognostic scores	18

Supplementary Table 8: Time-dependent AUC for all scores	20

Supplementary Table 9: Stratified analyses for the AUC at mean follow-up time of prognostic scores	22

Supplementary Figure 3: Calibration curves for prognostic models	25

Supplementary Table 10: *P* values for the likelihood ratio test comparing models based on clinical variables only and models based on clinical variables added with prognostic scores^1^	26

Supplementary Table 11: Differences in AUC comparing models based on clinical variables only and that added with prognostic scores	27

Supplementary Table 12: PROBAST risk of bias for all prognostic models to be validated	28

### Abbreviations used in this supplement

NR = Not reported

NP = Not performed

OS = Overall Survival

DFS = Disease Free Survival

PFS = Progression Free Survival

K-M = Kaplan-Meier

ROC = Receiver operating characteristic

AUC = Area under curves

MSI = Microsatelite instability

MSS = Microsatelite stable

PROBAST = Prediction model risk of bias assessment tool

### Supplementary Methods

**Systematic review and quality assessment**

We selected eligible studies from a 2019 systematic review summarizing prognostic DNA methylation biomarkers among patients with gastrointestinal cancer.^1^ Besides, we conducted an updated systematic search in PubMed and Web of Science for studies published between July 2019 and February 2022. We used a combination of Medical Subject Headings (MESH) and key words related to colon and rectum, cancer, epigenetic signatures, and prognosis, respectively. We included studies that were peer-reviewed articles, reported in English, included patients with CRC, performed an epigenome-wide methylation array on tumor tissue samples, and reported CpGs associated with survival in CRC patients. We extracted the following study-level information onto pre-designed spreadsheets: first author, year of publication, patient characteristics (mean or median age, the percentage of females, tumor site, and the percentage of patients with tumor stage III/IV), size of training set, endpoint, event rate, and the name of prognostically relevant CpGs. For studies that additionally constructed prognostic models based on CpGs, we also extracted the equation of these prognostic models and the model performance reported in the development study. We contacted the corresponding author of the study to request information when necessary.

We additionally assessed the methodological quality of each prognostic model based a tool to assess the risk of bias and applicability of prediction model studies (PROBAST).^2^ PROBAST assesses the risk of bias (ROB) of prediction model studies in four broad domains: participants (2 signaling questions), predictors (3 signaling questions), outcome (6 signaling questions), and analysis (9 signaling questions). Each domain is rated as high (the answer to any of the signaling question in that domain is "No" or "Probably no"), low (the answer to all signaling questions is "Yes" or "Probably yes"), or unclear (relevant information is missing for some of the signaling questions, and the answer to all remaining questions is "Yes", or "Probably yes") risk of bias.^2^ The overall ROB for a prognostic model was rated as high (the model is rated as high ROB for at least one domain), low (the model is rated as low ROB on all domains), or unclear (the model is rated as unclear ROB for at least one domain and was rated as low ROB in the remaining domains).^2^ The rationale for rating each criterion was recorded for each prognostic model.

**Data collection of the DACHS cohort**

At baseline, patients provided extensive information regarding sociodemographic characteristics, lifestyle, family and medical history, and symptoms of the disease during a face-to-face interview, carried out by trained interviewers using a standardized questionnaire. Tumor characteristics and stage of disease (6th edition of the TNM staging manual) were obtained from medical records and pathology reports. Molecular tumor tissue analyses were performed based on DNA extracted from formalin-fixed, paraffin- embedded tumor samples from the patients enrolled.^3,4^ Microsatellite instability (MSI) status was determined using a mononucleotide marker panel (BAT25, BAT26, and CAT25).^5^ The presence of KRAS mutation was determined by single-stranded conformational polymorphism technique.^5^ The expression of BRAF V600E was determined independently by two experienced pathologists using immunohistochemical analyses in tissue microarray blocks.^5^ CpG Island methylator phenotype (CIMP) status was determined using a five-marker methylation panel (MLH1, MINT1, MINT2, MINT31 and MGMT), and classified based on the number of hypermethylated loci: CIMP negative (none), CIMP-low (1 or 2 loci), or CIMP-high (3 or more loci).^5^ Genome-wide methylation analysis was performed on tissue DNA using the Illumina Human Methylation 450 Bead-Chip (Illumina, San Diego, CA, USA), which interrogated over 485000 CpG sites.^3^ At scheduled follow-up visits (3, 5, and 10 years after diagnosis), standardized information on CRC therapy, comorbidities, and recurrence were obtained from the physicians of the patients. Vital status, date and cause of death were collected from the local population registries and health authorities. Details regarding data collection and follow up could be found elsewhere.^3-7^

**DNA methylation pre-processing**

We filtered failed probes based on a detection p-value threshold of 0.01, and probes with a bead count less than three. Normalization procedure was performed to correct for technical difference between the type I and type II probes, and then the ComBat method was used to correct for batch effects. Next, all the prognostically relevant CpGs reported in included studies were selected from the genome-wide methylation array.

**Multiple imputation**

We used multiple imputation to impute (20 times) missing information in all patient characteristics. The multiple imputation process generated 20 plausible datasets accounting for the uncertainty associated with missing values. All analyses, except for the sensitivity stratified analyses, were performed in the 20 imputed datasets in parallel, and results obtained from each dataset were combined based on Rubin's rule. Two imputation models were created to separately impute clinical variables and four molecular characteristics (microsatellite instability [MSI], BRAF mutation, KRAS mutation, and CIMP mutation). Variables included in the first imputation model included year of diagnosis, age, gender, TNM stage at diagnosis, CRC location, treatment, and the primary outcome (i.e., the Nelson-Aalen estimator of the baseline cumulative hazard and the outcome indicator).^8^ Predictive mean matching, logistic regression, and proportional odds model were used to impute continuous, binary, and ordered variables, respectively. For the second imputation model, 1852 CpGs related with CIMP genes based on a systematic search,^9^ age, gender, TNM stage at diagnosis, CRC location, primary outcome, MSI, BRAF mutation, KRAS mutation, and CIMP mutation were included, and the random forest method was used to impute the four molecular variables.^3^

### Supplementary Table 1: Search strategy

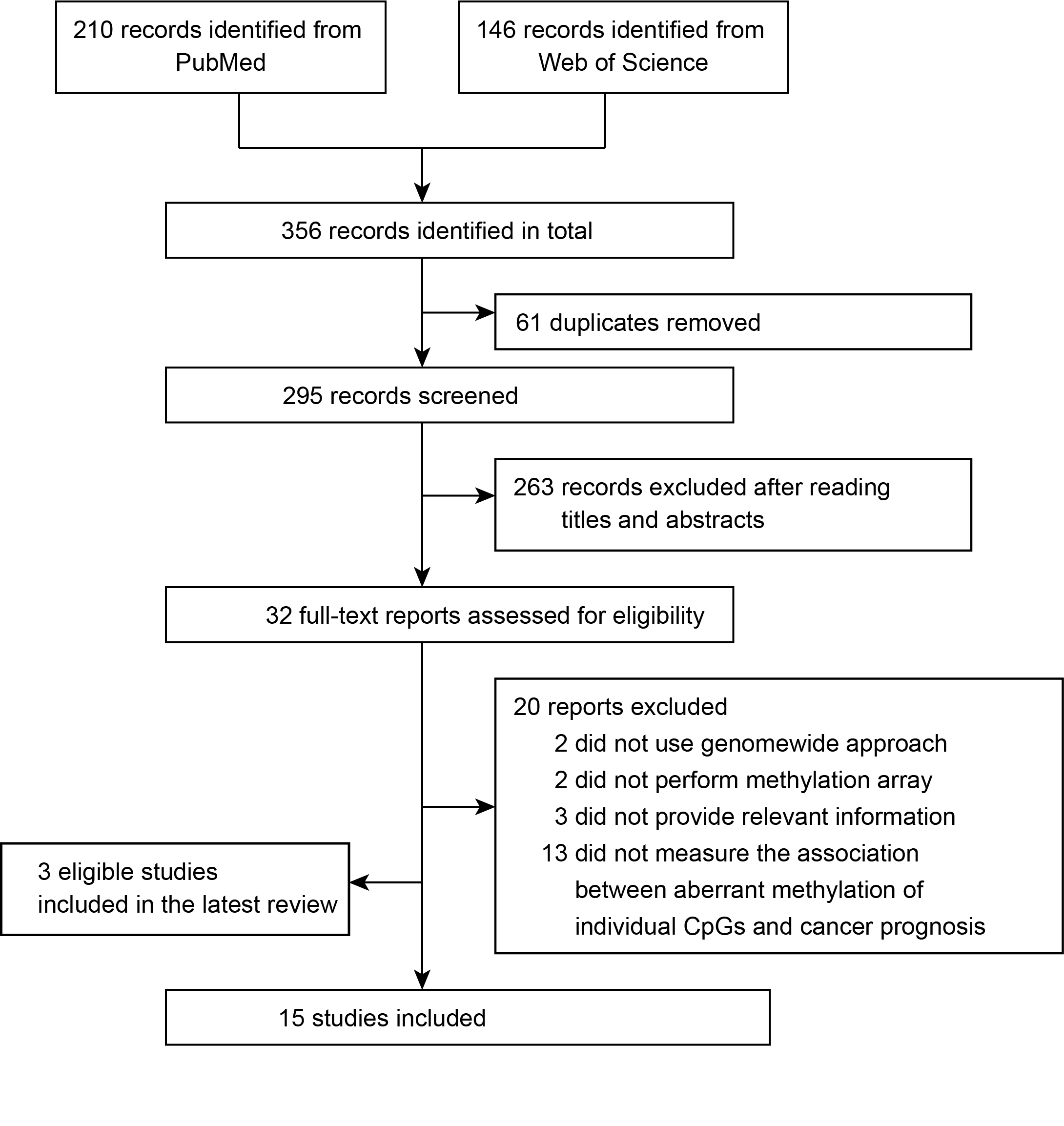

### Supplementary Figure 1: Inclusion of studies for validation

| **Keywords** |  | **Pubmed** | **Web of science^1^** |
| --- | --- | --- | --- |
| Colon and rectum | #1 | ((colon*(Title/Abstract)) OR (rect*(Title/Abstract))) OR (colorect*(Title/Abstract)) | ((AB=(colon*)) OR AB=(rect*)) OR AB=(colorect*) |
| Cancer | #2 | ((((neoplasms(MeSH Terms)) OR (cancer(Title/Abstract))) OR (tumor*(Title/Abstract))) tumour*(Title/Abstract)))OR (carcinoma*(Title/Abstract))) OR (adenocarcinoma*(Title/Abstract)) | ((((((TI=(neoplasms)) OR TI=(cancer)) OR TI=(tumor)) OR TI=(tumour)) OR TI=(carcinoma)) OR TI=(pan-cancer)) OR TI=(adenocarcinoma)) |
| Epigenetic signatures | #3 | ((((((((((methylation(MeSH Terms)) OR (dna methylation*(MeSH Terms))) OR (methylat*(Title/Abstract))) OR (epigenetic*(Title/Abstract))) OR (epigenetic marker*(Title/Abstract))) OR (DNA-methylation(Title/Abstract))) OR (hypermethylat*(Title/Abstract))) OR (hypomethylat*(Title/Abstract))) OR (CpG(Title/Abstract))) OR (epigenome-wide(Title/Abstract))) OR (methylation-based(Title/Abstract)) | ((((((((((AB=(methylation)) OR AB=(dna methylation)) OR AB=(methylat*)) OR AB=(epigenetic*)) OR AB=(epigenetic marker*)) OR AB=(DNA-methylation)) OR AB=(hypermethylat*)) OR AB=(hypomethylat*)) OR AB=(CpG)) OR AB=(epigenome-wide)) OR AB=(methylation-based) |
| Prognosis | #4 | ((((((((mortality(MeSH Terms)) OR (survival(MeSH Terms))) OR (survival analysis(MeSH Terms))) OR (mortality(Title/Abstract))) OR (survival(Title/Abstract))) OR (prognos*(Title/Abstract))) OR (predict*(Title/Abstract))) OR (outcome*(Title/Abstract))) OR (recurr*(Title/Abstract)) | ((((((TI=(mortality)) OR TI=(survival)) OR TI=(survival analysis)) OR TI=(prognos*)) OR TI=(predict*)) OR TI=(outcome*)) OR TI=(recurr*) |
| Study design | #5 | ((((database(Title/Abstract)) OR (dataset*(Title/Abstract))) OR (case-control(Title/Abstract))) OR (cohort(Title/Abstract))) OR (trial(Title/Abstract)) | ((((AB=(database)) OR AB=(dataset*)) OR AB=(case-control)) OR AB=(cohort)) OR AB=(trial) |
| Final | #5 | #1 AND #2 AND #3 AND #4 AND (humans(Filter)) AND (2019/7/1:2022/10(pdat)) AND (english(Filter))) | (#1 AND #2 AND #3 AND #4 AND #5) AND Articles (Documents Types) AND English (Languages) AND DOP =(2019-07-01 -2022-10-31) |

### Supplementary Table 2. Transparent Reporting of a multivariable prediction model for Individual Prognosis or Diagnosis statement Checklist

| **Section/Topic** | **Item** | **Checklist Item** | **Page** |
| --- | --- | --- | --- |
| **Title and abstract** | | | |
| Title | 1 | Identify the study as developing and/or validating a multivariable prediction model, the target population, and the outcome to be predicted. | p1 |
| Abstract | 2 | Provide a summary of objectives, study design, setting, participants, sample size, predictors, outcome, statistical analysis, results, and conclusions. | p3 |
| **Introduction** | | | |
| Background and objectives | 3a | Explain the medical context (including whether diagnostic or prognostic) and rationale for developing or validating the multivariable prediction model, including references to existing models. | p5 |
|  | 3b | Specify the objectives, including whether the study describes the development or validation of the model or both. | p6 |
| **Methods** | | | |
| Source of data | 4a | Describe the study design or source of data (e.g., randomized trial, cohort, or registry data), separately for the development and validation data sets, if applicable. | p6-7 |
|  | 4b | Specify the key study dates, including start of accrual; end of accrual; and, if applicable, end of follow-up. | p6-7 |
| Participants | 5a | Specify key elements of the study setting (e.g., primary care, secondary care, general population) including number and location of centres. | p7 |
|  | 5b | Describe eligibility criteria for participants. | p7 |
|  | 5c | Give details of treatments received, if relevant. | Not applicable |
| Outcome | 6a | Clearly define the outcome that is predicted by the prediction model, including how and when assessed. | Supplement, p8 |
|  | 6b | Report any actions to blind assessment of the outcome to be predicted. | Not applicable |
| Predictors | 7a | Clearly define all predictors used in developing or validating the multivariable prediction model, including how and when they were measured. | Supplement, p7 |
|  | 7b | Report any actions to blind assessment of predictors for the outcome and other predictors. | Not applicable |
| Sample size | 8 | Explain how the study size was arrived at. | p7 |
| Missing data | 9 | Describe how missing data were handled (e.g., complete-case analysis, single imputation, multiple imputation) with details of any imputation method. | Supplement, p8 |
| Statistical analysis methods | 10c | For validation, describe how the predictions were calculated. | p9 |
|  | 10d | Specify all measures used to assess model performance and, if relevant, to compare multiple models. | p9-10 |
|  | 10e | Describe any model updating (e.g., recalibration) arising from the validation, if done. | p10 |
| Risk groups | 11 | Provide details on how risk groups were created, if done. | Not applicable |
| Development vs. validation | 12 | For validation, identify any differences from the development data in setting, eligibility criteria, outcome, and predictors. | p10-11 |
| **Results** | | | |
| Participants | 13a | Describe the flow of participants through the study, including the number of participants with and without the outcome and, if applicable, a summary of the follow-up time. A diagram may be helpful. | p11 |
|  | 13b | Describe the characteristics of the participants (basic demographics, clinical features, available predictors), including the number of participants with missing data for predictors and outcome. | p11-12, table2 |
|  | 13c | For validation, show a comparison with the development data of the distribution of important variables (demographics, predictors and outcome). | p12, table1 and table2 |
| Model performance | 16 | Report performance measures (with CIs) for the prediction model. | p13-14, figure3 |
| Model-updating | 17 | If done, report the results from any model updating (i.e., model specification, model performance). | Not performed |
| **Discussion** | | | |
| Limitations | 18 | Discuss any limitations of the study (such as nonrepresentative sample, few events per predictor, missing data). | p19 |
| Interpretation | 19a | For validation, discuss the results with reference to performance in the development data, and any other validation data. | p17 |
|  | 19b | Give an overall interpretation of the results, considering objectives, limitations, results from similar studies, and other relevant evidence. | p19-20 |
| Implications | 20 | Discuss the potential clinical use of the model and implications for future research. | p20 |
| **Other information** | | | |
| Supplementary information | 21 | Provide information about the availability of supplementary resources, such as study protocol, Web calculator, and data sets. | supplementary |
| Funding | 22 | Give the source of funding and the role of the funders for the present study. | p21 |

**Supplementary Table 3. Characteristics of studies to be validated.**

| **Study** | **Mean/median Age (year)** | **Female (%)** | **Tumor stage III/IV (%)** | **Cancer site** | **Source of patients** | **Size of training set** | **Endpoint** | **Event rate** | **Construct prognostic score based on CpGs** |
| --- | --- | --- | --- | --- | --- | --- | --- | --- | --- |
| Yang et al, 2017^10^ | NR | NR | NR | Colon adenocarcinoma | TCGA | 301 | OS | NR | Yes, but NR |
| Hou et al, 2018^11^ | 64 | 46% | 43% | Colon and rectum | TCGA | 379 | OS | NR | NP |
| Gündert et al, 2019^3^ | 69 | 46% | 41% | Colon and rectum | DACHS cohort | 572 | OS, DSS^1^ | NR | Yes |
| Yang et al, 2019^12^ | 67 | 45% | 43% | Colon adenocarcinoma | TCGA | 272 | OS | Overall deaths: 14% | Yes |
| Gong et al, 2020^13^ | NR | 46% | NR | Colon and rectum | TCGA | 320 | OS | NR | Yes |
| Wang X et al, 2020^14^ | 66 | 43% | 45% | Colon and rectum | TCGA | 249 | OS, PFS^1^ | Cumulative incidence, OS: 3-year: 36.3%, 5-year: 61; PFS: 3-year: 57.9%, 5-year: 76.7% | Yes |
| Wang XY et al, 2020^15^ | 67 | 48% | 41% | Colon | TCGA | 461 | OS | Overall deaths: 19.3% | NP |
| Wang Y et al, 2020^16^ | >64, 60% | 49% | 17% | Colon | TCGA | 143 | OS | NR | Yes |
| Xiang et al, 2020^17^ | 67 | 47% | 40% | Colon adenocarcinoma | TCGA | 385 | OS | NR | Yes |
| Yang et al, 2020^18^ | NR | NR | NR | Colon | TCGA | 225 | OS | NR | Yes, but NR |
| Yin et al, 2020^19^ | >65, 55% | NR | NR | Colon adenocarcinoma | TCGA | 337 | OS | NR | NP |
| Chen et al, 2021^20^ | ≥65, 61% | 46% | 43% | Colon and rectum | TCGA | 127 | OS | Overall deaths: 23.6% | Yes |
| Huang et al, 2021^21^ | 66 | 46% | 47% | Colon and rectum | TCGA | 376 | OS, DFS^1^ | NR | Yes |
| Li et al, 2021^22^ | NR | NR | NR | Colon and rectum | TCGA | 327 | OS | NR | Yes |
| Xin et al, 2022^23^ | NR | 46% | 42% | Colon and rectum | TCGA | 621 | OS | Overall death: 20.5 | NP |

NR = Not reported; TCGA = the Cancer Genome Atlas Program; NP = Not performed; OS = Overall survival; DDS = Disease-specific survival; DFS = Disease-free survival; PFS = Progression-free survival. ^1^ In the three studies, prognostic CpGs were selected based on their associations with OS only, and were additionally validated for the other outcome.

### Supplementary Table 4: Outcomes of validation cohort

|  | **All-cause death** | | **Recurrence** | |
| --- | --- | --- | --- | --- |
|  | **Overall sample *N* = 2310** | **Complete cases^1^ *N* = 1734** | **Overall sample *N* = 2290** | **Complete cases^1^ *N* = 1718** |
| Median follow-up (years, IQR) | 10.43 (10.05-12.40) | 10.51 (10.08-13.01) | 9.81 (5.27-10.23) | 9.84 (5.32-10.24) |
| Total follow-up (person-years) | 17622 | 13535 | 13362 | 10195 |
| Event per 1000 person-years (95% CI) | 71.05 (67.17-75.09) | 70.19 (65.80 -74.80) | 57.03 (53.05-61.23) | 55.32 (50.85-60.08) |
| Number of events | 1252 | 950 | 762 | 564 |
| 3-year cumulative incidence | 21.10% | 21.45% | 23.20% | 23.33% |
| 5-year cumulative incidence | 32.02% | 31.72% | 27.85% | 27.56% |
| 10-year cumulative incidence | 50.73% | 50.33% | 31.35% | 31.10% |

^1^ Excluding sample with any missing values in covariables presented on Table 2.

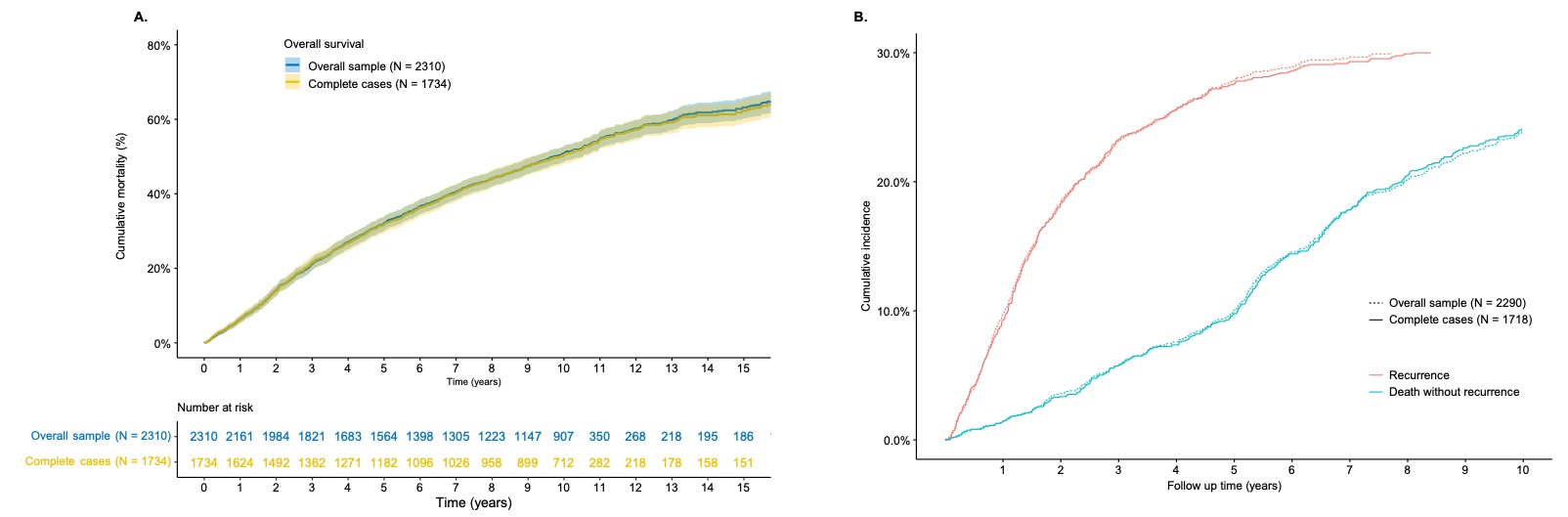

### Supplementary Figure 2: K-M survival curves for OS (A) and cumulative incidence curves for DFS (B) comparing the overall sample and complete cases

### Supplementary Table 5: Cox regression analyses for individual CpGs

| **Study** | **CpG name** | **Endpoint in development study** | **OS** | | | | | | **DFS** | | | | | |
| --- | --- | --- | --- | --- | --- | --- | --- | --- | --- | --- | --- | --- | --- | --- |
|  |  |  | **Unadjusted model** | | **Model 1^1^** | | **Model 2^2^** | | **Unadjusted model** | | **Model 1^1^** | | **Model 2^2^** | |
|  |  |  | **cHR (95%CI)** | ***p* value** | **aHR(95%CI)** | ***p* value** | **aHR(95%CI)** | **p value** | **cHR (95%CI)** | ***p* value** | **aHR(95%CI)** | ***p* value** | **aHR(95%CI)** | ***p* value** |
| Chen et al, 2021^20^ | cg11621464 | OS | 1.08 (1.02, 1.14) | 0.008 | 1.03 (0.97, 1.09) | 0.375 | 1.03 (0.97, 1.09) | 0.394 | 1.08 (1.00, 1.17) | 0.037 | 1.01 (0.94, 1.1) | 0.716 | 1.01 (0.93, 1.09) | 0.798 |
|  | cg13565656 | OS | 1.01 (0.96, 1.07) | 0.724 | 1.01 (0.95, 1.08) | 0.708 | 1.02 (0.95, 1.09) | 0.56 | 0.92 (0.85, 1.00) | 0.041 | 1 (0.91, 1.09) | 0.947 | 1.03 (0.94, 1.13) | 0.514 |
|  | cg18976437 | OS | 1.00 (0.95, 1.06) | 0.865 | 1.01 (0.96, 1.07) | 0.657 | 1.01 (0.96, 1.07) | 0.653 | 0.90 (0.84, 0.97) | 0.007 | 0.97 (0.9, 1.04) | 0.386 | 0.97 (0.9, 1.04) | 0.385 |
|  | cg20505223 | OS | 1.00 (0.95, 1.06) | 0.972 | 0.96 (0.91, 1.01) | 0.145 | 0.96 (0.91, 1.01) | 0.141 | 0.92 (0.86, 0.99) | 0.020 | 0.94 (0.88, 1.01) | 0.116 | 0.94 (0.88, 1.01) | 0.106 |
|  | cg20528583 | OS | 1.02 (0.96, 1.08) | 0.53 | 1 (0.94, 1.06) | 0.956 | 1 (0.94, 1.06) | 0.949 | 0.91 (0.84, 0.97) | 0.008 | 0.96 (0.89, 1.04) | 0.313 | 0.96 (0.89, 1.04) | 0.302 |
| Gong et al, 2020^13^ | cg00110724 | OS | 1.08 (1.01, 1.15) | 0.016 | 1.07 (1.01, 1.15) | 0.026 | 1.08 (1.01, 1.15) | 0.025 | 1.14 (1.04, 1.25) | 0.004 | 1.13 (1.03, 1.23) | 0.011 | 1.13 (1.03, 1.23) | 0.011 |
|  | cg09353563 | OS | 0.87 (0.83, 0.92) | <0.001 | 0.97 (0.92, 1.03) | 0.292 | 0.97 (0.92, 1.03) | 0.341 | 0.80 (0.75, 0.86) | <0.001 | 0.93 (0.86, 0.99) | 0.033 | 0.94 (0.87, 1.01) | 0.083 |
|  | cg14660573 | OS | 0.98 (0.93, 1.04) | 0.582 | 1.01 (0.95, 1.06) | 0.857 | 1.01 (0.95, 1.06) | 0.841 | 0.96 (0.88, 1.03) | 0.257 | 0.97 (0.9, 1.05) | 0.472 | 0.98 (0.9, 1.05) | 0.534 |
| Gündert et al, 2019^3^ | cg00832644 | OS and DSS | 0.91 (0.86, 0.97) | 0.002 | 0.96 (0.9, 1.02) | 0.164 | 0.95 (0.89, 1.02) | 0.16 | 0.88 (0.81, 0.94) | 0.001 | 0.91 (0.84, 0.99) | 0.033 | 0.9 (0.83, 0.98) | 0.016 |
|  | cg01131395 | OS and DSS | 0.84 (0.79, 0.89) | <0.001 | 0.9 (0.85, 0.96) | 0.002 | 0.9 (0.85, 0.96) | 0.002 | 0.72 (0.67, 0.78) | <0.001 | 0.83 (0.76, 0.9) | <0.001 | 0.84 (0.77, 0.91) | <0.001 |
|  | cg05646575 | OS and DSS | 0.79 (0.74, 0.84) | <0.001 | 0.85 (0.8, 0.9) | <0.001 | 0.85 (0.79, 0.9) | <0.001 | 0.71 (0.65, 0.76) | <0.001 | 0.8 (0.74, 0.87) | <0.001 | 0.8 (0.74, 0.86) | <0.001 |
|  | cg08617020 | OS and DSS | 0.88 (0.83, 0.94) | <0.001 | 0.92 (0.86, 0.98) | 0.007 | 0.92 (0.86, 0.98) | 0.007 | 0.86 (0.80, 0.93) | <0.001 | 0.9 (0.83, 0.98) | 0.01 | 0.89 (0.82, 0.96) | 0.004 |
|  | cg08729279 | OS and DSS | 0.88 (0.82, 0.93) | <0.001 | 0.9 (0.85, 0.96) | 0.002 | 0.9 (0.85, 0.96) | 0.002 | 0.82 (0.76, 0.89) | <0.001 | 0.87 (0.8, 0.94) | 0.001 | 0.86 (0.8, 0.94) | <0.001 |
|  | cg08804626 | OS and DSS | 0.85 (0.80, 0.90) | <0.001 | 0.86 (0.81, 0.92) | <0.001 | 0.85 (0.8, 0.91) | <0.001 | 0.81 (0.76, 0.86) | <0.001 | 0.84 (0.78, 0.9) | <0.001 | 0.81 (0.75, 0.87) | <0.001 |
|  | cg10758824 | OS and DSS | 0.88 (0.83, 0.94) | <0.001 | 0.94 (0.88, 1) | 0.068 | 0.94 (0.88, 1) | 0.068 | 0.78 (0.72, 0.85) | <0.001 | 0.89 (0.82, 0.97) | 0.008 | 0.9 (0.83, 0.98) | 0.011 |
|  | cg11056055 | OS and DSS | 0.87 (0.82, 0.92) | <0.001 | 0.9 (0.84, 0.95) | 0.001 | 0.89 (0.84, 0.95) | <0.001 | 0.84 (0.78, 0.91) | <0.001 | 0.85 (0.79, 0.92) | <0.001 | 0.84 (0.77, 0.9) | <0.001 |
|  | cg12510999 | OS and DSS | 0.83 (0.78, 0.88) | <0.001 | 0.88 (0.82, 0.94) | <0.001 | 0.88 (0.82, 0.94) | <0.001 | 0.72 (0.67, 0.78) | <0.001 | 0.83 (0.77, 0.9) | <0.001 | 0.83 (0.77, 0.9) | <0.001 |
|  | cg14270346 | OS and DSS | 0.84 (0.79, 0.89) | <0.001 | 0.9 (0.84, 0.96) | 0.001 | 0.9 (0.84, 0.96) | 0.001 | 0.78 (0.72, 0.84) | <0.001 | 0.84 (0.78, 0.92) | <0.001 | 0.84 (0.77, 0.91) | <0.001 |
|  | cg14983135 | OS and DSS | 0.91 (0.85, 0.97) | 0.002 | 0.94 (0.88, 1) | 0.052 | 0.94 (0.88, 1) | 0.05 | 0.84 (0.77, 0.91) | <0.001 | 0.86 (0.8, 0.94) | 0.001 | 0.86 (0.79, 0.93) | <0.001 |
|  | cg16336556 | OS and DSS | 0.82 (0.77, 0.88) | <0.001 | 0.92 (0.86, 0.98) | 0.013 | 0.92 (0.86, 0.98) | 0.013 | 0.72 (0.66, 0.79) | <0.001 | 0.87 (0.8, 0.95) | 0.001 | 0.87 (0.8, 0.95) | 0.002 |
|  | cg16399624 | OS and DSS | 0.89 (0.84, 0.95) | <0.001 | 0.91 (0.86, 0.97) | 0.003 | 0.91 (0.86, 0.97) | 0.002 | 0.86 (0.79, 0.92) | <0.001 | 0.89 (0.82, 0.96) | 0.002 | 0.88 (0.81, 0.94) | 0.001 |
|  | cg17431888 | OS and DSS | 0.90 (0.84, 0.96) | 0.002 | 0.97 (0.91, 1.04) | 0.392 | 0.97 (0.91, 1.04) | 0.391 | 0.80 (0.73, 0.88) | <0.001 | 0.89 (0.82, 0.97) | 0.011 | 0.9 (0.82, 0.98) | 0.015 |
|  | cg18195165 | OS and DSS | 0.86 (0.81, 0.92) | <0.001 | 0.89 (0.83, 0.94) | <0.001 | 0.89 (0.83, 0.94) | <0.001 | 0.82 (0.76, 0.89) | <0.001 | 0.88 (0.81, 0.95) | 0.001 | 0.86 (0.8, 0.93) | <0.001 |
|  | cg18736676 | OS and DSS | 0.89 (0.84, 0.95) | <0.001 | 0.91 (0.86, 0.97) | 0.002 | 0.91 (0.86, 0.97) | 0.002 | 0.85 (0.79, 0.91) | <0.001 | 0.91 (0.84, 0.98) | 0.012 | 0.91 (0.84, 0.98) | 0.011 |
|  | cg19184885 | OS and DSS | 0.81 (0.76, 0.86) | <0.001 | 0.89 (0.83, 0.95) | <0.001 | 0.89 (0.83, 0.95) | <0.001 | 0.70 (0.65, 0.75) | <0.001 | 0.83 (0.76, 0.9) | <0.001 | 0.83 (0.76, 0.9) | <0.001 |
|  | cg19340296 | OS and DSS | 0.83 (0.78, 0.88) | <0.001 | 0.9 (0.85, 0.97) | 0.003 | 0.9 (0.85, 0.97) | 0.003 | 0.73 (0.69, 0.79) | <0.001 | 0.86 (0.79, 0.93) | <0.001 | 0.86 (0.79, 0.93) | <0.001 |
|  | cg22522598 | OS and DSS | 0.93 (0.87, 0.99) | 0.02 | 0.99 (0.92, 1.05) | 0.683 | 0.99 (0.92, 1.05) | 0.683 | 0.82 (0.76, 0.89) | <0.001 | 0.94 (0.87, 1.03) | 0.184 | 0.94 (0.87, 1.03) | 0.193 |
|  | cg23750514 | OS and DSS | 0.81 (0.77, 0.86) | <0.001 | 0.83 (0.78, 0.88) | <0.001 | 0.82 (0.77, 0.87) | <0.001 | 0.79 (0.73, 0.85) | <0.001 | 0.83 (0.77, 0.9) | <0.001 | 0.82 (0.75, 0.88) | <0.001 |
| Hou et al, 2018^11^ | cg00147009 | OS | 0.91 (0.86, 0.96) | 0.001 | 0.93 (0.87, 0.98) | 0.013 | 0.93 (0.87, 0.98) | 0.012 | 0.87 (0.81, 0.95) | 0.001 | 0.9 (0.82, 0.97) | 0.01 | 0.89 (0.82, 0.97) | 0.008 |
|  | cg00199007 | OS | 0.95 (0.89, 1.01) | 0.09 | 0.97 (0.91, 1.03) | 0.371 | 0.97 (0.91, 1.04) | 0.418 | 0.91 (0.83, 0.99) | 0.030 | 0.96 (0.89, 1.05) | 0.387 | 0.98 (0.9, 1.06) | 0.592 |
|  | cg01467592 | OS | 1.00 (0.95, 1.06) | 0.893 | 1.04 (0.98, 1.1) | 0.175 | 1.04 (0.98, 1.1) | 0.166 | 1.01 (0.94, 1.09) | 0.732 | 1.06 (0.98, 1.15) | 0.124 | 1.07 (0.99, 1.15) | 0.097 |
|  | cg03895094 | OS | 1.01 (0.96, 1.07) | 0.658 | 1 (0.94, 1.06) | 0.95 | 1 (0.94, 1.06) | 0.882 | 1.05 (0.97, 1.13) | 0.270 | 1.02 (0.94, 1.11) | 0.617 | 1.01 (0.93, 1.1) | 0.8 |
|  | cg04138846 | OS | 1.09 (1.02, 1.16) | 0.007 | 1.09 (1.02, 1.16) | 0.009 | 1.09 (1.02, 1.16) | 0.009 | 1.09 (1.01, 1.19) | 0.032 | 1.11 (1.02, 1.21) | 0.012 | 1.11 (1.02, 1.21) | 0.014 |
|  | cg05821816 | OS | 0.92 (0.87, 0.98) | 0.01 | 0.93 (0.87, 0.99) | 0.015 | 0.93 (0.87, 0.99) | 0.015 | 0.92 (0.85, 1.00) | 0.060 | 0.92 (0.85, 0.99) | 0.035 | 0.92 (0.85, 1) | 0.039 |
|  | cg06328127 | OS | 1.08 (1.02, 1.14) | 0.007 | 1.08 (1.02, 1.14) | 0.009 | 1.08 (1.02, 1.14) | 0.009 | 1.08 (1.01, 1.17) | 0.035 | 1.11 (1.03, 1.2) | 0.005 | 1.11 (1.03, 1.2) | 0.007 |
|  | cg06882340 | OS | 0.94 (0.89, 0.99) | 0.019 | 0.98 (0.92, 1.03) | 0.417 | 0.97 (0.92, 1.03) | 0.366 | 0.93 (0.86, 1.00) | 0.042 | 0.97 (0.89, 1.05) | 0.399 | 0.95 (0.88, 1.03) | 0.248 |
|  | cg07206725 | OS | 1.00 (0.94, 1.06) | 0.973 | 1.01 (0.96, 1.07) | 0.686 | 1.01 (0.96, 1.07) | 0.665 | 1.01 (0.94, 1.09) | 0.713 | 1.02 (0.95, 1.1) | 0.601 | 1.02 (0.95, 1.1) | 0.519 |
|  | cg08021727 | OS | 1.13 (1.07, 1.20) | <0.001 | 1.1 (1.03, 1.16) | 0.003 | 1.1 (1.04, 1.17) | 0.002 | 1.24 (1.15, 1.35) | <0.001 | 1.14 (1.04, 1.24) | 0.004 | 1.15 (1.06, 1.25) | 0.001 |
|  | cg10248231 | OS | 1.03 (0.98, 1.09) | 0.248 | 1.02 (0.96, 1.08) | 0.549 | 1.02 (0.96, 1.08) | 0.55 | 1.13 (1.05, 1.21) | 0.002 | 1.05 (0.97, 1.13) | 0.203 | 1.05 (0.98, 1.14) | 0.185 |
|  | cg10584300 | OS | 0.98 (0.93, 1.04) | 0.502 | 0.98 (0.93, 1.03) | 0.444 | 0.98 (0.93, 1.03) | 0.413 | 0.97 (0.90, 1.04) | 0.409 | 0.99 (0.93, 1.06) | 0.761 | 0.98 (0.92, 1.05) | 0.635 |
| Huang, 2021^21^ | cg12751565 | OS and DFS | 1.11 (1.05, 1.17) | <0.001 | 1.08 (1.02, 1.15) | 0.007 | 1.08 (1.02, 1.15) | 0.007 | 1.27 (1.18, 1.38) | <0.001 | 1.16 (1.07, 1.25) | <0.001 | 1.16 (1.07, 1.26) | <0.001 |
|  | cg21614638 | OS and DFS | 1.09 (1.03, 1.15) | 0.002 | 1.04 (0.98, 1.1) | 0.213 | 1.03 (0.97, 1.1) | 0.269 | 1.17 (1.09, 1.26) | <0.001 | 1.05 (0.96, 1.13) | 0.28 | 1.02 (0.94, 1.1) | 0.665 |
|  | cg21770617 | OS and DFS | 1.08 (1.02, 1.14) | 0.01 | 1.11 (1.05, 1.18) | <0.001 | 1.12 (1.06, 1.19) | <0.001 | 1.18 (1.10, 1.27) | <0.001 | 1.13 (1.05, 1.22) | 0.001 | 1.15 (1.07, 1.24) | <0.001 |
| Li et al, 2021^22^ | cg01408654 | OS | 1.02 (0.96, 1.07) | 0.596 | 0.96 (0.91, 1.02) | 0.186 | 0.96 (0.91, 1.02) | 0.181 | 0.90 (0.84, 0.97) | 0.006 | 0.94 (0.88, 1.01) | 0.098 | 0.94 (0.88, 1.01) | 0.093 |
|  | cg04035209 | OS | 1.03 (0.97, 1.09) | 0.339 | 0.96 (0.9, 1.02) | 0.158 | 0.96 (0.9, 1.02) | 0.151 | 0.93 (0.86, 0.99) | 0.034 | 0.94 (0.87, 1.01) | 0.083 | 0.93 (0.86, 1) | 0.063 |
|  | cg10196720 | OS | 1.03 (0.97, 1.09) | 0.338 | 0.97 (0.92, 1.03) | 0.311 | 0.97 (0.91, 1.03) | 0.277 | 0.94 (0.87, 1.01) | 0.075 | 0.94 (0.88, 1.01) | 0.099 | 0.93 (0.87, 1) | 0.055 |
|  | cg10379890 | OS | 1.08 (1.02, 1.14) | 0.008 | 1.04 (0.98, 1.11) | 0.153 | 1.04 (0.98, 1.1) | 0.174 | 0.98 (0.91, 1.05) | 0.524 | 0.99 (0.92, 1.07) | 0.794 | 0.98 (0.91, 1.05) | 0.578 |
|  | cg11097433 | OS | 1.07 (1.01, 1.13) | 0.02 | 1.03 (0.97, 1.09) | 0.382 | 1.02 (0.97, 1.09) | 0.408 | 0.97 (0.90, 1.04) | 0.365 | 0.98 (0.91, 1.06) | 0.592 | 0.97 (0.9, 1.05) | 0.475 |
|  | cg14675211 | OS | 1.06 (1.00, 1.12) | 0.04 | 0.99 (0.94, 1.05) | 0.838 | 0.99 (0.94, 1.05) | 0.743 | 1.00 (0.93, 1.07) | 0.943 | 0.97 (0.9, 1.04) | 0.413 | 0.95 (0.88, 1.03) | 0.197 |
|  | cg15428578 | OS | 1.06 (1.00, 1.12) | 0.037 | 1.02 (0.96, 1.08) | 0.587 | 1.01 (0.96, 1.07) | 0.626 | 0.97 (0.90, 1.04) | 0.366 | 0.99 (0.92, 1.06) | 0.709 | 0.98 (0.91, 1.05) | 0.513 |
|  | cg19343464 | OS | 1.04 (0.98, 1.10) | 0.206 | 0.99 (0.93, 1.05) | 0.713 | 0.99 (0.93, 1.05) | 0.657 | 0.96 (0.89, 1.03) | 0.278 | 0.97 (0.9, 1.05) | 0.484 | 0.96 (0.89, 1.04) | 0.321 |
|  | cg21384402 | OS | 1.07 (1.01, 1.13) | 0.027 | 1 (0.95, 1.06) | 0.936 | 1 (0.95, 1.06) | 0.955 | 0.96 (0.89, 1.04) | 0.308 | 0.97 (0.9, 1.04) | 0.343 | 0.96 (0.89, 1.03) | 0.29 |
|  | cg27404023 | OS | 1.10 (1.04, 1.17) | 0.001 | 1.04 (0.98, 1.1) | 0.2 | 1.04 (0.98, 1.1) | 0.213 | 1.01 (0.93, 1.08) | 0.886 | 1.03 (0.95, 1.11) | 0.469 | 1.02 (0.95, 1.1) | 0.562 |
| Wang X et al, 2020^14^ | cg03091331 | OS and PFS | 0.96 (0.91, 1.02) | 0.161 | 0.99 (0.93, 1.05) | 0.705 | 0.99 (0.93, 1.06) | 0.814 | 0.85 (0.79, 0.92) | <0.001 | 0.96 (0.88, 1.04) | 0.348 | 0.98 (0.91, 1.07) | 0.712 |
|  | cg06884352 | OS and PFS | 1.03 (0.98, 1.09) | 0.276 | 1.06 (1, 1.12) | 0.047 | 1.06 (1, 1.13) | 0.034 | 1.11 (1.03, 1.19) | 0.008 | 1.09 (1.01, 1.18) | 0.021 | 1.11 (1.03, 1.2) | 0.006 |
|  | cg07707546 | OS and PFS | 0.99 (0.94, 1.05) | 0.731 | 0.96 (0.91, 1.02) | 0.21 | 0.97 (0.91, 1.02) | 0.226 | 0.88 (0.82, 0.95) | 0.002 | 0.93 (0.86, 1) | 0.051 | 0.93 (0.86, 1.01) | 0.075 |
|  | cg08081805 | OS and PFS | 1.04 (0.99, 1.10) | 0.138 | 0.98 (0.93, 1.04) | 0.58 | 0.98 (0.93, 1.04) | 0.57 | 0.98 (0.91, 1.06) | 0.624 | 0.97 (0.9, 1.04) | 0.393 | 0.97 (0.9, 1.04) | 0.384 |
|  | cg21347353 | OS and PFS | 0.99 (0.93, 1.04) | 0.652 | 0.98 (0.93, 1.04) | 0.517 | 0.98 (0.93, 1.04) | 0.505 | 0.88 (0.82, 0.96) | 0.002 | 0.94 (0.87, 1.01) | 0.101 | 0.94 (0.87, 1.01) | 0.09 |
|  | cg25164589 | OS and PFS | 0.95 (0.90, 1.01) | 0.098 | 0.97 (0.92, 1.03) | 0.342 | 0.97 (0.92, 1.03) | 0.374 | 0.87 (0.80, 0.94) | <0.001 | 0.92 (0.84, 0.99) | 0.037 | 0.93 (0.85, 1.01) | 0.073 |
| Wang XY et al, 2020^15^ | cg00660989 | OS | 0.93 (0.88, 0.98) | 0.008 | 0.91 (0.86, 0.96) | 0.001 | 0.91 (0.86, 0.96) | 0.001 | 0.86 (0.80, 0.93) | <0.001 | 0.88 (0.82, 0.95) | 0.001 | 0.9 (0.83, 0.97) | 0.005 |
|  | cg00929855 | OS | 0.96 (0.91, 1.02) | 0.194 | 0.98 (0.93, 1.04) | 0.543 | 0.99 (0.93, 1.04) | 0.6 | 0.88 (0.82, 0.95) | 0.001 | 0.92 (0.85, 0.99) | 0.031 | 0.93 (0.87, 1.01) | 0.074 |
|  | cg01340952 | OS | 0.93 (0.88, 0.99) | 0.018 | 0.94 (0.88, 0.99) | 0.026 | 0.94 (0.88, 1) | 0.034 | 0.84 (0.78, 0.91) | <0.001 | 0.88 (0.82, 0.96) | 0.003 | 0.9 (0.83, 0.98) | 0.013 |
|  | cg01639032 | OS | 0.97 (0.92, 1.03) | 0.348 | 0.99 (0.94, 1.05) | 0.851 | 1 (0.94, 1.06) | 0.928 | 0.89 (0.83, 0.96) | 0.002 | 0.94 (0.87, 1.01) | 0.087 | 0.95 (0.88, 1.02) | 0.188 |
|  | cg02704535 | OS | 0.97 (0.92, 1.03) | 0.301 | 1 (0.94, 1.05) | 0.862 | 1 (0.94, 1.06) | 0.939 | 0.89 (0.82, 0.95) | 0.001 | 0.93 (0.86, 1) | 0.059 | 0.94 (0.88, 1.02) | 0.136 |
|  | cg05194618 | OS | 0.93 (0.88, 0.98) | 0.012 | 0.92 (0.87, 0.97) | 0.004 | 0.92 (0.87, 0.98) | 0.005 | 0.85 (0.79, 0.91) | <0.001 | 0.88 (0.82, 0.95) | 0.001 | 0.9 (0.83, 0.97) | 0.006 |
|  | cg10598353 | OS | 0.97 (0.92, 1.02) | 0.262 | 1 (0.95, 1.06) | 0.907 | 1.01 (0.95, 1.06) | 0.836 | 0.89 (0.83, 0.96) | 0.001 | 0.94 (0.88, 1.02) | 0.13 | 0.96 (0.89, 1.03) | 0.254 |
|  | cg11353380 | OS | 0.97 (0.92, 1.03) | 0.297 | 0.99 (0.94, 1.05) | 0.854 | 1 (0.94, 1.06) | 0.933 | 0.89 (0.83, 0.96) | 0.004 | 0.93 (0.86, 1) | 0.065 | 0.94 (0.87, 1.02) | 0.149 |
|  | cg11485463 | OS | 0.97 (0.92, 1.03) | 0.277 | 1 (0.94, 1.05) | 0.87 | 1 (0.94, 1.06) | 0.953 | 0.88 (0.82, 0.95) | 0.001 | 0.94 (0.87, 1.01) | 0.102 | 0.95 (0.88, 1.03) | 0.222 |
|  | cg12643366 | OS | 0.97 (0.92, 1.02) | 0.261 | 0.99 (0.94, 1.05) | 0.736 | 0.99 (0.94, 1.05) | 0.811 | 0.89 (0.83, 0.96) | 0.002 | 0.93 (0.86, 1) | 0.055 | 0.94 (0.87, 1.02) | 0.132 |
|  | cg12883479 | OS | 0.97 (0.91, 1.02) | 0.221 | 1 (0.94, 1.06) | 0.926 | 1 (0.94, 1.06) | 0.994 | 0.88 (0.82, 0.95) | 0.001 | 0.93 (0.87, 1.01) | 0.07 | 0.95 (0.88, 1.02) | 0.146 |
|  | cg13413286 | OS | 0.96 (0.91, 1.01) | 0.15 | 1 (0.94, 1.05) | 0.923 | 1 (0.94, 1.06) | 0.992 | 0.88 (0.82, 0.95) | 0.001 | 0.94 (0.87, 1.01) | 0.101 | 0.95 (0.88, 1.03) | 0.202 |
|  | cg15174834 | OS | 0.97 (0.92, 1.02) | 0.239 | 0.99 (0.94, 1.05) | 0.763 | 0.99 (0.94, 1.05) | 0.835 | 0.88 (0.82, 0.95) | 0.001 | 0.92 (0.86, 1) | 0.037 | 0.94 (0.87, 1.01) | 0.089 |
|  | cg15185479 | OS | 0.97 (0.91, 1.02) | 0.216 | 0.99 (0.94, 1.05) | 0.754 | 0.99 (0.94, 1.05) | 0.813 | 0.88 (0.81, 0.95) | 0.001 | 0.92 (0.85, 0.99) | 0.031 | 0.93 (0.86, 1.01) | 0.067 |
|  | cg17494781 | OS | 0.97 (0.91, 1.02) | 0.213 | 1 (0.94, 1.06) | 0.971 | 1 (0.95, 1.06) | 0.963 | 0.89 (0.82, 0.95) | 0.001 | 0.94 (0.87, 1.01) | 0.098 | 0.95 (0.88, 1.02) | 0.189 |
|  | cg18466674 | OS | 0.97 (0.91, 1.02) | 0.229 | 1 (0.95, 1.06) | 0.995 | 1 (0.95, 1.06) | 0.93 | 0.89 (0.82, 0.95) | 0.001 | 0.94 (0.88, 1.02) | 0.124 | 0.96 (0.89, 1.03) | 0.23 |
|  | cg19677203 | OS | 0.97 (0.92, 1.03) | 0.294 | 1 (0.94, 1.05) | 0.87 | 1 (0.94, 1.06) | 0.952 | 0.88 (0.82, 0.95) | 0.001 | 0.93 (0.86, 1) | 0.066 | 0.95 (0.88, 1.02) | 0.158 |
|  | cg20428713 | OS | 1.02 (0.96, 1.08) | 0.481 | 1.06 (1, 1.12) | 0.061 | 1.07 (1.01, 1.13) | 0.034 | 1.06 (0.98, 1.14) | 0.160 | 1.09 (1.01, 1.18) | 0.023 | 1.13 (1.04, 1.22) | 0.002 |
|  | cg20607287 | OS | 0.99 (0.94, 1.05) | 0.713 | 1 (0.95, 1.06) | 0.969 | 1 (0.95, 1.06) | 0.914 | 0.92 (0.85, 0.99) | 0.020 | 0.96 (0.89, 1.04) | 0.32 | 0.97 (0.9, 1.05) | 0.477 |
|  | cg21096966 | OS | 0.92 (0.87, 0.97) | 0.003 | 0.91 (0.86, 0.96) | 0.001 | 0.91 (0.86, 0.96) | 0.001 | 0.86 (0.80, 0.92) | <0.001 | 0.89 (0.82, 0.96) | 0.002 | 0.91 (0.84, 0.98) | 0.009 |
|  | cg21122656 | OS | 0.97 (0.92, 1.02) | 0.258 | 0.99 (0.94, 1.05) | 0.719 | 0.99 (0.94, 1.05) | 0.788 | 0.88 (0.82, 0.95) | 0.001 | 0.92 (0.86, 1) | 0.038 | 0.94 (0.87, 1.01) | 0.09 |
|  | cg22715094 | OS | 0.95 (0.90, 1.01) | 0.096 | 0.98 (0.92, 1.03) | 0.427 | 0.98 (0.93, 1.04) | 0.482 | 0.87 (0.80, 0.93) | <0.001 | 0.9 (0.84, 0.98) | 0.01 | 0.92 (0.85, 0.99) | 0.027 |
|  | cg22847691 | OS | 0.97 (0.91, 1.02) | 0.23 | 0.99 (0.93, 1.04) | 0.631 | 0.99 (0.93, 1.05) | 0.697 | 0.88 (0.82, 0.95) | 0.001 | 0.92 (0.85, 0.99) | 0.03 | 0.93 (0.87, 1.01) | 0.076 |
|  | cg23285774 | OS | 0.93 (0.88, 0.98) | 0.009 | 0.92 (0.87, 0.97) | 0.002 | 0.91 (0.86, 0.97) | 0.002 | 0.90 (0.83, 0.97) | 0.006 | 0.87 (0.81, 0.94) | 0.001 | 0.87 (0.8, 0.94) | <0.001 |
|  | cg24888257 | OS | 0.95 (0.90, 1.00) | 0.067 | 0.94 (0.89, 0.99) | 0.025 | 0.94 (0.89, 0.99) | 0.026 | 0.92 (0.85, 0.99) | 0.026 | 0.94 (0.87, 1.01) | 0.117 | 0.94 (0.88, 1.02) | 0.132 |
| Wang Y et al, 2020^16^ | cg00177496 | OS | 1.02 (0.96, 1.08) | 0.492 | 1.04 (0.99, 1.1) | 0.131 | 1.04 (0.99, 1.1) | 0.118 | 1.06 (0.99, 1.13) | 0.093 | 1.06 (0.99, 1.14) | 0.088 | 1.06 (0.99, 1.14) | 0.081 |
|  | cg01963906 | OS | 0.94 (0.90, 1.00) | 0.036 | 0.98 (0.92, 1.03) | 0.367 | 0.97 (0.92, 1.03) | 0.362 | 0.89 (0.83, 0.95) | 0.001 | 0.95 (0.89, 1.02) | 0.195 | 0.95 (0.89, 1.03) | 0.205 |
|  | cg05165940 | OS | 1.09 (1.04, 1.15) | 0.001 | 1.13 (1.07, 1.19) | <0.001 | 1.15 (1.09, 1.22) | <0.001 | 1.07 (1.00, 1.15) | 0.055 | 1.14 (1.06, 1.23) | 0.001 | 1.19 (1.1, 1.28) | <0.001 |
|  | cg12921795 | OS | 0.94 (0.89, 1.00) | 0.053 | 0.99 (0.93, 1.04) | 0.624 | 0.99 (0.93, 1.04) | 0.621 | 0.94 (0.87, 1.02) | 0.120 | 0.98 (0.91, 1.05) | 0.557 | 0.98 (0.91, 1.05) | 0.552 |
|  | cg19414598 | OS | 0.99 (0.94, 1.05) | 0.723 | 1.01 (0.96, 1.07) | 0.694 | 1.02 (0.96, 1.07) | 0.604 | 0.92 (0.85, 0.99) | 0.034 | 0.98 (0.9, 1.06) | 0.565 | 0.99 (0.92, 1.08) | 0.898 |
|  | cg25783173 | OS | 1.01 (0.96, 1.07) | 0.654 | 1.03 (0.97, 1.09) | 0.353 | 1.03 (0.97, 1.09) | 0.358 | 0.92 (0.84, 1.02) | 0.106 | 0.95 (0.87, 1.05) | 0.331 | 0.95 (0.87, 1.05) | 0.328 |
| Xiang et al, 2020^17^ | cg03017653 | OS | 1.06 (1.01, 1.12) | 0.029 | 1.07 (1.01, 1.13) | 0.032 | 1.06 (1, 1.13) | 0.044 | 1.14 (1.06, 1.23) | <0.001 | 1.07 (0.99, 1.16) | 0.08 | 1.04 (0.96, 1.13) | 0.292 |
|  | cg03977782 | OS | 0.92 (0.87, 0.98) | 0.008 | 0.93 (0.88, 0.99) | 0.026 | 0.93 (0.87, 0.99) | 0.022 | 0.92 (0.84, 0.99) | 0.033 | 0.92 (0.84, 1) | 0.042 | 0.91 (0.84, 0.99) | 0.029 |
|  | cg05417950 | OS | 0.97 (0.91, 1.03) | 0.289 | 1.01 (0.96, 1.08) | 0.631 | 1.02 (0.96, 1.08) | 0.614 | 0.92 (0.84, 0.99) | 0.031 | 0.98 (0.91, 1.06) | 0.594 | 0.98 (0.91, 1.06) | 0.656 |
|  | cg06250108 | OS | 0.95 (0.89, 1.01) | 0.105 | 0.99 (0.93, 1.05) | 0.692 | 0.99 (0.93, 1.05) | 0.68 | 0.93 (0.85, 1.02) | 0.116 | 0.97 (0.89, 1.05) | 0.469 | 0.97 (0.89, 1.05) | 0.442 |
|  | cg09893305 | OS | 0.96 (0.91, 1.02) | 0.18 | 1.01 (0.96, 1.08) | 0.633 | 1.02 (0.96, 1.09) | 0.467 | 0.87 (0.81, 0.94) | <0.001 | 0.95 (0.88, 1.03) | 0.228 | 0.99 (0.91, 1.07) | 0.744 |
|  | cg10414946 | OS | 1.00 (0.94, 1.05) | 0.869 | 1.04 (0.98, 1.1) | 0.166 | 1.04 (0.99, 1.11) | 0.146 | 1.06 (0.98, 1.14) | 0.150 | 1.04 (0.97, 1.13) | 0.278 | 1.06 (0.98, 1.14) | 0.168 |
|  | cg15170424 | OS | 0.95 (0.89, 1.01) | 0.095 | 0.98 (0.92, 1.04) | 0.535 | 0.98 (0.93, 1.05) | 0.617 | 0.88 (0.80, 0.98) | 0.016 | 0.97 (0.88, 1.08) | 0.607 | 1.01 (0.91, 1.11) | 0.844 |
|  | cg15639045 | OS | 0.99 (0.93, 1.05) | 0.812 | 0.97 (0.91, 1.03) | 0.304 | 0.97 (0.91, 1.03) | 0.351 | 1.02 (0.95, 1.10) | 0.574 | 0.97 (0.9, 1.04) | 0.387 | 0.98 (0.91, 1.05) | 0.619 |
|  | cg15786837 | OS | 1.04 (0.98, 1.10) | 0.184 | 1.07 (1.01, 1.13) | 0.018 | 1.08 (1.02, 1.15) | 0.006 | 0.94 (0.86, 1.02) | 0.151 | 1.02 (0.93, 1.11) | 0.695 | 1.06 (0.98, 1.16) | 0.165 |
|  | cg17329249 | OS | 0.98 (0.93, 1.04) | 0.527 | 1 (0.93, 1.06) | 0.89 | 1 (0.94, 1.06) | 0.907 | 0.99 (0.92, 1.07) | 0.772 | 1 (0.92, 1.08) | 0.937 | 1 (0.92, 1.09) | 0.98 |
|  | cg21212956 | OS | 0.99 (0.93, 1.05) | 0.664 | 1 (0.94, 1.06) | 0.949 | 1 (0.94, 1.06) | 0.985 | 0.95 (0.88, 1.03) | 0.210 | 0.95 (0.87, 1.03) | 0.212 | 0.94 (0.87, 1.03) | 0.178 |
|  | cg24206256 | OS | 0.99 (0.93, 1.05) | 0.749 | 1 (0.94, 1.06) | 0.975 | 1 (0.94, 1.06) | 0.997 | 1.00 (0.92, 1.08) | 0.963 | 1.02 (0.94, 1.1) | 0.708 | 1.02 (0.94, 1.11) | 0.634 |
| Xin et al, 2021^23^ | cg00117463 | OS | 1.03 (0.97, 1.09) | 0.347 | 1.02 (0.96, 1.08) | 0.471 | 1.03 (0.97, 1.09) | 0.395 | 0.95 (0.88, 1.02) | 0.175 | 0.95 (0.87, 1.02) | 0.171 | 0.96 (0.89, 1.04) | 0.357 |
|  | cg00688963 | OS | 1.03 (0.97, 1.08) | 0.35 | 1 (0.94, 1.05) | 0.871 | 1 (0.94, 1.06) | 0.971 | 0.92 (0.85, 1.01) | 0.073 | 0.95 (0.87, 1.04) | 0.278 | 0.98 (0.89, 1.07) | 0.584 |
|  | cg02333852 | OS | 1.02 (0.97, 1.08) | 0.489 | 0.99 (0.94, 1.05) | 0.834 | 1 (0.94, 1.06) | 0.932 | 0.91 (0.83, 0.99) | 0.032 | 0.95 (0.86, 1.04) | 0.228 | 0.97 (0.88, 1.06) | 0.491 |
|  | cg11444379 | OS | 1.03 (0.98, 1.09) | 0.233 | 1.02 (0.96, 1.08) | 0.577 | 1.02 (0.96, 1.08) | 0.5 | 0.94 (0.87, 1.01) | 0.100 | 0.99 (0.91, 1.07) | 0.785 | 1.01 (0.93, 1.09) | 0.89 |
|  | cg12181621 | OS | 1.02 (0.96, 1.08) | 0.485 | 1.02 (0.96, 1.08) | 0.599 | 1.02 (0.96, 1.08) | 0.584 | 0.99 (0.92, 1.07) | 0.842 | 0.98 (0.9, 1.06) | 0.608 | 0.98 (0.91, 1.07) | 0.687 |
|  | cg13442960 | OS | 0.99 (0.94, 1.05) | 0.851 | 0.99 (0.92, 1.06) | 0.743 | 0.99 (0.92, 1.06) | 0.774 | 0.90 (0.83, 0.97) | 0.006 | 0.97 (0.87, 1.07) | 0.512 | 0.97 (0.88, 1.08) | 0.59 |
|  | cg14026459 | OS | 1.03 (0.98, 1.09) | 0.283 | 1.03 (0.96, 1.1) | 0.383 | 1.03 (0.96, 1.1) | 0.384 | 0.94 (0.87, 1.02) | 0.122 | 0.99 (0.91, 1.09) | 0.907 | 0.99 (0.91, 1.09) | 0.88 |
|  | cg19838156 | OS | 0.98 (0.93, 1.04) | 0.509 | 0.99 (0.93, 1.05) | 0.667 | 0.99 (0.93, 1.05) | 0.727 | 0.92 (0.85, 1.00) | 0.044 | 0.97 (0.89, 1.05) | 0.463 | 0.98 (0.9, 1.06) | 0.627 |
|  | cg21200129 | OS | 1.01 (0.96, 1.07) | 0.723 | 1.01 (0.93, 1.08) | 0.881 | 1.01 (0.94, 1.08) | 0.846 | 0.92 (0.85, 0.99) | 0.028 | 1 (0.9, 1.11) | 0.991 | 1.01 (0.91, 1.11) | 0.916 |
|  | cg23849078 | OS | 0.97 (0.91, 1.02) | 0.247 | 0.98 (0.92, 1.04) | 0.43 | 0.98 (0.92, 1.04) | 0.492 | 0.89 (0.82, 0.97) | 0.007 | 0.95 (0.87, 1.03) | 0.218 | 0.96 (0.88, 1.05) | 0.39 |
|  | cg26592281 | OS | 0.99 (0.94, 1.05) | 0.72 | 0.99 (0.94, 1.05) | 0.849 | 1 (0.94, 1.06) | 0.899 | 0.93 (0.85, 1.01) | 0.067 | 0.97 (0.9, 1.06) | 0.542 | 0.98 (0.9, 1.07) | 0.654 |
|  | cg26796283 | OS | 0.99 (0.94, 1.05) | 0.839 | 0.96 (0.91, 1.03) | 0.27 | 0.96 (0.91, 1.03) | 0.271 | 1.04 (0.96, 1.11) | 0.339 | 0.97 (0.9, 1.06) | 0.546 | 0.97 (0.9, 1.06) | 0.53 |
| Yang et al, 2017^10^ | cg00584485 | OS | 1.02 (0.97, 1.08) | 0.442 | 1.01 (0.96, 1.07) | 0.634 | 1.01 (0.96, 1.07) | 0.615 | 1.08 (1.00, 1.17) | 0.042 | 1.06 (0.98, 1.14) | 0.132 | 1.06 (0.99, 1.14) | 0.104 |
|  | cg00854817 | OS | 0.99 (0.94, 1.05) | 0.847 | 0.95 (0.9, 1.01) | 0.114 | 0.95 (0.9, 1.01) | 0.112 | 0.87 (0.81, 0.93) | <0.001 | 0.9 (0.84, 0.97) | 0.007 | 0.9 (0.84, 0.97) | 0.006 |
|  | cg02457680 | OS | 1.09 (1.03, 1.15) | 0.005 | 1.03 (0.97, 1.09) | 0.344 | 1.02 (0.97, 1.08) | 0.386 | 1.00 (0.93, 1.08) | 0.990 | 0.98 (0.92, 1.06) | 0.651 | 0.97 (0.91, 1.04) | 0.44 |
|  | cg03890037 | OS | 1.09 (1.03, 1.15) | 0.004 | 1.02 (0.96, 1.09) | 0.432 | 1.02 (0.96, 1.09) | 0.447 | 0.97 (0.91, 1.05) | 0.476 | 0.96 (0.89, 1.04) | 0.353 | 0.96 (0.89, 1.04) | 0.313 |
|  | cg04929703 | OS | 1.08 (1.02, 1.14) | 0.009 | 1 (0.95, 1.06) | 0.91 | 1 (0.94, 1.06) | 0.986 | 0.97 (0.90, 1.05) | 0.445 | 0.97 (0.9, 1.05) | 0.429 | 0.96 (0.89, 1.03) | 0.248 |
|  | cg05917460 | OS | 1.01 (0.96, 1.07) | 0.707 | 1.02 (0.96, 1.09) | 0.479 | 1.03 (0.97, 1.09) | 0.392 | 0.93 (0.86, 1.00) | 0.046 | 0.99 (0.91, 1.07) | 0.802 | 1.01 (0.93, 1.09) | 0.817 |
| Yang et al, 2017^10^ | cg07936950 | OS | 1.07 (1.01, 1.13) | 0.016 | 1.06 (1, 1.13) | 0.06 | 1.07 (1.01, 1.14) | 0.032 | 0.99 (0.91, 1.06) | 0.704 | 1.06 (0.97, 1.15) | 0.186 | 1.09 (1.01, 1.18) | 0.037 |
|  | cg08152546 | OS | 1.03 (0.98, 1.09) | 0.258 | 1.01 (0.95, 1.06) | 0.854 | 1 (0.95, 1.06) | 0.887 | 0.95 (0.89, 1.03) | 0.209 | 0.96 (0.9, 1.04) | 0.31 | 0.96 (0.89, 1.03) | 0.244 |
|  | cg08318726 | OS | 1.03 (0.98, 1.09) | 0.25 | 0.98 (0.93, 1.04) | 0.552 | 0.98 (0.93, 1.04) | 0.552 | 0.90 (0.84, 0.97) | 0.004 | 0.94 (0.87, 1.01) | 0.083 | 0.94 (0.87, 1.01) | 0.085 |
|  | cg08358166 | OS | 1.05 (0.99, 1.11) | 0.108 | 1.01 (0.95, 1.07) | 0.752 | 1.01 (0.95, 1.07) | 0.784 | 0.94 (0.88, 1.01) | 0.099 | 0.96 (0.9, 1.04) | 0.317 | 0.96 (0.89, 1.03) | 0.252 |
|  | cg09632907 | OS | 1.03 (0.98, 1.09) | 0.255 | 1.03 (0.97, 1.09) | 0.398 | 1.03 (0.97, 1.09) | 0.362 | 0.92 (0.85, 0.99) | 0.020 | 0.98 (0.91, 1.06) | 0.689 | 0.99 (0.92, 1.07) | 0.854 |
|  | cg10023272 | OS | 1.05 (1.00, 1.11) | 0.069 | 1 (0.94, 1.06) | 0.932 | 1 (0.94, 1.06) | 0.904 | 0.93 (0.86, 1.00) | 0.046 | 0.96 (0.89, 1.03) | 0.257 | 0.95 (0.88, 1.03) | 0.199 |
|  | cg12395205 | OS | 1.02 (0.97, 1.08) | 0.407 | 1 (0.94, 1.06) | 0.953 | 1 (0.94, 1.06) | 0.95 | 0.91 (0.85, 0.98) | 0.013 | 0.97 (0.9, 1.04) | 0.39 | 0.97 (0.9, 1.04) | 0.36 |
|  | cg12630714 | OS | 0.99 (0.93, 1.04) | 0.634 | 1.02 (0.96, 1.08) | 0.575 | 1.02 (0.96, 1.08) | 0.479 | 0.89 (0.83, 0.96) | 0.003 | 0.98 (0.91, 1.06) | 0.652 | 1 (0.93, 1.09) | 0.931 |
|  | cg13066016 | OS | 1.07 (1.01, 1.13) | 0.02 | 1.05 (0.99, 1.12) | 0.081 | 1.05 (0.99, 1.12) | 0.075 | 0.97 (0.90, 1.05) | 0.500 | 1.05 (0.97, 1.13) | 0.259 | 1.05 (0.97, 1.14) | 0.206 |
|  | cg13596833 | OS | 1.03 (0.98, 1.09) | 0.279 | 1 (0.94, 1.05) | 0.886 | 1 (0.94, 1.06) | 0.899 | 0.93 (0.87, 1.01) | 0.071 | 0.97 (0.9, 1.05) | 0.512 | 0.98 (0.91, 1.05) | 0.555 |
|  | cg14036143 | OS | 1.01 (0.96, 1.07) | 0.606 | 1.02 (0.97, 1.08) | 0.446 | 1.02 (0.97, 1.08) | 0.431 | 0.88 (0.82, 0.95) | 0.001 | 0.95 (0.88, 1.03) | 0.204 | 0.96 (0.89, 1.03) | 0.252 |
|  | cg15105326 | OS | 1.00 (0.94, 1.05) | 0.898 | 0.95 (0.9, 1.01) | 0.092 | 0.95 (0.9, 1) | 0.072 | 0.91 (0.84, 0.97) | 0.008 | 0.91 (0.85, 0.98) | 0.015 | 0.9 (0.84, 0.97) | 0.005 |
|  | cg15916004 | OS | 1.01 (0.95, 1.06) | 0.793 | 0.97 (0.92, 1.03) | 0.341 | 0.97 (0.92, 1.03) | 0.286 | 0.93 (0.86, 1.00) | 0.037 | 0.93 (0.86, 1) | 0.037 | 0.91 (0.85, 0.98) | 0.015 |
|  | cg17495087 | OS | 1.02 (0.96, 1.07) | 0.599 | 1.02 (0.96, 1.08) | 0.583 | 1.02 (0.96, 1.08) | 0.541 | 0.94 (0.88, 1.01) | 0.113 | 1 (0.93, 1.08) | 0.954 | 1.01 (0.93, 1.09) | 0.854 |
|  | cg18131582 | OS | 0.96 (0.91, 1.02) | 0.174 | 0.99 (0.94, 1.05) | 0.71 | 0.99 (0.94, 1.05) | 0.808 | 0.94 (0.87, 1.01) | 0.082 | 1 (0.93, 1.08) | 0.907 | 1.02 (0.95, 1.1) | 0.569 |
|  | cg18443359 | OS | 1.07 (1.01, 1.13) | 0.024 | 1.01 (0.96, 1.07) | 0.639 | 1.01 (0.95, 1.07) | 0.702 | 0.98 (0.91, 1.05) | 0.531 | 0.96 (0.89, 1.03) | 0.283 | 0.95 (0.88, 1.02) | 0.153 |
|  | cg19624849 | OS | 1.00 (0.95, 1.06) | 0.922 | 1.01 (0.96, 1.07) | 0.637 | 1.02 (0.96, 1.08) | 0.613 | 0.88 (0.82, 0.94) | <0.001 | 0.95 (0.88, 1.03) | 0.197 | 0.96 (0.89, 1.03) | 0.257 |
|  | cg20661138 | OS | 1.03 (0.97, 1.09) | 0.347 | 1 (0.94, 1.06) | 0.969 | 1 (0.94, 1.06) | 0.981 | 0.93 (0.87, 1.00) | 0.051 | 0.98 (0.91, 1.06) | 0.598 | 0.98 (0.91, 1.05) | 0.576 |
|  | cg21695661 | OS | 1.06 (1.00, 1.12) | 0.06 | 1.03 (0.97, 1.09) | 0.331 | 1.03 (0.97, 1.09) | 0.323 | 0.93 (0.87, 1.01) | 0.079 | 0.99 (0.91, 1.07) | 0.738 | 0.99 (0.92, 1.07) | 0.796 |
|  | cg22488797 | OS | 1.07 (1.01, 1.13) | 0.026 | 1.03 (0.97, 1.09) | 0.293 | 1.03 (0.97, 1.09) | 0.342 | 1.00 (0.92, 1.07) | 0.897 | 1.01 (0.93, 1.08) | 0.876 | 0.99 (0.92, 1.06) | 0.775 |
|  | cg24804195 | OS | 1.08 (1.02, 1.14) | 0.007 | 1.01 (0.96, 1.07) | 0.676 | 1.01 (0.96, 1.07) | 0.674 | 0.98 (0.91, 1.06) | 0.631 | 0.98 (0.91, 1.06) | 0.663 | 0.98 (0.91, 1.06) | 0.682 |
| Yang et al, 2019^12^ | cg02196655 | OS | 0.93 (0.88, 0.99) | 0.015 | 0.98 (0.92, 1.03) | 0.436 | 0.98 (0.92, 1.04) | 0.444 | 0.88 (0.81, 0.95) | 0.002 | 0.95 (0.88, 1.03) | 0.215 | 0.95 (0.88, 1.03) | 0.242 |
|  | cg03763616 | OS | 0.98 (0.92, 1.03) | 0.393 | 1.01 (0.96, 1.07) | 0.687 | 1.02 (0.96, 1.08) | 0.586 | 0.89 (0.82, 0.96) | 0.004 | 0.95 (0.88, 1.03) | 0.225 | 0.97 (0.89, 1.06) | 0.515 |
|  | cg03944089 | OS | 0.97 (0.92, 1.03) | 0.345 | 1.02 (0.96, 1.08) | 0.517 | 1.02 (0.96, 1.08) | 0.477 | 0.95 (0.88, 1.03) | 0.218 | 0.99 (0.92, 1.07) | 0.877 | 1 (0.93, 1.08) | 0.901 |
|  | cg06117855 | OS | 1.01 (0.96, 1.07) | 0.725 | 1.04 (0.98, 1.1) | 0.175 | 1.04 (0.98, 1.1) | 0.161 | 0.94 (0.87, 1.02) | 0.126 | 0.96 (0.88, 1.04) | 0.279 | 0.97 (0.89, 1.04) | 0.382 |
|  | cg07173760 | OS | 0.97 (0.91, 1.02) | 0.213 | 1.02 (0.97, 1.08) | 0.465 | 1.03 (0.97, 1.09) | 0.381 | 0.93 (0.86, 1.00) | 0.064 | 0.99 (0.92, 1.07) | 0.878 | 1.02 (0.94, 1.1) | 0.66 |
|  | cg07293947 | OS | 1.01 (0.95, 1.07) | 0.704 | 1.04 (0.98, 1.1) | 0.194 | 1.04 (0.99, 1.11) | 0.147 | 1.03 (0.95, 1.11) | 0.457 | 1.06 (0.99, 1.13) | 0.109 | 1.07 (1.01, 1.15) | 0.035 |
|  | cg07509155 | OS | 0.91 (0.86, 0.97) | 0.002 | 0.99 (0.93, 1.04) | 0.605 | 0.99 (0.93, 1.05) | 0.708 | 0.87 (0.80, 0.94) | 0.001 | 0.96 (0.89, 1.04) | 0.344 | 0.99 (0.91, 1.07) | 0.724 |
|  | cg09244244 | OS | 0.93 (0.87, 0.98) | 0.008 | 0.98 (0.93, 1.04) | 0.466 | 0.98 (0.93, 1.04) | 0.525 | 0.87 (0.81, 0.95) | 0.001 | 0.94 (0.87, 1.01) | 0.113 | 0.95 (0.88, 1.03) | 0.201 |
|  | cg10451565 | OS | 1.05 (1.00, 1.11) | 0.065 | 1.04 (0.98, 1.1) | 0.202 | 1.04 (0.98, 1.1) | 0.186 | 1.06 (0.98, 1.14) | 0.150 | 1.01 (0.94, 1.09) | 0.765 | 1.02 (0.95, 1.1) | 0.608 |
|  | cg12582008 | OS | 1.00 (0.95, 1.06) | 0.954 | 1 (0.95, 1.06) | 0.983 | 1.01 (0.95, 1.06) | 0.86 | 0.97 (0.90, 1.05) | 0.476 | 0.97 (0.9, 1.04) | 0.36 | 0.99 (0.92, 1.07) | 0.833 |
|  | cg13796218 | OS | 1.02 (0.97, 1.08) | 0.451 | 1 (0.94, 1.06) | 0.99 | 1 (0.95, 1.07) | 0.874 | 0.94 (0.86, 1.02) | 0.147 | 0.95 (0.88, 1.04) | 0.256 | 0.98 (0.9, 1.07) | 0.622 |
|  | cg20247048 | OS | 1.08 (1.02, 1.13) | 0.005 | 1.04 (0.99, 1.1) | 0.129 | 1.04 (0.99, 1.1) | 0.141 | 1.16 (1.09, 1.24) | <0.001 | 1.06 (0.99, 1.14) | 0.108 | 1.05 (0.98, 1.13) | 0.133 |
|  | cg21481775 | OS | 0.93 (0.88, 0.99) | 0.024 | 0.96 (0.91, 1.02) | 0.205 | 0.96 (0.91, 1.02) | 0.23 | 0.85 (0.78, 0.93) | <0.001 | 0.9 (0.83, 0.98) | 0.02 | 0.91 (0.84, 0.99) | 0.038 |
|  | cg23829949 | OS | 0.96 (0.90, 1.01) | 0.113 | 1.01 (0.96, 1.07) | 0.617 | 1.02 (0.97, 1.09) | 0.435 | 0.96 (0.89, 1.03) | 0.259 | 1.01 (0.93, 1.08) | 0.877 | 1.05 (0.97, 1.13) | 0.244 |
|  | cg23964386 | OS | 1.01 (0.95, 1.07) | 0.781 | 1.03 (0.97, 1.09) | 0.346 | 1.03 (0.97, 1.09) | 0.299 | 0.95 (0.88, 1.03) | 0.234 | 0.99 (0.92, 1.07) | 0.799 | 1 (0.93, 1.08) | 0.926 |
|  | cg24127989 | OS | 1.05 (0.99, 1.11) | 0.084 | 1.02 (0.97, 1.08) | 0.364 | 1.03 (0.97, 1.08) | 0.341 | 1.07 (1.00, 1.15) | 0.050 | 1.03 (0.96, 1.09) | 0.456 | 1.03 (0.96, 1.1) | 0.406 |
|  | cg24674703 | OS | 1.10 (1.04, 1.17) | <0.001 | 1.09 (1.03, 1.15) | 0.004 | 1.09 (1.03, 1.15) | 0.003 | 1.20 (1.11, 1.29) | <0.001 | 1.09 (1.01, 1.18) | 0.023 | 1.1 (1.02, 1.18) | 0.014 |
|  | cg24938727 | OS | 0.94 (0.89, 1.00) | 0.043 | 0.97 (0.92, 1.03) | 0.278 | 0.97 (0.92, 1.03) | 0.311 | 0.95 (0.88, 1.03) | 0.197 | 0.98 (0.9, 1.06) | 0.564 | 0.99 (0.91, 1.07) | 0.763 |
| Yang et al, 2020^18^ | cg00817367 | OS | 1.04 (0.99, 1.11) | 0.131 | 0.99 (0.93, 1.05) | 0.715 | 0.99 (0.93, 1.05) | 0.677 | 0.95 (0.88, 1.02) | 0.173 | 0.94 (0.87, 1.02) | 0.127 | 0.94 (0.87, 1.01) | 0.085 |
|  | cg02037307 | OS | 1.06 (1.00, 1.13) | 0.04 | 1.01 (0.95, 1.07) | 0.821 | 1 (0.95, 1.06) | 0.904 | 1.02 (0.95, 1.10) | 0.577 | 1.02 (0.94, 1.1) | 0.694 | 1 (0.93, 1.08) | 0.969 |
| Yang et al, 2020^18^ | cg02245020 | OS | 0.99 (0.93, 1.04) | 0.631 | 0.95 (0.9, 1.01) | 0.091 | 0.95 (0.9, 1) | 0.069 | 0.94 (0.87, 1.01) | 0.070 | 0.91 (0.85, 0.99) | 0.019 | 0.9 (0.83, 0.97) | 0.005 |
|  | cg03403065 | OS | 1.05 (0.99, 1.11) | 0.113 | 0.98 (0.93, 1.04) | 0.505 | 0.98 (0.92, 1.03) | 0.411 | 0.97 (0.90, 1.05) | 0.450 | 0.98 (0.91, 1.05) | 0.557 | 0.96 (0.89, 1.03) | 0.25 |
|  | cg04992638 | OS | 1.07 (1.01, 1.13) | 0.023 | 1.05 (1, 1.12) | 0.068 | 1.05 (1, 1.12) | 0.073 | 1.03 (0.96, 1.11) | 0.443 | 1.03 (0.95, 1.11) | 0.463 | 1.02 (0.95, 1.1) | 0.549 |
|  | cg06176750 | OS | 0.99 (0.93, 1.04) | 0.618 | 1.03 (0.97, 1.09) | 0.303 | 1.03 (0.98, 1.09) | 0.258 | 1.00 (0.93, 1.08) | 0.952 | 1.02 (0.94, 1.09) | 0.695 | 1.03 (0.96, 1.11) | 0.453 |
|  | cg06218079 | OS | 0.87 (0.82, 0.92) | <0.001 | 0.99 (0.93, 1.05) | 0.633 | 0.99 (0.93, 1.05) | 0.692 | 0.82 (0.76, 0.89) | <0.001 | 0.97 (0.9, 1.05) | 0.455 | 0.98 (0.91, 1.07) | 0.695 |
|  | cg06531379 | OS | 1.03 (0.97, 1.09) | 0.351 | 1 (0.95, 1.06) | 0.988 | 1 (0.94, 1.05) | 0.867 | 1.01 (0.94, 1.09) | 0.825 | 0.98 (0.91, 1.06) | 0.581 | 0.95 (0.88, 1.03) | 0.217 |
|  | cg06952671 | OS | 1.09 (1.03, 1.16) | 0.003 | 1.06 (1, 1.13) | 0.036 | 1.06 (1, 1.13) | 0.044 | 1.04 (0.96, 1.12) | 0.352 | 1.05 (0.97, 1.13) | 0.246 | 1.03 (0.95, 1.11) | 0.437 |
|  | cg07162000 | OS | 0.95 (0.90, 1.00) | 0.067 | 1.01 (0.95, 1.06) | 0.85 | 1 (0.95, 1.06) | 0.876 | 0.92 (0.85, 0.99) | 0.037 | 1 (0.93, 1.08) | 0.989 | 1 (0.92, 1.08) | 0.951 |
|  | cg13294849 | OS | 1.04 (0.99, 1.10) | 0.151 | 1 (0.94, 1.06) | 0.967 | 1 (0.94, 1.06) | 0.959 | 0.95 (0.88, 1.02) | 0.127 | 0.97 (0.9, 1.05) | 0.484 | 0.97 (0.9, 1.05) | 0.459 |
|  | cg14175690 | OS | 1.16 (1.09, 1.23) | <0.001 | 1.08 (1.01, 1.15) | 0.016 | 1.08 (1.01, 1.14) | 0.019 | 1.07 (0.99, 1.16) | 0.088 | 1.03 (0.95, 1.11) | 0.461 | 1.02 (0.95, 1.11) | 0.58 |
|  | cg14817541 | OS | 1.07 (1.01, 1.13) | 0.026 | 1.02 (0.97, 1.08) | 0.432 | 1.02 (0.97, 1.08) | 0.447 | 0.98 (0.91, 1.06) | 0.626 | 1.01 (0.93, 1.08) | 0.876 | 1 (0.93, 1.08) | 0.953 |
|  | cg14861089 | OS | 1.07 (1.01, 1.14) | 0.018 | 1.03 (0.97, 1.09) | 0.357 | 1.03 (0.97, 1.09) | 0.38 | 0.97 (0.90, 1.04) | 0.379 | 0.95 (0.89, 1.03) | 0.213 | 0.95 (0.88, 1.02) | 0.142 |
|  | cg15848890 | OS | 0.97 (0.91, 1.02) | 0.222 | 1 (0.95, 1.06) | 0.956 | 1 (0.95, 1.06) | 0.898 | 1.02 (0.95, 1.10) | 0.547 | 1.04 (0.96, 1.12) | 0.339 | 1.05 (0.97, 1.13) | 0.205 |
|  | cg16863522 | OS | 1.01 (0.96, 1.07) | 0.693 | 1.02 (0.96, 1.08) | 0.546 | 1.02 (0.96, 1.08) | 0.525 | 1.03 (0.95, 1.10) | 0.491 | 1.01 (0.94, 1.09) | 0.822 | 1.02 (0.94, 1.09) | 0.679 |
|  | cg17300544 | OS | 1.01 (0.96, 1.07) | 0.673 | 1 (0.95, 1.06) | 0.973 | 1 (0.94, 1.06) | 0.906 | 1.00 (0.92, 1.07) | 0.908 | 0.98 (0.91, 1.06) | 0.6 | 0.96 (0.89, 1.04) | 0.29 |
|  | cg19114576 | OS | 0.97 (0.91, 1.02) | 0.227 | 0.99 (0.93, 1.04) | 0.609 | 0.99 (0.93, 1.06) | 0.833 | 0.96 (0.89, 1.04) | 0.301 | 1.01 (0.93, 1.09) | 0.793 | 1.07 (0.98, 1.16) | 0.117 |
|  | cg27361134 | OS | 1.00 (0.95, 1.06) | 0.911 | 1 (0.94, 1.06) | 0.924 | 1 (0.94, 1.06) | 0.92 | 0.94 (0.87, 1.01) | 0.079 | 1 (0.93, 1.08) | 0.999 | 1 (0.93, 1.08) | 0.981 |
| Yin et al, 2020^19^ | cg01102158 | OS | 0.94 (0.88, 1.00) | 0.059 | 0.97 (0.91, 1.03) | 0.281 | 0.97 (0.91, 1.03) | 0.311 | 0.92 (0.84, 1.01) | 0.098 | 0.97 (0.88, 1.06) | 0.486 | 0.98 (0.89, 1.08) | 0.672 |
|  | cg02009256 | OS | 0.96 (0.90, 1.02) | 0.21 | 0.97 (0.91, 1.03) | 0.329 | 0.97 (0.91, 1.03) | 0.361 | 0.86 (0.77, 0.96) | 0.006 | 0.92 (0.82, 1.02) | 0.103 | 0.93 (0.84, 1.03) | 0.183 |
|  | cg02654360 | OS | 0.95 (0.90, 1.01) | 0.105 | 0.95 (0.89, 1) | 0.069 | 0.95 (0.89, 1.01) | 0.08 | 0.87 (0.80, 0.95) | 0.002 | 0.89 (0.82, 0.97) | 0.008 | 0.9 (0.82, 0.98) | 0.019 |
|  | cg02760766 | OS | 1.05 (1.00, 1.11) | 0.04 | 1.05 (1, 1.1) | 0.069 | 1.05 (1, 1.1) | 0.075 | 1.08 (1.02, 1.14) | 0.010 | 1.04 (0.98, 1.11) | 0.154 | 1.04 (0.98, 1.11) | 0.183 |
|  | cg04096096 | OS | 0.97 (0.92, 1.03) | 0.323 | 0.96 (0.91, 1.02) | 0.227 | 0.96 (0.91, 1.02) | 0.216 | 0.92 (0.85, 1.00) | 0.052 | 0.93 (0.86, 1.01) | 0.107 | 0.93 (0.86, 1.01) | 0.098 |
|  | cg04353251 | OS | 1.01 (0.96, 1.07) | 0.618 | 1.02 (0.97, 1.08) | 0.404 | 1.03 (0.97, 1.09) | 0.335 | 0.98 (0.90, 1.06) | 0.562 | 1.02 (0.94, 1.1) | 0.672 | 1.03 (0.96, 1.12) | 0.406 |
|  | cg06671690 | OS | 0.97 (0.92, 1.03) | 0.339 | 0.99 (0.93, 1.05) | 0.655 | 0.99 (0.93, 1.06) | 0.826 | 0.89 (0.81, 0.98) | 0.020 | 0.96 (0.87, 1.06) | 0.47 | 1.01 (0.91, 1.12) | 0.816 |
|  | cg06685724 | OS | 0.93 (0.88, 0.99) | 0.025 | 0.96 (0.91, 1.02) | 0.186 | 0.96 (0.91, 1.02) | 0.198 | 0.91 (0.84, 0.99) | 0.023 | 0.92 (0.84, 0.99) | 0.035 | 0.92 (0.85, 1) | 0.054 |
|  | cg11027354 | OS | 0.98 (0.92, 1.03) | 0.418 | 0.98 (0.92, 1.04) | 0.426 | 0.98 (0.92, 1.04) | 0.435 | 0.92 (0.85, 1.00) | 0.049 | 0.93 (0.86, 1.01) | 0.076 | 0.93 (0.86, 1.01) | 0.097 |
|  | cg12091396 | OS | 1.02 (0.97, 1.08) | 0.394 | 1 (0.95, 1.06) | 0.974 | 1 (0.95, 1.06) | 0.886 | 0.93 (0.84, 1.02) | 0.120 | 0.97 (0.89, 1.06) | 0.512 | 0.99 (0.91, 1.09) | 0.867 |
|  | cg13883256 | OS | 1.00 (0.95, 1.06) | 0.978 | 1.04 (0.99, 1.1) | 0.139 | 1.05 (0.99, 1.11) | 0.086 | 0.96 (0.88, 1.05) | 0.359 | 1.05 (0.96, 1.14) | 0.274 | 1.1 (1.01, 1.19) | 0.033 |
|  | cg15844835 | OS | 0.97 (0.91, 1.02) | 0.245 | 0.97 (0.92, 1.03) | 0.388 | 0.98 (0.92, 1.04) | 0.443 | 0.90 (0.82, 0.98) | 0.021 | 0.93 (0.84, 1.02) | 0.112 | 0.94 (0.86, 1.04) | 0.238 |
|  | cg18421529 | OS | 0.98 (0.93, 1.04) | 0.538 | 1 (0.94, 1.06) | 0.897 | 1 (0.94, 1.06) | 0.939 | 0.92 (0.84, 1.01) | 0.085 | 0.95 (0.86, 1.04) | 0.234 | 0.96 (0.87, 1.05) | 0.36 |
|  | cg19585103 | OS | 0.96 (0.91, 1.02) | 0.214 | 0.96 (0.9, 1.02) | 0.147 | 0.96 (0.9, 1.02) | 0.148 | 0.95 (0.88, 1.03) | 0.224 | 0.94 (0.87, 1.01) | 0.102 | 0.94 (0.87, 1.02) | 0.114 |
|  | cg20717205 | OS | 0.98 (0.92, 1.04) | 0.536 | 1 (0.94, 1.06) | 0.917 | 1 (0.94, 1.06) | 0.891 | 0.88 (0.79, 0.97) | 0.014 | 0.92 (0.83, 1.02) | 0.127 | 0.92 (0.83, 1.02) | 0.101 |
|  | cg22267597 | OS | 0.96 (0.91, 1.02) | 0.213 | 0.95 (0.89, 1.01) | 0.115 | 0.95 (0.9, 1.02) | 0.14 | 0.89 (0.81, 0.97) | 0.010 | 0.91 (0.83, 0.99) | 0.033 | 0.92 (0.84, 1.01) | 0.08 |
|  | cg22346124 | OS | 0.98 (0.92, 1.04) | 0.497 | 0.99 (0.93, 1.05) | 0.753 | 0.99 (0.93, 1.05) | 0.816 | 0.95 (0.87, 1.03) | 0.224 | 0.99 (0.91, 1.08) | 0.793 | 1 (0.92, 1.09) | 0.953 |
|  | cg23835677 | OS | 0.98 (0.92, 1.04) | 0.493 | 0.96 (0.9, 1.01) | 0.138 | 0.96 (0.9, 1.02) | 0.152 | 0.92 (0.83, 1.01) | 0.069 | 0.95 (0.87, 1.03) | 0.211 | 0.96 (0.88, 1.04) | 0.311 |
|  | cg25074185 | OS | 0.92 (0.87, 0.98) | 0.008 | 0.93 (0.87, 0.99) | 0.022 | 0.93 (0.87, 0.99) | 0.021 | 0.89 (0.82, 0.96) | 0.004 | 0.88 (0.81, 0.96) | 0.004 | 0.88 (0.81, 0.96) | 0.004 |
|  | cg26151087 | OS | 1.00 (0.94, 1.05) | 0.903 | 0.99 (0.94, 1.05) | 0.715 | 0.99 (0.94, 1.05) | 0.723 | 0.94 (0.87, 1.02) | 0.148 | 0.95 (0.88, 1.03) | 0.237 | 0.95 (0.88, 1.03) | 0.238 |
|  | cg27525037 | OS | 0.89 (0.84, 0.94) | <0.001 | 0.91 (0.85, 0.96) | 0.001 | 0.91 (0.85, 0.96) | 0.001 | 0.84 (0.77, 0.91) | <0.001 | 0.87 (0.8, 0.95) | 0.002 | 0.87 (0.8, 0.95) | 0.002 |
|  | cg27624313 | OS | 1.05 (0.99, 1.10) | 0.117 | 1.01 (0.96, 1.07) | 0.624 | 1.01 (0.96, 1.07) | 0.629 | 1.04 (0.97, 1.12) | 0.271 | 0.99 (0.92, 1.06) | 0.829 | 0.99 (0.92, 1.06) | 0.822 |

Cells painted with green represent having a p value ≤ 0,05, and those painted with yellow represent having a *p* value above 0.05 but below 0.1. ^1^ Model1 refers to multivariable Cox regression analyses with adjustment of age, sex, TNM stage, treatment, and cancer location. ^2^ Model2 refers to multivariable Cox regression analyses with adjustment of age, sex, TNM stage, treatment, cancer location, and MSI.

### Supplementary Table 6: Equations of prognostic models, their distributions in the validation cohort, and their performance in the original development study

| Study | Outcome | Equation to calculate the prognostic score | PI Distribution  Median (IQR) | Performance in the development study | | |
| --- | --- | --- | --- | --- | --- | --- |
|  |  |  |  | Apparent performance | Internal Performance | External performance |
| Chen et al, 2021^20^ | OS | Risk score= 1.87*cg11621464 + 1.11*cg13565656 + (-1.74*cg18976437) + (2.40*cg20505223) + (-1.97*cg20528583) | 0.2 (-1.8, 2.0) | High-risk group vs low-risk group;  cHR (95%CI): 3.18 (1.82–5.56) aHR (95%CI): 6.17 (2.37-16.0) AUC = 0.78 | cHR (95%CI): 1.75 (1.03–4.17) aHR (95%CI): 11.35 (0.98-130.98)  AUC = 0.73 | NP |
| Gong et al, 2020^13^ | OS | Risk score = 0.09*cg14660573 + −0.04*cg 09353563 + 2.06*cg00110724 | 0.4 (-0.5, 1.1) | log-rank test: p = 0.003, AUC (all sample) = 0.67, AUC (age<73) = 0.59, AUC (age>73) = 0.85, AUC (female) = 0.80, AUC (male) = 0.68 | log-rank test: p = 0.03 | NP |
| Gündert et al, 2019^3^ | OS and DSS | Risk score = (0.18*cg23750514) + (0.15*cg01131395) + (0.26*cg12510999) + (0.19*cg18195165) + (0.3*cg05646575) + (0.24*cg11056055) + (0.21*cg10758824) + (0.31*cg16336556) + (0.22*cg19184885) + (0.12*cg19340296) + (0.22*cg22522598) + (0.29*cg17431888) + (0.15*cg18736676) + (0.23*cg08804626) + (0.19*cg08617020) + (0.31*cg14270346) + (0.07*cg16399624) + (0.22*cg14983135) + (0.24*cg00832644) | 0.3 (-1.6, 1.9) | Change from the lower to the upper quartile; aHR (95%CI), OS: 0.51 (0.41-0.63), DSS: 0.50 (0.38-0.67) | NP | aHR (95%CI), OS: 0.61 (0.45-0.82), DSS: 0.55 (0.38-0.79) |
| Huang et al, 2021^21^ | OS | Risk sore = (0.754*cg21614638) + (1.230*cg21770617) + (0.701*cg12751565 | 0.0 (-1.3, 1.2) | aHR (95%CI): 1.71 (1.06-2.75) AUC, 1-year: 0.61, 3-year: 0.65, 5-year: 0.64 | NP | aHR (95%CI): 1.69 (1.10-2.58) AUC, 3-year: 0.63, 5-year: 0.65, 8-year: 0.63 |
| Huang et al, 2021^21^ | DFS | Risk sore = (0.573*cg21614638) + (0.888*cg21770617) + (0.477*cg12751565) | 0.0 (-0.9, 0.9) | aHR (95%CI): 1.55 (1.08-2.25) AUC, 1-year: 0.61, 3-year: 0.58, 5-year: 0.65 | NP | aHR (95%CI): 1.68 (1.10-2.56) AUC, 3-year: 0.61, 5-year: 0.62, 8-year: 0.61 |
| Li et al, 2021^22^ | OS | Risk score =(-2.1*cg01408654) + (-2.2*cg04035209) + (-3*cg10196720) + (-5*cg10379890) + (2.3*cg11097433) + (4.86*cg14675211) + (2.93*cg15428578) + (-4.7*cg19343464) + (2.7*cg21384402) + (3.7*cg27404023) | -0.4 (-4.2, 4.0) | Log-rank test: p<0.001 aHR (95%CI): 0.96 (1.71-2.89) | NP | NP |
| Wang X et al, 2020^14^ | OS and PFS | Risk score = 0.919*cg03091331 + 0.963*cg06884352 + 0.703*cg07707546 + 0.721*cg08081805 + 0.587*cg21347353 + 0.528*cg25164589 | -0.2 (-1.6, 1.5) | cHR (95%CI), OS: 4.13 (2.10 - 8.01), PFS: 1.99 (1.18-3.34) AUC (95% CI), 3-year: 0.68 (0.59=0.78), 5-year: 0.70 (0.60-0.81) | cHR (95%CI), OS: 2.80 (1.17 - 6.68), PFS: 2.31 (1.08-4.97) | NP |
| Wang Y et al, 2020^16^ | OS | Risk score = (38.52*cg00177496) – (4.13*cg01963906) + (2.574*cg05165940) – (79.32*cg12921795) + (2.31*cg19414598) + (6.061*cg25783173) | 11.2 (-33.6, 44.6) | Log-rank test: p<0.001 AUC: 0.83 | Log-rank test: p<0.001 AUC: 0.79 | NP |
| Xiang et al, 2020^17^ | OS | Risk score = (3.02*cg03017653) + (22.84*cg03977782) + (3.00*cg05417950) + (36.54*cg06250108) + (2.18*cg09893305) + (3.12*cg10414946) + (3.06*cg15170424) + (5.77*cg15639045) + (-9.51*cg15786837) + (22.02*cg 17329249) + (8.12*cg21212956) + (− 23.31*cg24206256) | -10.7 (-34.3, 21.7) | cHR (95%CI): 2.72 (2.10-3.52) aHR (95%CI): 2.87 (2.15-3.82) Log-rank test: p<0.001 AUC: 0.87 | NP | Log-rank test: p<0.001 AUC: 0.68 |
| Yang et al, 2019^18^ | OS | Risk Score=0.12*cg02196655 +1.35*cg03763616+0.73*cg03944089+0.73*cg0611 78 55+0.76*cg07173760-3.96*cg07293947-0.76*cg07509 155+0.58*cg09244244+0.4*cg10451565+0.28*cg1258 2008+1.99*cg13796218+3.6*cg20247048+1.34*cg214 81775+0.42*cg23829949-0.28*cg23964386+0.96*cg 24127989-0.45*cg24674703+0.84*cg24938727 | -0.4 (-3.8, 3.4) | AUC = 0.81, log-rank test: p=0.02 | Log-rank test: p=0.03 | NP |

### Supplementary Table 7: Cox regression analyses for prognostic scores

| **Score (outcome)** | **Quartiles of prognostic score** | **OS** | | | | | | **DFS** | | | | | |
| --- | --- | --- | --- | --- | --- | --- | --- | --- | --- | --- | --- | --- | --- |
|  |  | **Unadjusted model** | | **Model 1** | | **Model 2** | | **Unadjusted model** | | **Model 1** | | **Model 2** | |
|  |  | **cHR (95%CI)** | ***p* value** | **aHR(95%CI)** | ***p* value** | **aHR (95%CI)** | **p value** | **cHR (95%CI)** | ***p* value** | **aHR(95%CI)** | ***p* value** | **aHR(95%CI)** | ***p* value** |
| Chen et al, 2021^20^ (OS) | Q1 | 1 |  | 1 |  | 1 |  | 1 |  | 1 |  |  |  |
|  | Q2 | 1.05 (0.90, 1.23) | 0.522 | 0.96 (0.82, 1.13) | 0.65 | 0.97 (0.82, 1.13) | 0.677 | 1.01 (0.81, 1.25) | 0.936 | 0.93 (0.75, 1.16) | 0.543 | 0.95 (0.76, 1.19) | 0.67 |
|  | Q3 | 1.05 (0.90, 1.23) | 0.508 | 0.99 (0.84, 1.16) | 0.911 | 1 (0.85, 1.17) | 0.955 | 1.03 (0.83, 1.28) | 0.780 | 0.94 (0.76, 1.17) | 0.578 | 0.96 (0.77, 1.19) | 0.69 |
|  | Q4 | 1.12 (0.95, 1.30) | 0.173 | 1 (0.85, 1.17) | 0.982 | 1 (0.85, 1.18) | 0.994 | 1.23 (1.00, 1.52) | 0.048 | 0.99 (0.8, 1.23) | 0.928 | 1 (0.81, 1.24) | 0.986 |
| Gong et al, 2020^13^ (OS) | Q1 | 1 |  | 1 |  | 1 |  | 1 |  | 1 |  |  |  |
|  | Q2 | 1.05 (0.90, 1.24) | 0.520 | 1.01 (0.86, 1.19) | 0.895 | 1.01 (0.86, 1.19) | 0.881 | 1.11 (0.89, 1.38) | 0.352 | 1.05 (0.84, 1.31) | 0.661 | 1.07 (0.86, 1.33) | 0.567 |
|  | Q3 | 1.20 (1.02, 1.40) | 0.025 | 1.14 (0.98, 1.34) | 0.098 | 1.14 (0.97, 1.34) | 0.1 | 1.25 (1.01, 1.54) | 0.040 | 1.18 (0.95, 1.46) | 0.137 | 1.17 (0.94, 1.45) | 0.159 |
|  | Q4 | 1.12 (0.95, 1.31) | 0.176 | 1.18 (1, 1.38) | 0.05 | 1.18 (1, 1.38) | 0.048 | 1.19 (0.96, 1.47) | 0.122 | 1.32 (1.06, 1.65) | 0.012 | 1.33 (1.07, 1.66) | 0.011 |
| Gündert et al, 2019^3^ (OS and DSS) | Q1 | 1 |  | 1 |  | 1 |  | 1 |  | 1 |  |  |  |
|  | Q2 | 0.79 (0.67, 0.94) | 0.008 | 0.73 (0.61, 0.87) | <0.001 | 0.73 (0.61, 0.87) | <0.001 | 0.77 (0.63, 0.95) | 0.014 | 0.72 (0.58, 0.89) | 0.003 | 0.7 (0.57, 0.87) | 0.001 |
|  | Q3 | 0.67 (0.56, 0.81) | <0.001 | 0.83 (0.69, 0.99) | 0.043 | 0.83 (0.69, 0.99) | 0.04 | 0.55 (0.44, 0.70) | <0.001 | 0.81 (0.64, 1.03) | 0.087 | 0.82 (0.64, 1.03) | 0.092 |
|  | Q4 | 0.59 (0.49, 0.70) | <0.001 | 0.73 (0.6, 0.88) | 0.001 | 0.72 (0.6, 0.87) | 0.001 | 0.39 (0.30, 0.50) | <0.001 | 0.6 (0.46, 0.78) | <0.001 | 0.58 (0.44, 0.75) | <0.001 |
| Huang et al, 2021^21^ (OS) | Q1 | 1 |  | 1 |  | 1 |  | 1 |  | 1 |  |  |  |
|  | Q2 | 1.03 (0.88, 1.22) | 0.684 | 1.1 (0.93, 1.3) | 0.255 | 1.1 (0.93, 1.3) | 0.259 | 1.13 (0.89, 1.42) | 0.320 | 1.18 (0.93, 1.49) | 0.181 | 1.18 (0.93, 1.49) | 0.178 |
|  | Q3 | 1.25 (1.06, 1.46) | 0.007 | 1.13 (0.96, 1.33) | 0.141 | 1.13 (0.96, 1.33) | 0.141 | 1.51 (1.21, 1.88) | <0.001 | 1.16 (0.92, 1.45) | 0.206 | 1.16 (0.92, 1.45) | 0.202 |
|  | Q4 | 1.40 (1.20, 1.64) | <0.001 | 1.4 (1.19, 1.65) | <0.001 | 1.4 (1.19, 1.65) | <0.001 | 1.88 (1.51, 2.32) | <0.001 | 1.54 (1.24, 1.93) | <0.001 | 1.55 (1.24, 1.93) | <0.001 |
| Huang et al, 2021^21^ (DFS) | Q1 | 1 |  | 1 |  | 1 |  | 1 |  | 1 |  |  |  |
|  | Q2 | 1.05 (0.89, 1.24) | 0.531 | 1.1 (0.93, 1.3) | 0.25 | 1.1 (0.93, 1.3) | 0.252 | 1.14 (0.90, 1.44) | 0.285 | 1.15 (0.91, 1.46) | 0.253 | 1.15 (0.91, 1.46) | 0.25 |
|  | Q3 | 1.26 (1.07, 1.48) | 0.004 | 1.16 (0.98, 1.36) | 0.077 | 1.16 (0.98, 1.36) | 0.076 | 1.52 (1.22, 1.90) | <0.001 | 1.17 (0.94, 1.47) | 0.166 | 1.18 (0.94, 1.48) | 0.158 |
|  | Q4 | 1.39 (1.19, 1.63) | <0.001 | 1.38 (1.17, 1.62) | <0.001 | 1.37 (1.17, 1.62) | <0.001 | 1.89 (1.52, 2.34) | <0.001 | 1.51 (1.21, 1.88) | <0.001 | 1.5 (1.2, 1.88) | <0.001 |
| Li et al, 2021^22^ (OS) | Q1 | 1 |  | 1 |  | 1 |  | 1 |  | 1 |  |  |  |
|  | Q2 | 1.09 (0.93, 1.27) | 0.296 | 1.15 (0.98, 1.35) | 0.09 | 1.15 (0.98, 1.35) | 0.082 | 0.96 (0.77, 1.19) | 0.697 | 1.06 (0.84, 1.32) | 0.635 | 1.07 (0.86, 1.34) | 0.539 |
|  | Q3 | 1.12 (0.96, 1.32) | 0.150 | 1.13 (0.96, 1.32) | 0.146 | 1.13 (0.96, 1.32) | 0.136 | 1.13 (0.91, 1.39) | 0.278 | 1.14 (0.92, 1.41) | 0.235 | 1.14 (0.92, 1.42) | 0.221 |
|  | Q4 | 1.13 (0.96, 1.32) | 0.141 | 1.11 (0.94, 1.3) | 0.213 | 1.11 (0.94, 1.3) | 0.22 | 1.25 (1.01, 1.54) | 0.036 | 1.15 (0.93, 1.42) | 0.193 | 1.14 (0.92, 1.41) | 0.224 |
| Wang X et al, 2020^14^ (OS and PFS) | Q1 | 1 |  | 1 |  | 1 |  | 1 |  | 1 |  |  |  |
|  | Q2 | 0.98 (0.84, 1.15) | 0.819 | 1.02 (0.87, 1.19) | 0.832 | 1.02 (0.87, 1.19) | 0.825 | 0.83 (0.68, 1.02) | 0.076 | 0.92 (0.75, 1.13) | 0.424 | 0.93 (0.75, 1.14) | 0.479 |
|  | Q3 | 0.98 (0.84, 1.15) | 0.832 | 1.03 (0.88, 1.21) | 0.718 | 1.04 (0.88, 1.22) | 0.674 | 0.87 (0.71, 1.07) | 0.188 | 1.02 (0.83, 1.25) | 0.878 | 1.06 (0.86, 1.3) | 0.61 |
|  | Q4 | 1.00 (0.85, 1.17) | 0.969 | 1.03 (0.88, 1.22) | 0.697 | 1.04 (0.88, 1.23) | 0.621 | 0.73 (0.59, 0.91) | 0.004 | 0.95 (0.76, 1.18) | 0.62 | 0.99 (0.79, 1.23) | 0.927 |
| Wang Y et al, 2020^16^ (OS) | Q1 | 1 |  | 1 |  | 1 |  | 1 |  | 1 |  |  |  |
|  | Q2 | 1.01 (0.86, 1.19) | 0.883 | 0.93 (0.79, 1.09) | 0.358 | 0.93 (0.79, 1.09) | 0.359 | 1.02 (0.83, 1.27) | 0.825 | 0.94 (0.75, 1.17) | 0.554 | 0.93 (0.74, 1.16) | 0.499 |
|  | Q3 | 1.20 (1.03, 1.41) | 0.022 | 1.14 (0.97, 1.34) | 0.104 | 1.14 (0.97, 1.34) | 0.101 | 1.19 (0.97, 1.48) | 0.100 | 1.11 (0.89, 1.37) | 0.351 | 1.11 (0.89, 1.37) | 0.357 |
|  | Q4 | 1.28 (1.09, 1.49) | 0.002 | 1.18 (1.01, 1.39) | 0.035 | 1.19 (1.01, 1.39) | 0.033 | 1.25 (1.02, 1.55) | 0.034 | 1.13 (0.91, 1.4) | 0.267 | 1.13 (0.92, 1.4) | 0.251 |
| Xiang et al, 2020^17^ (OS) | Q1 | 1 |  | 1 |  | 1 |  | 1 |  | 1 |  |  |  |
|  | Q2 | 0.99 (0.85, 1.15) | 0.888 | 0.94 (0.81, 1.1) | 0.443 | 0.94 (0.8, 1.1) | 0.431 | 0.91 (0.75, 1.12) | 0.384 | 0.83 (0.68, 1.02) | 0.08 | 0.82 (0.66, 1.01) | 0.057 |
|  | Q3 | 0.89 (0.76, 1.04) | 0.131 | 0.97 (0.83, 1.14) | 0.735 | 0.97 (0.83, 1.14) | 0.711 | 0.86 (0.70, 1.06) | 0.160 | 0.91 (0.74, 1.12) | 0.383 | 0.89 (0.72, 1.1) | 0.295 |
|  | Q4 | 0.84 (0.72, 0.98) | 0.029 | 0.86 (0.74, 1.01) | 0.071 | 0.86 (0.73, 1.01) | 0.063 | 0.73 (0.59, 0.91) | 0.004 | 0.72 (0.58, 0.9) | 0.004 | 0.71 (0.57, 0.88) | 0.002 |
| Yang et al, 2019^18^ (OS) | Q1 | 1 |  | 1 |  | 1 |  | 1 |  | 1 |  |  |  |
|  | Q2 | 0.94 (0.81, 1.11) | 0.482 | 0.87 (0.74, 1.02) | 0.085 | 0.87 (0.74, 1.02) | 0.085 | 0.92 (0.74, 1.14) | 0.446 | 0.8 (0.65, 1) | 0.052 | 0.81 (0.65, 1) | 0.055 |
|  | Q3 | 1.09 (0.93, 1.27) | 0.305 | 1.05 (0.9, 1.23) | 0.544 | 1.05 (0.9, 1.23) | 0.535 | 1.08 (0.87, 1.33) | 0.483 | 0.98 (0.8, 1.21) | 0.869 | 0.99 (0.8, 1.22) | 0.934 |
|  | Q4 | 1.07 (0.92, 1.25) | 0.396 | 1.03 (0.88, 1.21) | 0.691 | 1.04 (0.88, 1.22) | 0.657 | 1.02 (0.83, 1.26) | 0.827 | 0.9 (0.73, 1.11) | 0.334 | 0.92 (0.74, 1.14) | 0.443 |

### Supplementary Table 8: Time-dependent AUC for all scores

|  |  | **OS** | | **DFS** | |
| --- | --- | --- | --- | --- | --- |
|  | **Times** | **Continuous score** | **Quartiles of score** | **Continuous score** | **Quartiles of score** |
| **Scores significant in multicox** | |  |  |  |  |
| Gong et al, 2020^13^ | 1 year | 0.57 (0.52, 0.62) | 0.55 (0.51, 0.6) | 0.57 (0.53, 0.61) | 0.56 (0.52, 0.59) |
|  | 3 years | 0.56 (0.53, 0.58) | 0.55 (0.52, 0.58) | 0.54 (0.51, 0.57) | 0.53 (0.5, 0.56) |
|  | 5 years | 0.54 (0.52, 0.57) | 0.53 (0.51, 0.56) | 0.54 (0.51, 0.57) | 0.53 (0.5, 0.55) |
|  | 8 years | 0.53 (0.51, 0.56) | 0.53 (0.5, 0.55) | 0.54 (0.51, 0.57) | 0.53 (0.5, 0.56) |
| Gündert et al, 2019^3^ | 1 year | 0.64 (0.59, 0.69) | 0.62 (0.57, 0.67) | 0.67 (0.63, 0.71) | 0.65 (0.61, 0.68) |
|  | 3 years | 0.63 (0.61, 0.66) | 0.6 (0.57, 0.63) | 0.64 (0.61, 0.67) | 0.62 (0.6, 0.65) |
|  | 5 years | 0.6 (0.58, 0.63) | 0.58 (0.56, 0.61) | 0.63 (0.6, 0.66) | 0.62 (0.59, 0.65) |
|  | 8 years | 0.59 (0.57, 0.62) | 0.58 (0.55, 0.6) | 0.62 (0.58, 0.65) | 0.61 (0.57, 0.64) |
| Huang et al, 2021 (OS)^21^ | 1 year | 0.57 (0.52, 0.62) | 0.56 (0.52, 0.61) | 0.6 (0.56, 0.64) | 0.59 (0.55, 0.62) |
|  | 3 years | 0.56 (0.53, 0.59) | 0.55 (0.52, 0.58) | 0.58 (0.55, 0.61) | 0.57 (0.55, 0.6) |
|  | 5 years | 0.56 (0.54, 0.59) | 0.56 (0.54, 0.58) | 0.59 (0.56, 0.62) | 0.58 (0.56, 0.61) |
|  | 8 years | 0.55 (0.52, 0.57) | 0.54 (0.52, 0.57) | 0.58 (0.55, 0.61) | 0.58 (0.55, 0.61) |
| Huang et al, 2021 (DFS)^21^ | 1 year | 0.57 (0.52, 0.62) | 0.56 (0.52, 0.61) | 0.6 (0.56, 0.64) | 0.59 (0.55, 0.62) |
|  | 3 years | 0.56 (0.53, 0.59) | 0.55 (0.52, 0.58) | 0.58 (0.55, 0.61) | 0.57 (0.55, 0.6) |
|  | 5 years | 0.56 (0.54, 0.59) | 0.56 (0.53, 0.58) | 0.59 (0.56, 0.61) | 0.58 (0.55, 0.6) |
|  | 8 years | 0.55 (0.52, 0.57) | 0.54 (0.52, 0.57) | 0.58 (0.55, 0.61) | 0.58 (0.55, 0.61) |
| Wang Y et al, 2020^16^ | 1 year | 0.59 (0.54, 0.64) | 0.58 (0.53, 0.62) | 0.57 (0.53, 0.61) | 0.55 (0.51, 0.59) |
|  | 3 years | 0.56 (0.54, 0.59) | 0.56 (0.54, 0.59) | 0.55 (0.52, 0.57) | 0.54 (0.52, 0.57) |
|  | 5 years | 0.55 (0.53, 0.58) | 0.55 (0.53, 0.58) | 0.54 (0.52, 0.57) | 0.54 (0.52, 0.57) |
|  | 8 years | 0.55 (0.53, 0.57) | 0.55 (0.53, 0.57) | 0.55 (0.52, 0.58) | 0.54 (0.52, 0.57) |
| **Scores not significant in multicox** | |  |  |  |  |
| Chen et al, 2021^20^ | 1 year | 0.53 (0.5, 0.55) | 0.55 (0.5, 0.6) | 0.53 (0.5, 0.55) | 0.53 (0.49, 0.57) |
|  | 3 years | 0.55 (0.5, 0.6) | 0.54 (0.51, 0.57) | 0.53 (0.49, 0.57) | 0.53 (0.5, 0.56) |
|  | 5 years | 0.54 (0.51, 0.57) | 0.52 (0.5, 0.55) | 0.53 (0.5, 0.56) | 0.53 (0.5, 0.55) |
|  | 8 years | 0.53 (0.5, 0.55) | 0.52 (0.5, 0.54) | 0.53 (0.5, 0.56) | 0.51 (0.49, 0.54) |
| Li et al, 2021^22^ | 1 year | 0.51 (0.49, 0.54) | 0.51 (0.47, 0.56) | 0.54 (0.51, 0.57) | 0.52 (0.48, 0.56) |
|  | 3 years | 0.52 (0.48, 0.57) | 0.5 (0.47, 0.53) | 0.53 (0.49, 0.57) | 0.52 (0.49, 0.55) |
|  | 5 years | 0.51 (0.48, 0.54) | 0.5 (0.48, 0.53) | 0.53 (0.5, 0.55) | 0.54 (0.51, 0.56) |
|  | 8 years | 0.51 (0.48, 0.53) | 0.52 (0.5, 0.54) | 0.54 (0.51, 0.56) | 0.54 (0.51, 0.56) |
| Wang X et al, 2020^14^ | 1 year | 0.51 (0.49, 0.54) | 0.53 (0.49, 0.58) | 0.54 (0.51, 0.57) | 0.51 (0.47, 0.55) |
|  | 3 years | 0.5 (0.45, 0.54) | 0.5 (0.47, 0.53) | 0.51 (0.47, 0.55) | 0.53 (0.51, 0.56) |
|  | 5 years | 0.51 (0.48, 0.54) | 0.51 (0.48, 0.53) | 0.53 (0.5, 0.56) | 0.54 (0.51, 0.57) |
|  | 8 years | 0.51 (0.49, 0.54) | 0.52 (0.49, 0.54) | 0.54 (0.52, 0.57) | 0.53 (0.5, 0.56) |
| Xiang et al, 2020^17^ | 1 year | 0.54 (0.51, 0.56) | 0.56 (0.51, 0.61) | 0.54 (0.51, 0.57) | 0.54 (0.5, 0.58) |
|  | 3 years | 0.55 (0.51, 0.6) | 0.53 (0.5, 0.56) | 0.54 (0.5, 0.58) | 0.52 (0.5, 0.55) |
|  | 5 years | 0.53 (0.5, 0.56) | 0.53 (0.51, 0.55) | 0.53 (0.5, 0.56) | 0.53 (0.5, 0.55) |
|  | 8 years | 0.53 (0.51, 0.56) | 0.54 (0.51, 0.56) | 0.53 (0.51, 0.56) | 0.55 (0.52, 0.58) |
| Yang et al, 2019^18^ | 1 year | 0.51 (0.48, 0.53) | 0.51 (0.47, 0.56) | 0.53 (0.5, 0.56) | 0.54 (0.5, 0.57) |
|  | 3 years | 0.51 (0.46, 0.56) | 0.51 (0.48, 0.54) | 0.54 (0.5, 0.58) | 0.53 (0.5, 0.56) |
|  | 5 years | 0.51 (0.48, 0.54) | 0.51 (0.49, 0.54) | 0.53 (0.51, 0.56) | 0.53 (0.5, 0.55) |
|  | 8 years | 0.51 (0.49, 0.54) | 0.51 (0.49, 0.53) | 0.53 (0.51, 0.56) | 0.53 (0.5, 0.56) |

### Supplementary Table 9: Stratified analyses for the AUC at mean follow-up time of prognostic scores

| Score | Group | OS | | DFS | |
| --- | --- | --- | --- | --- | --- |
|  |  | **Sample size** | **AUC (95%CI)** | **Sample Size** | **AUC (95%CI)** |
|  | **Age (year)** | | | | |
| Gong et al, 2020 | <70 | 1157 | 0.53 (0.49, 0.56) | 1149 | 0.53 (0.49, 0.56) |
|  | >=70 | 1153 | 0.55 (0.52, 0.58) | 1141 | 0.54 (0.51, 0.58) |
| Gündert et al, 2019 | <70 | 872 | 0.59 (0.55, 0.63) | 866 | 0.63 (0.58, 0.67) |
|  | >=70 | 866 | 0.59 (0.55, 0.63) | 858 | 0.63 (0.58, 0.67) |
| Huang et al, 2021 (OS) | <70 | 1157 | 0.54 (0.51, 0.58) | 1149 | 0.57 (0.53, 0.61) |
|  | >=70 | 1153 | 0.56 (0.52, 0.59) | 1141 | 0.6 (0.56, 0.63) |
| Huang et al, 2021 (DFS) | <70 | 1157 | 0.54 (0.51, 0.58) | 1149 | 0.57 (0.53, 0.61) |
|  | >=70 | 1153 | 0.56 (0.52, 0.59) | 1141 | 0.6 (0.56, 0.63) |
| Wang Y et al, 2020 | <70 | 1157 | 0.53 (0.5, 0.57) | 1149 | 0.55 (0.51, 0.58) |
|  | >=70 | 1153 | 0.54 (0.5, 0.57) | 1141 | 0.53 (0.49, 0.57) |
|  | **Gender** | | | | |
| Gong et al, 2020 | Female | 964 | 0.53 (0.49, 0.57) | 955 | 0.53 (0.49, 0.58) |
|  | Male | 1346 | 0.54 (0.51, 0.57) | 1335 | 0.54 (0.5, 0.58) |
| Gündert et al, 2019 | Female | 699 | 0.61 (0.57, 0.66) | 692 | 0.64 (0.59, 0.68) |
|  | Male | 1039 | 0.58 (0.54, 0.61) | 1032 | 0.61 (0.58, 0.65) |
| Huang et al, 2021 (OS) | Female | 964 | 0.55 (0.51, 0.58) | 955 | 0.59 (0.54, 0.63) |
|  | Male | 1346 | 0.54 (0.51, 0.57) | 1335 | 0.58 (0.54, 0.61) |
| Huang et al, 2021 (DFS) | Female | 964 | 0.55 (0.51, 0.58) | 955 | 0.59 (0.54, 0.63) |
|  | Male | 1346 | 0.54 (0.51, 0.57) | 1335 | 0.58 (0.54, 0.61) |
| Wang Y et al, 2020 | Female | 964 | 0.52 (0.48, 0.56) | 955 | 0.53 (0.48, 0.57) |
|  | Male | 1346 | 0.56 (0.53, 0.6) | 1335 | 0.55 (0.51, 0.58) |
|  | **TNM stage** | | | | |
| Gong et al, 2020 | I & II | 1185 | 0.53 (0.49, 0.56) | 1178 | 0.54 (0.49, 0.59) |
|  | III | 747 | 0.54 (0.5, 0.58) | 737 | 0.54 (0.49, 0.59) |
|  | IV | 336 | 0.56 (0.5, 0.62) | 333 | 0.56 (0.5, 0.62) |
| Gündert et al, 2019 | I & II | 850 | 0.54 (0.5, 0.58) | 845 | 0.6 (0.54, 0.67) |
|  | III | 519 | 0.57 (0.52, 0.62) | 513 | 0.58 (0.53, 0.64) |
|  | IV | 335 | 0.62 (0.56, 0.68) | 332 | 0.61 (0.55, 0.67) |
| Huang et al, 2021 (OS) | I & II | 1185 | 0.52 (0.49, 0.56) | 1178 | 0.54 (0.48, 0.59) |
|  | III | 747 | 0.48 (0.44, 0.53) | 737 | 0.53 (0.48, 0.57) |
|  | IV | 336 | 0.59 (0.53, 0.65) | 333 | 0.59 (0.53, 0.65) |
| Huang et al, 2021 (DFS) | I & II | 1185 | 0.52 (0.49, 0.56) | 1178 | 0.54 (0.48, 0.59) |
|  | III | 747 | 0.48 (0.44, 0.53) | 737 | 0.53 (0.48, 0.57) |
|  | IV | 336 | 0.59 (0.52, 0.65) | 333 | 0.59 (0.52, 0.65) |
| Wang Y et al, 2020 | I & II | 1185 | 0.54 (0.5, 0.58) | 1178 | 0.52 (0.46, 0.58) |
|  | III | 747 | 0.54 (0.5, 0.58) | 737 | 0.53 (0.48, 0.58) |
|  | IV | 336 | 0.53 (0.47, 0.59) | 333 | 0.53 (0.47, 0.59) |
|  | **Cancer location** | | | | |
| Gong et al, 2020 | Distal colon | 636 | 0.51 (0.46, 0.56) | 632 | 0.51 (0.46, 0.56) |
|  | Proximal colon | 851 | 0.56 (0.53, 0.6) | 844 | 0.58 (0.53, 0.62) |
|  | Rectum | 822 | 0.51 (0.47, 0.55) | 813 | 0.52 (0.47, 0.56) |
| Gündert et al, 2019 | Distal colon | 476 | 0.6 (0.55, 0.65) | 473 | 0.64 (0.59, 0.7) |
|  | Proximal colon | 617 | 0.61 (0.57, 0.66) | 613 | 0.66 (0.61, 0.71) |
|  | Rectum | 822 | 0.57 (0.53, 0.61) | 813 | 0.6 (0.56, 0.65) |
| Huang et al, 2021 (OS) | Distal colon | 636 | 0.55 (0.5, 0.59) | 632 | 0.56 (0.51, 0.61) |
|  | Proximal colon | 851 | 0.58 (0.54, 0.62) | 844 | 0.61 (0.56, 0.66) |
|  | Rectum | 822 | 0.51 (0.47, 0.56) | 813 | 0.56 (0.52, 0.61) |
| Huang et al, 2021 (DFS) | Distal colon | 636 | 0.55 (0.5, 0.59) | 632 | 0.56 (0.51, 0.61) |
|  | Proximal colon | 851 | 0.58 (0.54, 0.62) | 844 | 0.61 (0.56, 0.66) |
|  | Rectum | 822 | 0.51 (0.47, 0.55) | 813 | 0.56 (0.52, 0.61) |
| Wang Y et al, 2020 | Distal colon | 636 | 0.55 (0.51, 0.6) | 632 | 0.56 (0.5, 0.61) |
|  | Proximal colon | 851 | 0.56 (0.52, 0.6) | 844 | 0.54 (0.49, 0.58) |
|  | Rectum | 822 | 0.53 (0.49, 0.57) | 813 | 0.53 (0.48, 0.57) |
|  | **Treatment** | | | | |
| Gong et al, 2020 | No | 1217 | 0.54 (0.51, 0.58) | 1211 | 0.57 (0.52, 0.61) |
|  | Yes | 1087 | 0.53 (0.49, 0.56) | 1076 | 0.52 (0.49, 0.56) |
| Gündert et al, 2019 | No | 885 | 0.57 (0.53, 0.61) | 882 | 0.63 (0.58, 0.68) |
|  | Yes | 848 | 0.58 (0.54, 0.62) | 839 | 0.58 (0.54, 0.62) |
| Huang et al, 2021 (OS) | No | 1217 | 0.54 (0.5, 0.57) | 1211 | 0.57 (0.52, 0.62) |
|  | Yes | 1087 | 0.53 (0.49, 0.56) | 1076 | 0.54 (0.5, 0.57) |
| Huang et al, 2021 (DFS) | No | 1217 | 0.53 (0.5, 0.57) | 1211 | 0.57 (0.52, 0.62) |
|  | Yes | 1087 | 0.53 (0.49, 0.56) | 1076 | 0.54 (0.5, 0.57) |
| Wang Y et al, 2020 | No | 1217 | 0.55 (0.52, 0.59) | 1211 | 0.53 (0.48, 0.57) |
|  | Yes | 1087 | 0.54 (0.51, 0.58) | 1076 | 0.54 (0.51, 0.58) |
|  | **MSI status** | | | | |
| Gong et al, 2020 | MSI | 223 | 0.54 (0.46, 0.62) | 221 | 0.57 (0.46, 0.69) |
|  | MSS | 1769 | 0.54 (0.51, 0.56) | 1751 | 0.54 (0.51, 0.57) |
| Gündert et al, 2019 | MSI | 153 | 0.57 (0.47, 0.66) | 151 | 0.53 (0.38, 0.67) |
|  | MSS | 1303 | 0.6 (0.57, 0.63) | 1291 | 0.65 (0.61, 0.68) |
| Huang et al, 2021 (OS) | MSI | 223 | 0.54 (0.46, 0.62) | 221 | 0.55 (0.42, 0.68) |
|  | MSS | 1769 | 0.54 (0.51, 0.56) | 1751 | 0.57 (0.54, 0.6) |
| Huang et al, 2021 (DFS) | MSI | 223 | 0.54 (0.46, 0.62) | 221 | 0.55 (0.42, 0.68) |
|  | MSS | 1769 | 0.54 (0.51, 0.56) | 1751 | 0.57 (0.54, 0.6) |
| Wang Y et al, 2020 | MSI | 223 | 0.6 (0.52, 0.68) | 221 | 0.59 (0.46, 0.71) |
|  | MSS | 1769 | 0.54 (0.52, 0.57) | 1751 | 0.54 (0.51, 0.57) |
|  | **BRAF mutation** | | | | |
| Gong et al, 2020 | Yes | 166 | 0.57 (0.48, 0.66) | 163 | 0.63 (0.52, 0.73) |
|  | No | 1914 | 0.54 (0.51, 0.56) | 1897 | 0.54 (0.51, 0.57) |
| Gündert et al, 2019 | Yes | 123 | 0.68 (0.59, 0.77) | 120 | 0.72 (0.61, 0.82) |
|  | No | 1444 | 0.58 (0.55, 0.61) | 1433 | 0.62 (0.58, 0.65) |
| Huang et al, 2021 (OS) | Yes | 166 | 0.66 (0.58, 0.74) | 163 | 0.74 (0.66, 0.83) |
|  | No | 1914 | 0.54 (0.51, 0.56) | 1897 | 0.57 (0.54, 0.6) |
| Huang et al, 2021 (DFS) | Yes | 166 | 0.66 (0.58, 0.74) | 163 | 0.75 (0.66, 0.83) |
|  | No | 1914 | 0.54 (0.51, 0.56) | 1897 | 0.57 (0.54, 0.6) |
| Wang Y et al, 2020 | Yes | 166 | 0.59 (0.51, 0.68) | 163 | 0.54 (0.43, 0.65) |
|  | No | 1914 | 0.54 (0.51, 0.56) | 1897 | 0.53 (0.5, 0.56) |
|  | **KRAS mutation** | | | | |
| Gong et al, 2020 | Yes | 693 | 0.55 (0.51, 0.59) | 688 | 0.56 (0.51, 0.6) |
|  | No | 1388 | 0.53 (0.5, 0.56) | 1374 | 0.54 (0.5, 0.57) |
| Gündert et al, 2019 | Yes | 693 | 0.53 (0.48, 0.57) | 688 | 0.56 (0.51, 0.61) |
|  | No | 1388 | 0.56 (0.53, 0.59) | 1374 | 0.59 (0.56, 0.63) |
| Huang et al, 2021 (OS) | Yes | 693 | 0.52 (0.48, 0.57) | 688 | 0.56 (0.51, 0.61) |
|  | No | 1388 | 0.56 (0.53, 0.59) | 1374 | 0.59 (0.56, 0.63) |
| Huang et al, 2021 (DFS) | Yes | 693 | 0.53 (0.49, 0.58) | 688 | 0.54 (0.49, 0.58) |
|  | No | 1388 | 0.55 (0.52, 0.58) | 1374 | 0.53 (0.5, 0.57) |
| Wang Y et al, 2020 | Yes | 532 | 0.6 (0.55, 0.65) | 530 | 0.63 (0.58, 0.69) |
|  | No | 1062 | 0.58 (0.55, 0.62) | 1050 | 0.63 (0.59, 0.67) |
|  | **CIMP phenotype** | | | | |
| Gong et al, 2020 | high | 388 | 0.59 (0.53, 0.65) | 382 | 0.62 (0.55, 0.69) |
|  | low | 1811 | 0.52 (0.5, 0.55) | 1797 | 0.53 (0.5, 0.56) |
| Gündert et al, 2019 | high | 388 | 0.57 (0.52, 0.63) | 382 | 0.68 (0.61, 0.75) |
|  | low | 1811 | 0.54 (0.51, 0.57) | 1797 | 0.56 (0.53, 0.59) |
| Huang et al, 2021 (OS) | high | 388 | 0.58 (0.52, 0.63) | 382 | 0.68 (0.61, 0.75) |
|  | low | 1811 | 0.54 (0.51, 0.57) | 1797 | 0.56 (0.53, 0.59) |
| Huang et al, 2021 (DFS) | high | 388 | 0.57 (0.51, 0.63) | 382 | 0.56 (0.48, 0.63) |
|  | low | 1811 | 0.55 (0.52, 0.57) | 1797 | 0.54 (0.51, 0.57) |
| Wang Y et al, 2020 | high | 295 | 0.58 (0.52, 0.65) | 290 | 0.58 (0.5, 0.66) |
|  | low | 1346 | 0.59 (0.56, 0.62) | 1337 | 0.64 (0.61, 0.68) |

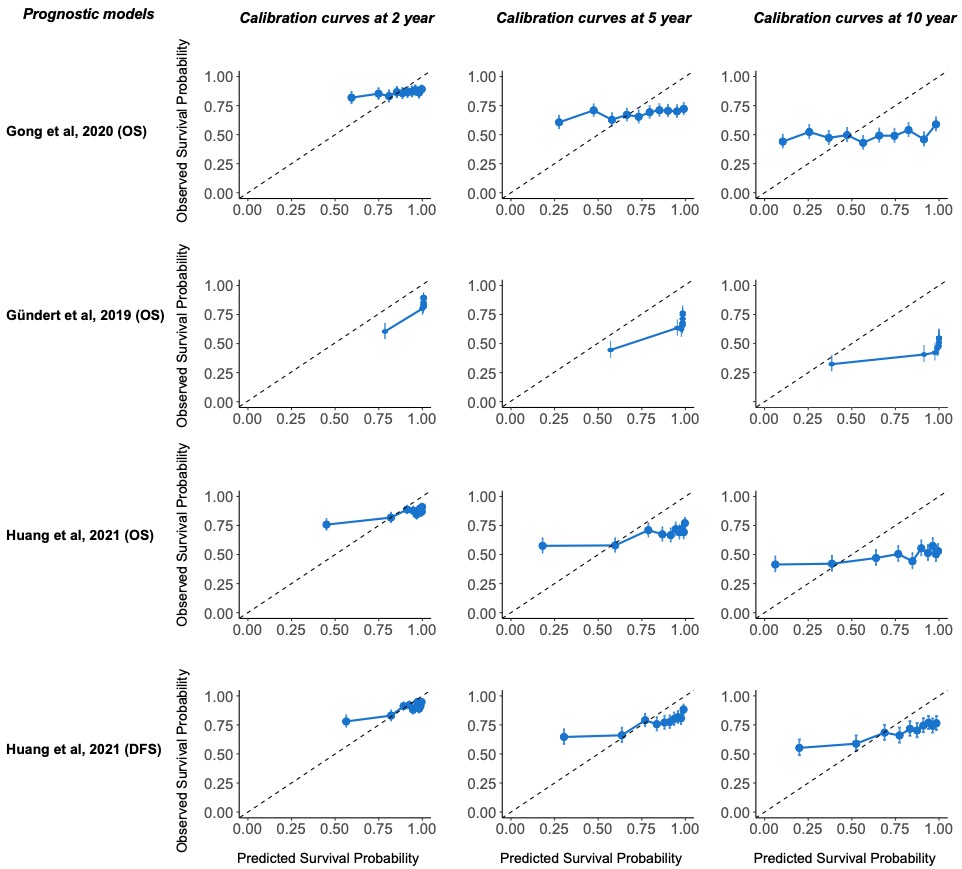

### Supplementary Figure 3: Calibration curves for prognostic models

The baseline survival for each prognostic model was recalibrated. The calibration curves for the prognostic model developed by Wang Y et al, 2020 was unable to show, because the recalibrated baseline survival equaled to 1 at each time point.

### Supplementary Table 10: *P* values for the likelihood ratio test comparing models based on clinical variables only and models based on clinical variables added with prognostic scores^1^

| **Imputation dataset** | **OS** | | | | | **DFS** | | | | |
| --- | --- | --- | --- | --- | --- | --- | --- | --- | --- | --- |
|  | **Huang et al, 2021 (OS)** | **Huang et al, 2021 (DFS)** | **Wang Y et al, 2020** | **Gong et al, 2020** | **Gündert et al, 2019** | **Huang et al, 2021 (OS)** | **Huang et al, 2021 (DFS)** | **Wang Y et al, 2020** | **Gong et al, 2020** | **Gündert et al, 2019** |
| #1 | <0.001 | <0.001 | 0.212 | 0.01 | <0.001 | <0.001 | <0.001 | 0.201 | 0.003 | <0.001 |
| #2 | <0.001 | <0.001 | 0.188 | 0.008 | <0.001 | <0.001 | <0.001 | 0.155 | 0.003 | <0.001 |
| #3 | <0.001 | <0.001 | 0.188 | 0.016 | <0.001 | <0.001 | <0.001 | 0.174 | 0.005 | <0.001 |
| #4 | <0.001 | <0.001 | 0.161 | 0.008 | <0.001 | <0.001 | <0.001 | 0.147 | 0.002 | <0.001 |
| #5 | <0.001 | <0.001 | 0.152 | 0.007 | <0.001 | <0.001 | <0.001 | 0.142 | 0.001 | <0.001 |
| #6 | <0.001 | <0.001 | 0.142 | 0.01 | <0.001 | <0.001 | <0.001 | 0.119 | 0.003 | <0.001 |
| #7 | <0.001 | <0.001 | 0.16 | 0.011 | <0.001 | <0.001 | <0.001 | 0.142 | 0.004 | <0.001 |
| #8 | <0.001 | <0.001 | 0.175 | 0.011 | <0.001 | <0.001 | <0.001 | 0.138 | 0.003 | <0.001 |
| #9 | <0.001 | <0.001 | 0.157 | 0.008 | <0.001 | <0.001 | <0.001 | 0.145 | 0.002 | <0.001 |
| #10 | <0.001 | <0.001 | 0.117 | 0.013 | <0.001 | <0.001 | <0.001 | 0.097 | 0.004 | <0.001 |
| #11 | <0.001 | <0.001 | 0.163 | 0.01 | <0.001 | <0.001 | <0.001 | 0.154 | 0.003 | <0.001 |
| #12 | <0.001 | <0.001 | 0.112 | 0.011 | <0.001 | <0.001 | <0.001 | 0.094 | 0.003 | <0.001 |
| #13 | <0.001 | <0.001 | 0.126 | 0.013 | <0.001 | <0.001 | <0.001 | 0.110 | 0.005 | <0.001 |
| #14 | <0.001 | <0.001 | 0.123 | 0.017 | <0.001 | <0.001 | <0.001 | 0.105 | 0.006 | <0.001 |
| #15 | <0.001 | <0.001 | 0.159 | 0.012 | <0.001 | <0.001 | <0.001 | 0.127 | 0.005 | <0.001 |
| #16 | <0.001 | <0.001 | 0.124 | 0.014 | <0.001 | <0.001 | <0.001 | 0.105 | 0.003 | <0.001 |
| #17 | <0.001 | <0.001 | 0.165 | 0.008 | <0.001 | <0.001 | <0.001 | 0.152 | 0.002 | <0.001 |
| #18 | <0.001 | <0.001 | 0.121 | 0.01 | <0.001 | <0.001 | <0.001 | 0.106 | 0.003 | <0.001 |
| #19 | <0.001 | <0.001 | 0.121 | 0.014 | <0.001 | <0.001 | <0.001 | 0.108 | 0.006 | <0.001 |
| #20 | <0.001 | <0.001 | 0.162 | 0.008 | <0.001 | <0.001 | <0.001 | 0.128 | 0.002 | <0.001 |

^1^ The two models compared are: 1) Outcome ~ age + sex + TNM stage, and 2) Outcome ~ age + sex + TNM stage + prognostic model.

### Supplementary Table 11: Differences in AUC comparing models based on clinical variables only and that added with prognostic scores

|  |  | **AUC difference^1^** | |
| --- | --- | --- | --- |
|  | **Times** | **OS** | **DFS** |
| Gong et al, 2020 | 1 year | 0.002 (-0.001, 0.005) | 0.004 (-0.001, 0.009) |
|  | 3 years | 0.002 (-0.001, 0.004) | 0.001 (-0.002, 0.005) |
|  | 5 years | 0.001 (-0.001, 0.004) | 0 (-0.003, 0.004) |
|  | 8 years | 0.001 (-0.001, 0.004) | 0.002 (-0.002, 0.006) |
| Gündert et al, 2019 | 1 year | -0.001 (-0.008, 0.007) | 0.009 (0.001, 0.017) |
|  | 3 years | 0.003 (-0.002, 0.008) | 0.006 (0, 0.012) |
|  | 5 years | 0 (-0.004, 0.005) | 0.005 (-0.001, 0.01) |
|  | 8 years | 0 (-0.005, 0.004) | 0.003 (-0.003, 0.009) |
| Huang et al, 2021 (OS) | 1 year | -0.002 (-0.007, 0.003) | 0.003 (-0.003, 0.009) |
|  | 3 years | 0.001 (-0.003, 0.004) | -0.001 (-0.005, 0.004) |
|  | 5 years | 0.001 (-0.002, 0.005) | 0 (-0.004, 0.005) |
|  | 8 years | -0.001 (-0.005, 0.002) | 0 (-0.005, 0.005) |
| Huang et al, 2021 (DFS) | 1 year | -0.002 (-0.007, 0.003) | 0.003 (-0.003, 0.009) |
|  | 3 years | 0.001 (-0.003, 0.004) | -0.001 (-0.005, 0.004) |
|  | 5 years | 0.002 (-0.002, 0.005) | 0 (-0.004, 0.005) |
|  | 8 years | -0.001 (-0.005, 0.002) | 0 (-0.004, 0.005) |
| Wang Y et al, 2020 | 1 year | 0.003 (0.001, 0.005) | 0.003 (0.001, 0.006) |
|  | 3 years | 0.002 (0.001, 0.003) | 0.002 (0, 0.003) |
|  | 5 years | 0 (-0.001, 0.001) | 0 (-0.002, 0.002) |
|  | 8 years | 0 (-0.001, 0.001) | 0 (-0.002, 0.002) |

^1^Compare the AUC score for the following two models: 1) Outcome ~ age + sex + TNM stage, and 2) Outcome ~ age + sex + TNM stage + continuous prognostic score; AUC difference = AUC (2) – AUC (1).

### Supplementary Table 12: PROBAST risk of bias for all prognostic models to be validated

|  | **Chen et al, 2021** | | **Gong et al, 2020** | | **Gündert et al, 2019** | | **Huang et al, 2021** | | **Li et al, 2021** | | **Wang X et al, 2020** | | **Wang Y et al, 2020** | | **Xiang et al, 2020** |  | **Yang et al, 2019** |  |
| --- | --- | --- | --- | --- | --- | --- | --- | --- | --- | --- | --- | --- | --- | --- | --- | --- | --- | --- |
|  | **Answer** | **Rationale** | **Answer** | **Rationale** | **Answer** | **Rationale** | **Answer** | **Rationale** | **Answer** | **Rationale** | **Answer** | **Rationale** | **Answer** | **Rationale** | **Answer** | **Rationale** | **Answer** | **Rationale** |
| **DOMAIN 1: Participants** |  |  |  |  |  |  |  |  |  |  |  |  |  |  |  |  |  |  |
| 1.1 Were appropriate data sources used, e.g. cohort, RCT or nested case-control study data? | Probably yes | Data were retrieved from the TCGA database | Probably yes | Data were retrieved from the TCGA database | Yes | Data were retrieved from the prospective database | Yes | Data were retrieved from TCGA and ArrayExpress databases | Yes | Data were retrieved from the TCGA database | Yes | Data were retrieved from the TCGA database | Yes | Data were retrieved from the TCGA database | Yes | Data were retrieved from the TCGA database | Yes | Data were retrieved from the TCGA database |
| 1.2 Were all inclusions and exclusions of participants appropriate? | No information | 182 stage II/III CRC samples with Illumina 450K methylation information were download, but more detailed inclusion/exclusion criteria were not provided. | Probably no | Only patients with complete clinical data were selected | Probably no | Patients who had received neoadjuvant therapy were excluded | Probably no | Only patients with both DNA methylation data and corresponding RNA!seq expression data were included | Probably no | Only patients with both methylation and transcriptome information were included | Probably no | Patients who had received neoadjuvant therapy were excluded | No information | Detailed inclusion/exclusion criteria were not provided. | No information | Detailed inclusion/exclusion criteria were not provided. | No information | Detailed inclusion/exclusion criteria were not provided. |
| ***Overall ROB Domain 1*** | ***Unclear risk of bias*** | | ***High risk of bias*** | | ***High risk of bias*** | | ***High risk of bias*** | | ***High risk of bias*** | | ***High risk of bias*** | | ***Unclear risk of bias*** | | ***Unclear risk of bias*** | | ***Unclear risk of bias*** | |
| **DOMAIN 2: Predictors** |  |  |  |  |  |  |  |  |  |  |  |  |  |  |  |  |  |  |
| 2.1 Were predictors defined and assessed in a similar way for all participants? | Yes | Illumina 450 K Methylation Beadchip | Yes | Illumina 450 K Methylation Beadchip | Yes | Illumina 450 K Methylation Beadchip | Yes | DNA methylation data and RNA seq expression data were obtained with the Illumina Infinium Human Methylation 450 platform and the Illumina HiSeq 2000 RNA Sequencing platform. | No information | Methods used to measure methylation information were not reported, which could be Illumina 450 K Methylation Beadchip or Illumina 270 K Methylation Beadchip, or both. | Yes | Illumina 450 K Methylation Beadchip | Yes | Illumina 450 K Methylation Beadchip | Yes | Illumina 450 K Methylation Beadchip | Yes | Illumina 450 K Methylation Beadchip |
| 2.2 Were predictor assessments made without knowledge of outcome data? | Yes | Methylation information was measured before an outcome could occur | Yes | Methylation information was measured before an outcome could occur | Yes | Methylation information was measured before an outcome could occur | Yes | Methylation information was measured before an outcome could occur | Probably yes | Methylation information was measured before an outcome could occur in TCGA | Probably yes | Methylation information was measured before an outcome could occur in TCGA | Probably yes | Methylation information was measured before an outcome could occur in TCGA | Probably yes | Methylation information was measured before an outcome could occur in TCGA | Probably yes | Methylation information was measured before an outcome could occur in TCGA |
| 2.3 Are all predictors available at the time the model is intended to be used? | Yes | Same as above | Yes | Same as above | Yes | Same as above | Yes | Same as above | Probably yes | Same as above | Probably yes | Same as above | Probably yes | Same as above | Probably yes | Same as above | Probably yes | Same as above |
| ***Overall ROB Domain 2*** | ***Low risk of bias*** | | ***Low risk of bias*** | | ***Low risk of bias*** | | ***Low risk of bias*** | | ***Low risk of bias*** | | ***Low risk of bias*** | | ***Low risk of bias*** | | ***Low risk of bias*** | | ***Low risk of bias*** | |
| **DOMAIN 3: Outcome** |  |  |  |  |  |  |  |  |  |  |  |  |  |  |  |  |  |  |
| 3.1 Was the outcome determined appropriately? | Probably yes | The outcome was overall survival, which is a well-established outcome | Probably yes | The outcome was overall survival, which is a well-established outcome | Probably yes | The main outcome was overall survival, which is a well-established outcome | Probably yes | The outcomes of overall survival is a standardized outcome, and disease-free survival was clearly defined. | Probably yes | The outcomes of overall survival is a standardized outcome, and disease-free survival was clearly defined. | Probably yes | The outcomes of overall survival is a standardized outcome, and disease-free survival was clearly defined. | Probably yes | The outcomes of overall survival is a standardized outcome, and disease-free survival was clearly defined. | Probably yes | The outcomes of overall survival is a standardized outcome, and disease-free survival was clearly defined. | Probably yes | The outcomes of overall survival is a standardized outcome, and disease-free survival was clearly defined. |
| 3.2 Was a pre-specified or standard outcome definition used? | Probably yes | Same as above | Probably yes | Same as above | Probably yes | Same as above | Probably yes | Same as above | Probably yes | Same as above | Probably yes | Same as above | Probably yes | Same as above | Probably yes | Same as above | Probably yes | Same as above |
| 3.3 Were predictors excluded from the outcome definition? | Yes | Methylation information was measured before an outcome could occur | Yes | Methylation information was measured before an outcome could occur | Yes | Methylation information was measured before an outcome could occur | Yes | Methylation and RNA information was measured before an outcome could occur | Yes | Methylation and RNA information was measured before an outcome could occur | Yes | Methylation and RNA information was measured before an outcome could occur | Yes | Methylation and RNA information was measured before an outcome could occur | Yes | Methylation and RNA information was measured before an outcome could occur | Yes | Methylation and RNA information was measured before an outcome could occur |
| 3.4 Was the outcome defined and determined in a similar way for all participants? | Probably yes | Follow-up information was collected in a standardized manner in the TCGA | Probably yes | Follow-up information was collected in a standardized manner in the TCGA | Probably yes | Follow-up information was collected in a standardized manner | Probably yes | Follow-up information was collected in a standardized manner in the TCGA | Probably yes | Follow-up information was collected in a standardized manner in the TCGA | Probably yes | Follow-up information was collected in a standardized manner in the TCGA | Probably yes | Follow-up information was collected in a standardized manner in the TCGA | Probably yes | Follow-up information was collected in a standardized manner in the TCGA | Probably yes | Follow-up information was collected in a standardized manner in the TCGA |
| 3.5 Was the outcome determined without knowledge of predictor information? | Yes | Methylation information was measured before an outcome could occur | Yes | Methylation information was measured before an outcome could occur | Yes | Methylation information was measured before an outcome could occur | Yes | Methylation information was measured before an outcome could occur | Yes | Methylation information was measured before an outcome could occur | Yes | Methylation information was measured before an outcome could occur | Yes | Methylation information was measured before an outcome could occur | Yes | Methylation information was measured before an outcome could occur | Yes | Methylation information was measured before an outcome could occur |
| 3.6 Was the time interval between predictor assessment and outcome determination appropriate? | Probably yes | The follow-up time (around 10 years) was long enough to observe the death outcome | Probably yes | The follow-up time (around 10 years) was long enough to observe the outcome | Probably yes | The follow-up time (around 5 yearss) was long enough to observe the death outcome | Probably yes | The follow-up time (around 8 yearss) was long enough to observe the death outcome | Probably yes | The follow-up time (around 10 years) was long enough to observe the death outcome | Probably yes | The follow-up time (around 10 years) was long enough to observe the death outcome | Probably yes | The follow-up time (around 10 years) was long enough to observe the death outcome | Probably yes | The follow-up time (around 10 years) was long enough to observe the death outcome | Probably yes | The follow-up time (around 10 years) was long enough to observe the death outcome |
| ***Overall ROB Domain 3*** | ***Low risk of bias*** | | ***Low risk of bias*** | | ***Low risk of bias*** | | ***Low risk of bias*** | | ***Low risk of bias*** | | ***Low risk of bias*** | | ***Low risk of bias*** | | ***Low risk of bias*** | | ***Low risk of bias*** | |
| **DOMAIN 4: Analysis** |  |  |  |  |  |  |  |  |  |  |  |  |  |  |  |  |  |  |
| 4.1 Were there a reasonable number of participants with the outcome? | No | The number of events per variable = 30 death/349 candidate CpGs = 0.085 is too small | No | The number of death was unknown, but the number of events per variable is definitely very small in that genome-wide CpGs were investigated | No | The number of death was unknown, but the number of events per variable is definitely very small in that genome-wide CpGs were investigated | No | The number of death was unknown, but the number of events per variable is definitely very small in that genome-wide CpGs were investigated | No | The number of death was unknown, but the number of events per variable is definitely very small in that genome-wide CpGs were investigated | No | The number of death was unknown, but the number of events per variable is definitely very small in that genome-wide CpGs were investigated | No | The number of death was unknown, but the number of events per variable is definitely very small in that genome-wide CpGs were investigated | No | The number of death was unknown, but the number of events per variable is definitely very small in that genome-wide CpGs were investigated | No | The number of death was unknown, but the number of events per variable is definitely very small in that genome-wide CpGs were investigated |
| 4.2 Were continuous and categorical predictors handled appropriately? | Probably yes | β values were kept as continuous variables. Though the relationship between the β values of CpGs and risk of death was unexamined, testing all CpGs is impossible, and their relationship is generally non-linear. | Probably yes | β values were kept as continuous variables. Though the relationship between the β values of CpGs and risk of death was unexamined, testing all CpGs is impossible, and their relationship is generally non-linear. | Probably yes | β values were kept as continuous variables. Though the relationship between the β values of CpGs and risk of death was unexamined, testing all CpGs is impossible, and their relationship is generally non-linear. | Probably yes | β values were kept as continuous variables. Though the relationship between the β values of CpGs and risk of death was unexamined, testing all CpGs is impossible, and their relationship is generally non-linear. | Probably yes | β values were kept as continuous variables. Though the relationship between the β values of CpGs and risk of death was unexamined, testing all CpGs is impossible, and their relationship is generally non-linear. | Probably yes | β values were kept as continuous variables. Though the relationship between the β values of CpGs and risk of death was unexamined, testing all CpGs is impossible, and their relationship is generally non-linear. | Probably yes | β values were kept as continuous variables. Though the relationship between the β values of CpGs and risk of death was unexamined, testing all CpGs is impossible, and their relationship is generally non-linear. | Probably yes | β values were kept as continuous variables. Though the relationship between the β values of CpGs and risk of death was unexamined, testing all CpGs is impossible, and their relationship is generally non-linear. | Probably yes | β values were kept as continuous variables. Though the relationship between the β values of CpGs and risk of death was unexamined, testing all CpGs is impossible, and their relationship is generally non-linear. |
| 4.3 Were all enrolled participants included in the analysis? | Probably no | Patients with missing clinical variables were likely to be excluded. | No | Only patients with complete clinical data were selected in the analysis | Yes | All patients meeting the inclusion criteria were included in the analysis | Probably no | Patients with missing clinical variables were likely to be excluded. | Probably no | Patients with missing clinical variables were likely to be excluded. | Probably no | Patients with missing clinical variables were likely to be excluded. | Probably no | Patients with missing clinical variables were likely to be excluded. | Probably yes | Patients with missing values were also shown in Table 3 | No information | No information was provided regarding whether patients with missing data/outliers were excluded |
| 4.4 Were participants with missing data handled appropriately? | Probably no | Same as above | No | Same as above | Yes | Multiple imputation was used | Probably no | Same as above | Probably no | Same as above | Probably no | Same as above | Probably no | Same as above | No information | The 'impute' R package was used, but more details regarding handling of missing data was not reported | No information | No information was provided regarding how the missing data were handled. |
| 4.5 Was selection of predictors based on univariable analysis avoided? | Probably yes | Selection of CpGs was partly based on gene function (immune genes) | No | Selection was entirely based on p-values in univariable and multivariable Cox analyses | Probably yes | Selection was also based on the Brier score for each predictor | Probably yes | Select CpGs located in the promoter region of genes significantly associated with survival | Probably yes | Select differently methylated CpGs associated with differently expressed genes comparing tumor and normal tissues | Probably yes | CpGs were also selected based on standard deviation and clustering methods | No | Selection was entirely based on p-values in univariable and multivariable Cox analyses | Yes | Selection was entirely based on p-values in univariable and multivariable Cox analyses | Probably no | selection was only based on differentially methylation CpGs comparing tumor and normal tissues and importance value obtained by random forest |
| 4.6 Were complexities in the data (e.g. censoring, competing risks, sampling of controls) accounted for appropriately? | Probably yes | Cox regression analysis was used to handle censored data | Probably yes | Cox regression analysis was used to handle censored data | Yes | Cox models were used for time-to-event data, and p-values were adjusted via independent hypothesis weighting | Probably yes | Cox regression analysis was used to handle censored data | Probably yes | Cox regression analysis was used to handle censored data | Probably yes | Cox regression analysis was used to handle censored data | Probably yes | Cox regression analysis was used to handle censored data | Probably yes | Cox regression analysis was used to handle censored data | Probably no | Censored data were likely not to be accounted for when using random forest method |
| 4.7 Were relevant model performance measures evaluated appropriately? | No | Only discrimination was assessed by K-M curves and AUC, whereas calibration was not evaluated. | No | Only discrimination was assessed by K-M curves and AUC, whereas calibration was not evaluated. | Probably no | Discrimination was measured by AUCs, and overall model performance was measured by Brier score. But calibration was not examined. | No | Only discrimination was assessed by K-M curves and AUC, whereas calibration was not evaluated. The calibration was evaluated for another prognostic model consisting of CpG-based prognostic score and age, gender, TNM stage. | No | Only discrimination was assessed by K-M curves, whereas calibration was not evaluated. | No | Only discrimination was assessed by K-M curves and AUC, whereas calibration was not evaluated. The calibration was evaluated for another prognostic model consisting of CpG-based prognostic score plus age and TNM stage. | No | Only discrimination was assessed by K-M curves and AUC, whereas calibration was not evaluated. | No | Only discrimination was assessed by K-M curves and AUC, whereas calibration was not evaluated. The calibration was evaluated for another prognostic model consisting of CpG-based prognostic score plus age, TNM stage, and N stage. | No | Only discrimination was assessed by AUC, whereas calibration was not evaluated. The calibration was evaluated for another prognostic model consisting of CpG-based prognostic T stage, N stage, and lymph node metastasis |
| 4.8 Were model overfitting and optimism in model performance accounted for? | No | Internal validation consists only of a single random split sample of participant data | No | Internal validation consists only of a single random split sample of participant data | Yes | 10-fold internal cross-validation approach with three repetitions was performed | No | Internal validation consists only of a single random split sample of participant data | Probably yes | 5-fold internal cross-validation approach with three repetitions was performed | Yes | The shrinkage technique (regularization) was used | No | Internal validation consists only of a single random split sample of participant data | Yes | The shrinkage technique (regularization) was used | Yes | Random forest with three-fold cross-validation was used to control overfitting |
| 4.9 Do predictors and their assigned weights in the final model correspond to the results from multivariable analysis? | Yes | Both predictors and regression coefficients in the reported final model correspond to the reported results of the multivariable regression analysis | Yes | Both predictors and regression coefficients in the reported final model correspond to the reported results of the multivariable regression analysis | Yes | Both predictors and regression coefficients in the reported final model correspond to the reported results of the multivariable regression analysis | Yes | Both predictors and regression coefficients in the reported final model correspond to the reported results of the multivariable regression analysis | Yes | Both predictors and regression coefficients in the reported final model correspond to the reported results of the multivariable regression analysis | Yes | Both predictors and regression coefficients in the reported final model correspond to the reported results of the multivariable regression analysis | Yes | Both predictors and regression coefficients in the reported final model correspond to the reported results of the multivariable regression analysis | Yes | Both predictors and regression coefficients in the reported final model correspond to the reported results of the multivariable regression analysis | Yes | Both predictors and regression coefficients in the reported final model correspond to the reported results of the multivariable regression analysis |
| ***Overall ROB Domain 4*** | ***High risk of bias*** | | ***High risk of bias*** | | ***High risk of bias*** | | ***High risk of bias*** | | ***High risk of bias*** | | ***High risk of bias*** | | ***High risk of bias*** | | ***High risk of bias*** | | ***High risk of bias*** | |

**References**

1.	d'Errico M, Alwers E, Zhang Y, Edelmann D, Brenner H, Hoffmeister M. Identification of prognostic DNA methylation biomarkers in patients with gastrointestinal adenocarcinomas: A systematic review of epigenome-wide studies. *Cancer Treat Rev* 2020; **82**: 101933.

2.	Moons KGM, Wolff RF, Riley RD, et al. PROBAST: A Tool to Assess Risk of Bias and Applicability of Prediction Model Studies: Explanation and Elaboration. *Ann Intern Med* 2019; **170**(1): W1-W33.

3.	Gündert M, Edelmann D, Benner A, et al. Genome-wide DNA methylation analysis reveals a prognostic classifier for non-metastatic colorectal cancer (ProMCol classifier). *Gut* 2019; **68**(1): 101-10.

4.	Bläker H, Alwers E, Arnold A, et al. The Association Between Mutations in BRAF and Colorectal Cancer-Specific Survival Depends on Microsatellite Status and Tumor Stage. *Clin Gastroenterol Hepatol* 2019; **17**(3): 455-62.

5.	Hoffmeister M, Jansen L, Rudolph A, et al. Statin use and survival after colorectal cancer: the importance of comprehensive confounder adjustment. *J Natl Cancer Inst* 2015; **107**(6): djv045.

6.	Brenner H, Chang-Claude J, Jansen L, Knebel P, Stock C, Hoffmeister M. Reduced risk of colorectal cancer up to 10 years after screening, surveillance, or diagnostic colonoscopy. *Gastroenterology* 2014; **146**(3): 709-17.

7.	Alwers E, Bläker H, Walter V, et al. External validation of molecular subtype classifications of colorectal cancer based on microsatellite instability, CIMP, BRAF and KRAS. *BMC Cancer* 2019; **19**(1): 681.

8.	van Buuren S, K. G-O. mice: Multivariate Imputation by Chained Equations in R. *J Stat Softw* 2011; **45**: 1–67.

9.	Jia M, Zhang Y, Jansen L, et al. A prognostic CpG score derived from epigenome-wide profiling of tumor tissue was independently associated with colorectal cancer survival. *Clin Epigenetics* 2019; **11**(1): 109.

10.	Yang X, Gao L, Zhang S. Comparative pan-cancer DNA methylation analysis reveals cancer common and specific patterns. *Brief Bioinform* 2017; **18**(5): 761-73.

11.	Hou X, He X, Wang K, et al. Genome-Wide Network-Based Analysis of Colorectal Cancer Identifies Novel Prognostic Factors and an Integrative Prognostic Index. *Cell Physiol Biochem* 2018; **49**(5): 1703-16.

12.	Yang CS, Zhang Y, Xu XQ, Li WH. Molecular subtypes based on DNA methylation predict prognosis in colon adenocarcinoma patients. *Aging (Albany NY)* 2019; **11**(24): 11880-92.

13.	Gong S, Ye W, Liu T, Jian S, Liu W. The Development of Three-DNA Methylation Signature as a Novel Prognostic Biomarker in Patients with Colorectal Cancer. *Biomed Res Int* 2020; **2020**: 3497810.

14.	Wang X, Wang D, Liu J, Feng M, Wu X. A novel CpG-methylation-based nomogram predicts survival in colorectal cancer. *Epigenetics* 2020; **15**(11): 1213-27.

15.	Wang XY, Zhang DS, Zhang C, Sun YM. Identification of epigenetic methylation-driven signature and risk loci associated with survival for colon cancer. *Ann Transl Med* 2020; **8**(6): 324.

16.	Wang Y, Zhang M, Hu X, Qin W, Wu H, Wei M. Colon cancer-specific diagnostic and prognostic biomarkers based on genome-wide abnormal DNA methylation. *Aging (Albany NY)* 2020; **12**(22): 22626-55.

17.	Xiang R, Fu T. Gastrointestinal adenocarcinoma analysis identifies promoter methylation-based cancer subtypes and signatures. *Sci Rep* 2020; **10**(1): 21234.

18.	Yang H, Jin W, Liu H, et al. A novel prognostic model based on multi-omics features predicts the prognosis of colon cancer patients. *Mol Genet Genomic Med* 2020; **8**(7): e1255.

19.	Yin ZJ, Yan XM, Wang QM, et al. Detecting Prognosis Risk Biomarkers for Colon Cancer Through Multi-Omics-Based Prognostic Analysis and Target Regulation Simulation Modeling. *Front Genet* 2020; **11**: 524.

20.	Chen F, Pei LJ, Liu SY, et al. Identification of a Novel Immune-Related CpG Methylation Signature to Predict Prognosis in Stage II/III Colorectal Cancer. *Front Genet* 2021; **12**: 684349.

21.	Huang H, Zhang L, Fu J, et al. Development and validation of 3-CpG methylation prognostic signature based on different survival indicators for colorectal cancer. *Mol Carcinog* 2021; **60**(6): 403-12.

22.	Li DH, Du XH, Liu M, Zhang R. A 10-gene-methylation-based signature for prognosis prediction of colorectal cancer. *Cancer Genet* 2021; **252-253**: 80-6.

23.	Xin JY, Wu YL, Ben S, et al. CoSMeD: a user-friendly web server to estimate 5-year survival probability of left-sided and right-sided colorectal cancer patients using molecular data. *Bioinformatics* 2022; **38**(1): 278-81.
